# Genetically Adjusted PSA Levels for Prostate Cancer Screening

**DOI:** 10.1101/2022.04.18.22273850

**Authors:** Linda Kachuri, Thomas J. Hoffmann, Yu Jiang, Sonja I. Berndt, John P. Shelley, Kerry Schaffer, Mitchell J. Machiela, Neal D. Freedman, Wen-Yi Huang, Shengchao A. Li, Ryder Easterlin, Phyllis J. Goodman, Cathee Till, Ian Thompson, Hans Lilja, Stephen K. Van Den Eeden, Stephen J. Chanock, Christopher A. Haiman, David V. Conti, Robert J. Klein, Jonathan D. Mosley, Rebecca E. Graff, John S. Witte

**Affiliations:** Department of Epidemiology & Biostatistics, University of California San Francisco, San Francisco, CA, USA; Department of Epidemiology & Population Health, Stanford University School of Medicine, Stanford, CA, USA; Stanford Cancer Institute, Stanford University School of Medicine, Stanford, CA, USA; Institute of Human Genetics, University of California San Francisco, San Francisco, CA, USA; Division of Cancer Epidemiology and Genetics, National Cancer Institute, Rockville, MD, USA; Department of Biomedical Informatics, Vanderbilt University Medical Center, Nashville, TN, USA; Division of Hematology and Oncology, Vanderbilt University Medical Center, Nashville, TN, USA; Biological and Medical Informatics, University of California San Francisco, San Francisco, CA, USA; Fred Hutchinson Cancer Research Center, Seattle, WA, USA; SWOG Statistics and Data Management Center, Fred Hutchinson Cancer Research Center, Seattle, WA, USA; CHRISTUS Santa Rosa Medical Center Hospital, San Antonio, TX, USA; Departments of Laboratory Medicine, Surgery, Medicine, Memorial Sloan Kettering Cancer Center, New York, NY, USA; Department of Translational Medicine, Lund University, Skåne University Hospital, Malmö, Sweden; Division of Research, Kaiser Permanente Northern California, Oakland, CA, USA; Center for Genetic Epidemiology, Department of Population and Preventive Health Sciences, Keck School of Medicine, University of Southern California, Los Angeles, CA, USA; Norris Comprehensive Cancer Center, Keck School of Medicine, University of Southern California, Los Angeles, CA, USA; Department of Genetics and Genomic Sciences, Icahn School of Medicine at Mount Sinai, New York, NY, USA; Department of Internal Medicine, Vanderbilt University Medical Center, Nashville, TN, USA; Departments of Biomedical Data Science and Genetics (by courtesy), Stanford University, Stanford, CA, USA

## Abstract

Prostate-specific antigen (PSA) screening for prostate cancer remains controversial because it increases overdiagnosis and overtreatment of clinically insignificant tumors. Accounting for genetic determinants of constitutive, non-cancer-related PSA variation has potential to improve screening utility. We discovered 128 genome-wide significant associations (*P*<5×10^-8^) in a multi-ancestry meta-analysis of 95,768 men and developed a PSA polygenic score (PGS_PSA_) that explains 9.61% of constitutive PSA variation. We found that in men of European ancestry, using PGS-adjusted PSA would avoid 31% of negative prostate biopsies, but also result in 12% fewer biopsies in patients with prostate cancer, mostly with Gleason score <7 tumors. Genetically adjusted PSA was more predictive of aggressive prostate cancer (odds ratio (OR)=3.44, *P*=6.2×10^-14^; AUC=0.755) than unadjusted PSA (OR=3.31, *P*=1.1×10^-12^; AUC=0.738) in 106 cases and 23,667 controls. Compared to a prostate cancer PGS alone (AUC=0.712), including genetically adjusted PSA improved detection of aggressive disease (AUC=0.786, *P*=7.2×10^-4^). Our findings highlight the potential utility of incorporating PGS for personalized biomarkers in prostate cancer screening.

## MAIN

Prostate-specific antigen (PSA) is an enzyme produced by the prostate gland, which degrades gel-forming seminal proteins to release of motile sperm and is encoded by the kallikrein-3 (*KLK3*) gene^1-3^. As prostate epithelial tissue becomes disrupted by a tumor, greater PSA concentrations are released into circulation^2,3^. PSA levels can also rise due to prostatic inflammation, infection, benign prostatic hyperplasia, older age, and increased prostate volume^3-5^. Increased body mass index is associated with lower PSA levels, but the underlying mechanisms remain unclear^6,7^. Low PSA levels thus do not rule out prostate cancer and PSA elevation is not sufficient for a conclusive diagnosis^8^. Although PSA testing reduces deaths from prostate cancer^9^, between 20 and 60% of cancers detected using PSA testing are estimated to be overdiagnoses^10-12^. In addition, the long-term risk of lethal prostate cancer remains low, especially in men with PSA below the age-specific median^13,14^. As a result, clinical guidelines in the United States and globally advise against population-level PSA screening and promote a shared decision-making model^15,16^.

One avenue for refining PSA screening is by accounting for variability in PSA due to genetic factors. PSA is highly heritable, with 40 independent loci identified in the largest previous genome-wide association study (GWAS) ^17,18^. The goal of genetically correcting PSA levels is to increase the relative variation in PSA attributable to prostate cancer, thereby improving their predictive value for disease detection. The first study to genetically correct PSA using just four variants reclassified 3% of participants to warranting biopsy and 3% to avoiding biopsy^19^. Incorporating additional genetic predictors has the potential to personalize PSA testing, reduce overdiagnosis-related morbidity, and improve detection of lethal disease. To maximize the utility of this approach, it is critical to distinguish genetic variants that influence constitutive PSA levels from those affecting prostate tumor development. PSA and prostate cancer share many genetic loci^17,19-22^, but the extent to which this overlap reflects screening bias remains unclear, as GWAS of prostate cancer may capture signals for disease susceptibility and incidental detection due to benign PSA elevation.

Our study explores the genetic architecture of PSA levels in men without prostate cancer, with a view toward assessing whether genetic adjustment of PSA improves clinical decision-making related to prostate cancer diagnosis. It also provides a novel framework for the clinical translation of polygenic scores (PGS) for non-causal cancer biomarkers.

## RESULTS

The study design of the Precision PSA study is illustrated in **Figure 1**. Using data from five studies **(Methods)**, we conducted genome-wide analyses of PSA levels ≤10 ng/mL in cis-gender men never diagnosed with prostate cancer. GWAS results were meta-analyzed within ancestry groups and then combined across populations for a total sample size of 95,768 individuals.

**Fig. 1:**
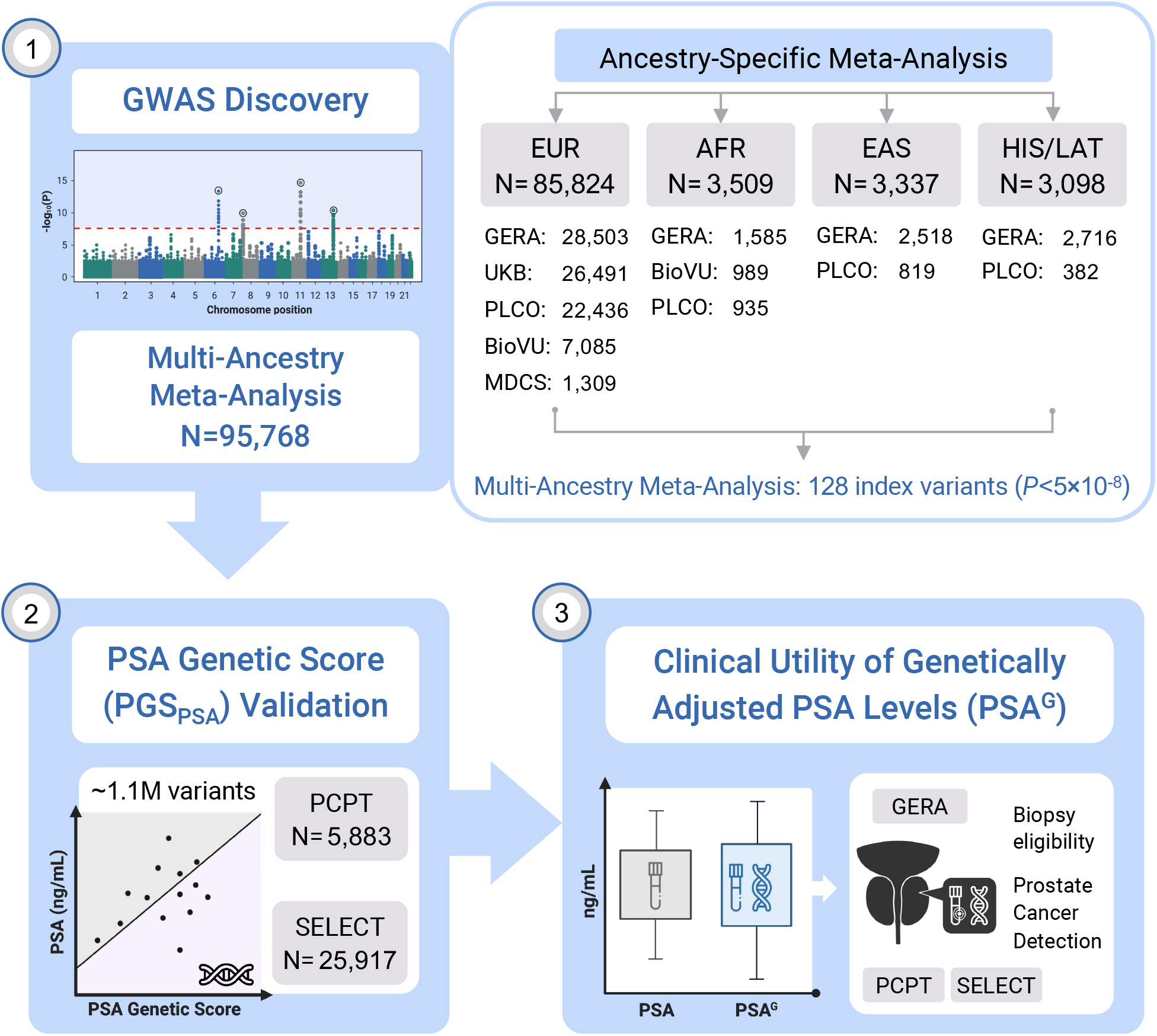
Overview of the Precision PSA study design. Genome-wide association analyses were conducted in men without prostate cancer and meta-analyzed within each population: European ancestry (EUR), African ancestry (AFR), East Asian ancestry (EAS) and Hispanic/Latino (HIS/LAT). Ancestry-stratified results were used to develop a genome-wide PSA genetic score (PGS_PSA_) comprised of approximately 1.1 million variants and were also combined into a multi-ancestry meta-analysis of 95,768 men. PGS_PSA_ was validated in the Prostate Cancer Prevention Trial (PCPT) and the Selenium and Vitamin E Cancer Prevention Trial (SELECT), and was used to compute genetically adjusted PSA values (PSA^G^). We examined how using PSA^G^ values affects eligibility for prostate biopsy and evaluated associations with incident prostate cancer.

### Genetic Architecture of PSA Variation

The heritability (h^2^) of PSA levels was investigated using several methods to assess sensitivity to underlying modeling assumptions **(Methods)**. Across 26,491 men of European ancestry in the UK Biobank (UKB) with linked clinical records, the median PSA value was 2.35 ng/mL **(Supplementary Fig. 1)**. Using individual-level data for variants with minor allele frequency (MAF)≥0.01 and imputation INFO>0.80, PSA heritability was h^2^=0.41 (95% CI: 0.36-0.46) based on GCTA^23^ and h^2^=0.30 (95% CI: 0.26-0.33) based on LDAK^24^ **(Supplementary Table 1; Extended Data Fig. 1)**. Applying LDAK to GWAS summary statistics generated from the same individuals produced similar estimates (h^2^=0.35, 0.28-0.43), whereas other methods^25,26^ were biased downward. In the European ancestry GWAS meta-analysis (N_EUR_=85,824), LDAK estimated h^2^=0.30 (95% 0.29-0.31). Sample sizes for other ancestries were too small for reliable heritability estimates.

The multi-ancestry meta-analysis of 95,768 men from five studies identified 128 independent index variants (*P*<5.0×10^-8^, linkage disequilibrium (LD) *r*^2^<0.01 within ±10 Mb windows) across 90 chromosomal cytoband regions **(Figure 2)**. The strongest associations were in known PSA loci^17,19,21,22^, such as *KLK3* (rs17632542, *P*=3.2×10^-638^), 10q26.12 (rs10886902, *P*=8.2×10^-118^), *MSMB* (rs10993994, *P*=7.3×10^-87^), *NKX3-1* (rs1160267, *P*=6.3×10^-83^), *CLPTM1L* (rs401681, *P*=7.0×10^-54^), and *HNF1B* (rs10908278, *P*=2.1×10^-46^). Eighty-two index variants were independent of previously detected associations in the Genetic Epidemiology Research on Adult Health and Aging (GERA) cohort^17^; they mapped to 56 cytobands where PSA signals have not previously been reported. Associations initially detected in the UKB **(Extended Data Fig. 1b)** strengthened in the meta-analysis: *TEX11* in Xq13.1 (rs62608084, *P*=1.7×10^-24^); *THADA* in 2p21 (rs11899863, *P*=1.7×10^-13^); *OTX1* in 2p15 (rs58235267, *P*=4.9×10^-13^); *SALL3* in 18q23 (rs71279357, *P*=1.8×10^-12^); and *ST6GAL1* in 3q27.3 (rs12629450, *P*=2.6×10^-10^). Additional novel findings included *CDK5RAP1* (rs291671, *P*=1.2×10^-18^), *LDAH* (rs10193919, *P*=1.5×10^-15^), *ABCC4* (rs61965887, *P*=3.7×10^-14^), *INKA2* (rs2076591, *P*=2.6×10^-13^), *SUDS3* (rs1045542, *P*=1.2×10^-13^), *FAF1* (rs12569177, *P*=3.2×10^-13^), *JARID2* (rs926309, *P*=1.6×10^-12^), *GPC3* (rs4829762, *P*=5.9×10^- 12^), *EDA* (rs2520386, *P*=4.2×10^-11^), and *ODF3* (rs7103852, *P*=1.2×10^-9^) **(Supplementary Tables 2-3)**.

**Fig. 2:**
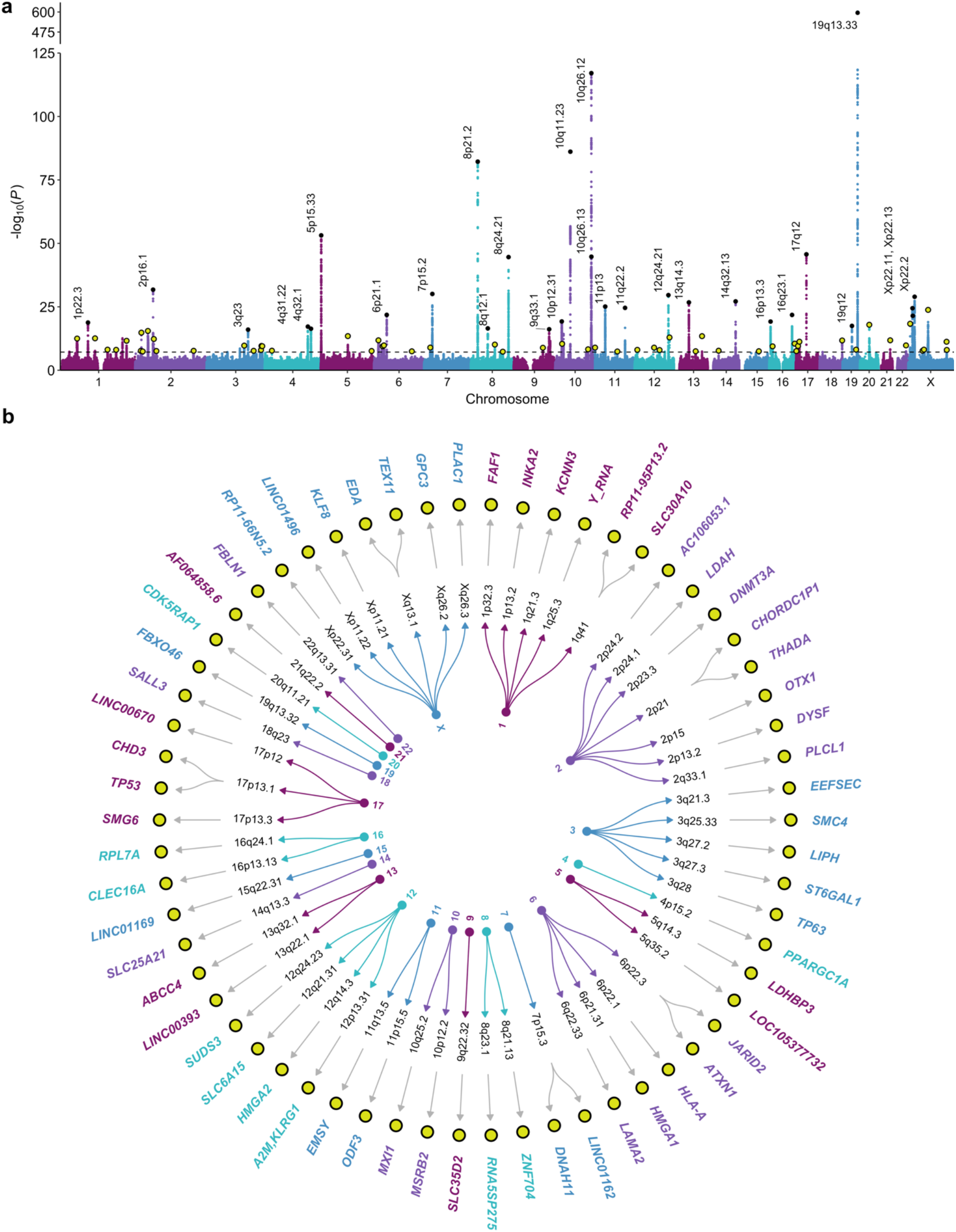
Multi-ancestry genome-wide association study (GWAS) of PSA levels. **a**, Manhattan plot depicting the results of the GWAS meta-analysis of PSA levels in 95,768 men without prostate cancer. The genome-wide significance threshold of *P*<5×10^-8^ is indicated by the dotted black line. Index variants within known PSA-associated loci are annotated with the corresponding cytoband. Novel findings are highlighted in yellow. **b**, Circular dendogram shows the nearest gene(s) for novel PSA-associated variants. Genome-wide significant (*P*<5×10^-8^) index variants were selected using linkage disequilibrium (LD)-based clumping (LD r^2^<0.01 within 10Mb windows). All GWAS p-values are two-sided and derived from a fixed-effects inverse-variance-weighted meta-analysis using METAL.

Of the 128 index variants, 96 reached genome-wide significance in the European ancestry meta-analysis, as did three in the East Asian ancestry meta-analysis (*KLK3*: rs2735837, rs374546878; *MSMB*: rs10993994; N_EAS_=3,337), two in the Hispanic/Latino meta-analysis (*KLK3*: rs17632542, rs2735837; N_HIS/LAT_=3,098), and one (*FGFR2*: rs10749415; N_AFR_=3,509) in the African ancestry meta-analysis **(Supplementary Table 4)**. Effect sizes from the European ancestry GWAS were modestly correlated with estimates from other ancestries (Spearman’s ρ_HIS/LAT_=0.48, *P*=1.1×10^-8^; ρ_AFR_=0.27, *P*=2.0×10^-3^; ρ_EAS_=0.16, *P*=0.068) **(Supplementary Fig. 2)**. However, cross-population comparisons of correlations should be interpreted with caution as they are confounded by higher sampling error in groups with smaller sample sizes.

There was heterogeneity (Cochran’s Q *P*_Q_<0.05) across ancestry-specific fixed-effects meta-analyses for 12 out of 128 index variants, four of which had effects in different directions: rs58235267 (*OTX1*), rs1054713 (*KLK1*), rs10250340 (*EIF4HP1*), and rs7020681 (*SLC35D2*) **(Supplementary Table 5)**. An alternative meta-analysis approach, MR-MEGA^27^, which partitions effect size heterogeneity into components correlated with ancestry and residual variation, identified one additional signal in 5q15 (rs291812, *P*_MR-MEGA_=1.0×10^-8^) that was driven by East Asian ancestry (*P*_EAS_=1.2×10^-6^; **Supplementary Table 6)**.

Predicted functional consequences of the 128 index variants were explored using CADD^28^ scores >13 (corresponding to the 5% most deleterious substitutions) were observed for 16 out of the 128 index variants detected with the original approach, including 10 new signals: rs10193919 (*LDAH*); rs7732515 in 5q14.3; rs11899863 (*THADA*); rs58235267 (*OTX1*); rs926309 (*JARID2*); rs4829762 (*GPC3*), rs13268, a missense variant in *FBLN1*; rs78378222 in *TP53*, rs3760230 in *SMG6*; and rs712329 in *SLC25A21* **(Supplementary Table 7)**. Sixty-one variants had significant (FDR<0.05) effects on gene expression, including 15 prostate tissue eQTLs for 17 eGenes, 55 blood eQTLs for 185 eGenes, and 9 eQTLs with effects in both tissues. Notable eGenes included *RUVBL1*, a chromatin-remodeling factor that modulates pro-inflammatory NF-κB signaling and transcription of Myc and β-catenin^29^; *ODF3*, which maintains elastic structures in the sperm tail^30^; and *LDAH*, which promotes cholesterol mobilization in macrophages^31^. Several PSA-associated variants were eQTLs for genes involved in immune response (*IFITM2, IFITM3, HS1BP3*).

### Impact of PSA-Related Selection Bias on Prostate Cancer GWAS

Since prostate cancer detection often hinges on PSA elevation, genetic factors resulting in higher constitutive PSA levels may appear to increase prostate cancer risk because of more frequent screening. Of the 128 lead PSA variants, 52 (41%) were associated with prostate cancer at the Bonferroni-corrected threshold (p<0.05/128) in the PRACTICAL consortium’s European ancestry GWAS^32^ **(Supplementary Table 8)**. Using the method by Dudbridge et al.^33^, we investigated whether index event bias could partly explain these shared signals^33,34^ **(Methods; Figure 3; Supplementary Table 9)**. Applying the estimated bias correction factor (*b*=1.144) decreased the number of variants associated with prostate cancer from 52 to 34 **(Extended Data Fig. 2)**. When we corrected 209 European ancestry prostate cancer risk variants (*P*<5.0×10^-8^; LD *r*^2^<0.01) for screening bias, 93 (45%) remained genome-wide significant. Notably, rs76765083 (*KLK3*) remained genome-wide significant, but reversed direction. Sensitivity analyses using SlopeHunter^35^ resulted in 150 (72%) variants with *P*<5×10^-8^ **(Supplementary Table 10)**.

**Fig. 3:**
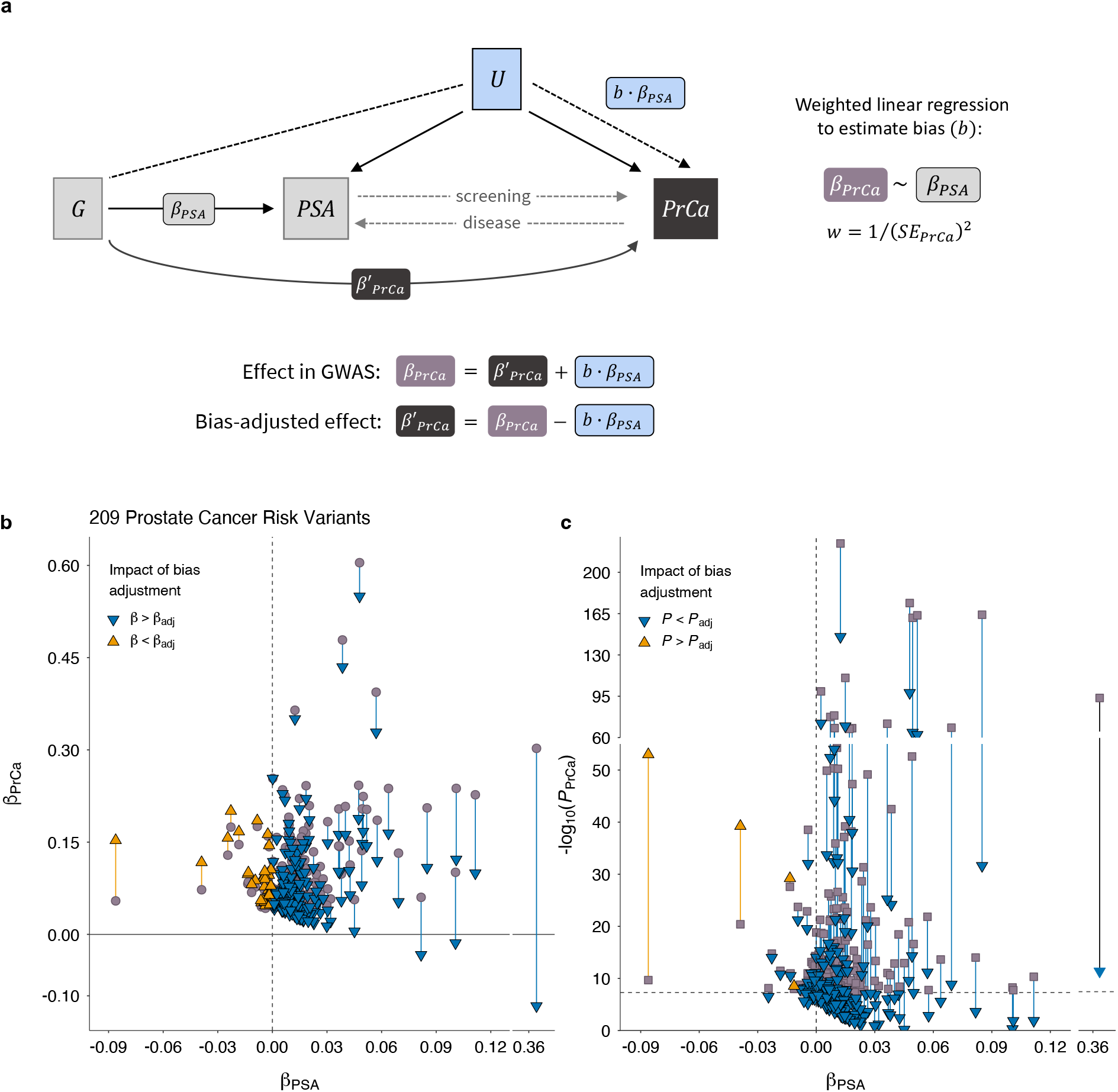
Influence of PSA-related index event bias on prostate cancer GWAS. **a**, Conceptual diagram depicts how selection on PSA levels induces an association between genetic variant G and U, a composite confounder that captures polygenic and non-genetic factors. This selection induces an association with prostate cancer (PrCa) via path G – U → PrCa, in addition to the direct G → PrCa effects. Bi-directional dotted lines show that PSA is not only a disease biomarker, but also influences screening behavior and the likelihood of prostate cancer detection. **b-c**, The impact of bias correction is shown for 209 prostate cancer risk variants. Independent risk variants were selected from the PRACTICAL GWAS meta-analysis (85,554 cases and 91,972 controls of European ancestry) by Conti et al.^32^ using linkage disequilibrium (LD) clumping (LD *r*^2^<0.01, *P*<5×10^-8^). For each variant, associations with PSA (β_PSA_) are based on an inverse-variance-weighted fixed-effects meta-analysis in men of European ancestry (n=85,824). **b**, GWAS effect sizes for prostate cancer (β_PrCa_) are aligned to the risk-increasing allele. Bias-adjusted effect sizes (β_adj_) are denoted by triangles **c**, Two-sided GWAS p-values for prostate cancer (*P*_PrCa_) were derived from an inverse-variance-weighted fixed-effects meta-analysis. Two-sided bias-adjusted p-values (*P*_adj_), denoted by triangles, were calculated from a chi-squared test statistic based on β_adj_ and corresponding standard errors. Genome-wide significance threshold (*P*<5×10^-8^) is indicated by the dotted line.

### Development and Validation of PGS_PSA_

We considered two approaches for constructing a PGS for PSA: clumping genome-wide significant associations from the multi-ancestry meta-analysis (PGS_128_) and a genome-wide score generated using the Bayesian PRS-CSx algorithm (PGS_CSx_)^36^ **(Methods)**. Each score was validated in the Prostate Cancer Prevention Trial (PCPT) and Selenium and Vitamin E Cancer Prevention Trial (SELECT), which were excluded from the discovery GWAS. Most of the men in both cohorts were of European ancestry, although SELECT offered larger sample sizes for other ancestry groups **(Extended Data Fig. 3)**. PGS_CSx_ was ultimately selected, as it was more predictive of baseline PSA than PGS_128_ in multi-ancestry analyses and most ancestry subgroups **(Supplementary Table 11**).

In PCPT, PGS_CSx_ accounted for 8.13% of variation in baseline PSA levels (β per SD increase = 0.186, *P*=3.3×10^-112^) in the pooled multi-ancestry sample of 5883 men **(Fig. 4a-c; Supplementary Table 11)**. PGS_CSx_ was associated with PSA across age groups, although effects attenuated in participants aged ≥70 **(Extended Data Fig. 4)**. PGS_CSx_ was validated in 5725 participants of European (EUR≥0.80) ancestry (PGS_CSx_: β=0.194, *P*=1.7×10^-115^), but neither PGS_128_ nor PGS_CSx_ reached nominal significance in the admixed European and African ancestry (0.20<AFR/EUR<0.80; n=103) or East Asian ancestry (EAS≥0.80; n=55) populations.

**Fig. 4:**
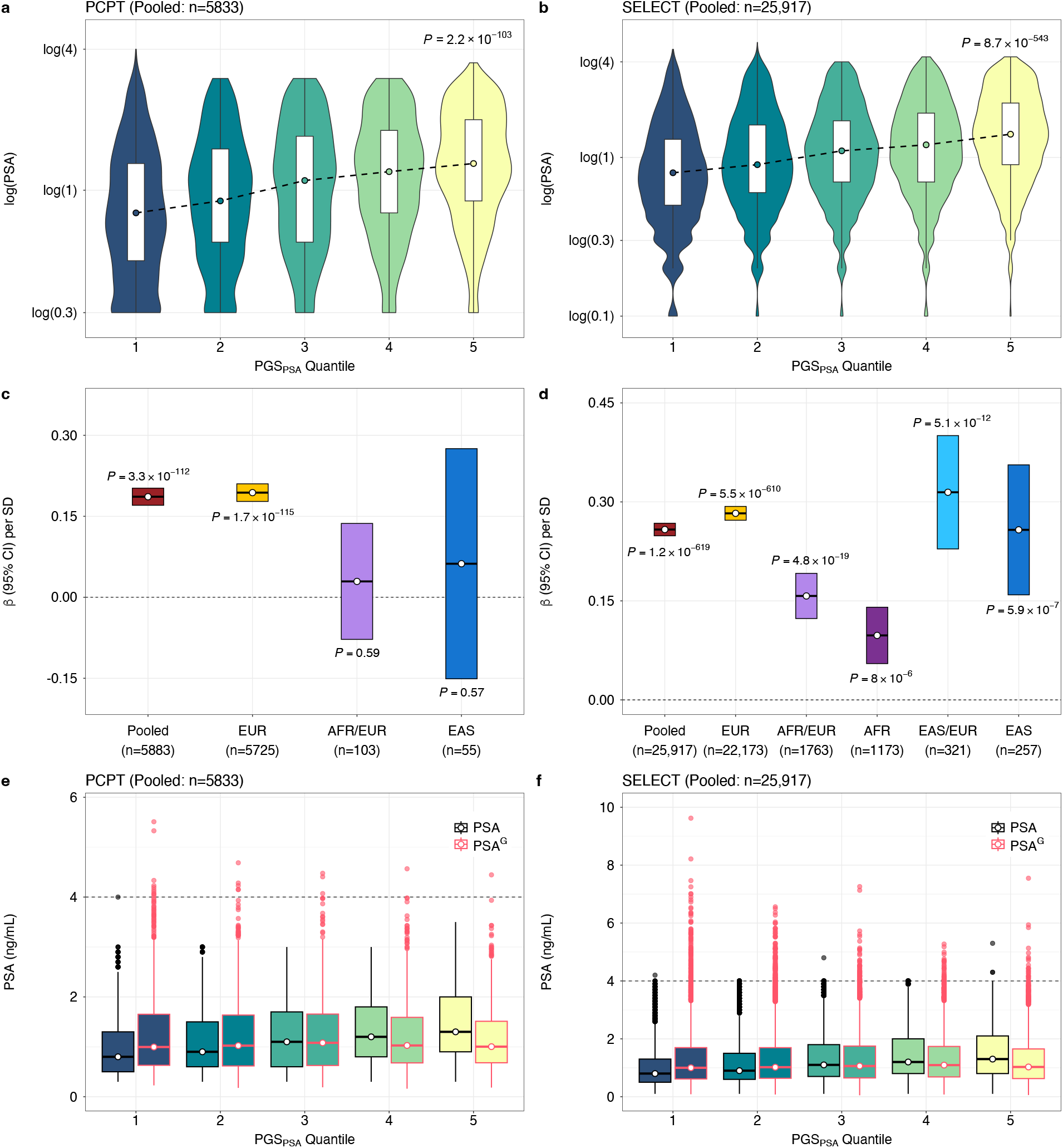
Validation of the polygenic score for PSA (PGS_PSA_) in two cancer prevention trials. Performance of PGS_PSA_ was evaluated in the Prostate Cancer Prevention Trial (PCPT) and Selenium and Vitamin E Cancer Prevention Trial (SELECT). Violin plots show the distribution of baseline log(PSA) within quantiles of PGS_PSA_, comprised of 1,058,173 and 1,071,278 variants in **a**, PCPT and **b**, SELECT. Box plots extend from the 25^th^ to the 75^th^ percentiles, with a trend line connecting the median value within each age stratum. Two-sided p-values are derived from linear regression models for the effect of a quantile increase in PGS_PSA_ on log(PSA). **c-d**, Crossbar plots show the effect estimates (β) and corresponding 95% intervals per standard deviation (SD) increase in the standardized PGS_PSA_ on baseline log(PSA) **c**, PCPT and **d**, SELECT. Ancestry-stratified and pooled, multi-ancestry estimates are presented. Two-sided p-values based on linear regression models are annotated. **e-f**. Comparison of distributions for PSA and genetically adjusted PSA (PSA^G^), with the horizontal line at 4 ng/mL, a commonly used threshold for further diagnostic testing. Box plots show the median value with lower and upper hinges corresponding to the 25^th^ to the 75^th^ percentiles, or first and third quartiles. Whiskers extend as a multiple of the inter-quartile range (IQR*1.5). Outlying values beyond the end of the whiskers are plotted individually.

In SELECT, PGS_CSx_ was associated with baseline PSA levels in the pooled sample of 25,917 men (β=0.258, *P*=1.3×10^-619^) and among men of European ancestry (β_PGS_=0.283, *P*=5.5×10^-610^; n=22,253), accounting for 9.61% to 10.94% of variation, respectively **(Fig. 4b-d; Supplementary Table 11)**. PGS_CSx_ also validated in the East Asian (n=257) (β=0.258, *P*=5.9×10^-7^) and admixed EAS/EUR (n=321; β=0.315, *P*=5.2×10^-12^) ancestry groups. In men with admixed AFR/EUR ancestry (n=1763), PGS_CSx_ explained 4.22% of PSA variation (β=0.157, *P*=4.8×10^-19^). PGS_128_ was more predictive than PGS_CSx_ (β=0.163, *P*=8.2×10^-11^ vs. β=0.098, *P*=8.0×10^-6^) in men of African ancestry (AFR≥0.80; n=1173) and the pooled AFR and admixed (0.20<EUR/AFR<0.80) group n=2936).

We also examined associations with temporal trends in PSA: velocity, calculated using log(PSA) values at two time points, and doubling time in months **(Methods; Supplementary Table 12)**. In men with a PSA increase (SELECT pooled sample: n=14,908), PGS_CSx_ was associated with less rapid velocity (PGS_CSx_: β= -4.06×10^-4^, *P*=3.7×10^-5^;) and longer doubling time (PGS_CSx_: β=10.41, *P*=1.9×10^-8^). In men with a PSA decrease between the first and last time point (SELECT pooled sample: n=6970), PGS_CSx_ was only suggestively associated with slowing PSA decline (β=5.02×10^-4^, *P*=0.068). The same pattern was observed in PCPT, with higher PGS_CSx_ values conferring less rapid changes in PSA.

PGS_CSx_, referred to as PGS_PSA_ from here onward, was used to genetically adjust baseline or earliest pre-randomization PSA values (PSA^G^) for each individual, relative to the population mean **(Methods, Equations 1-2)**. PSA^G^ and unadjusted PSA were strongly correlated in PCPT (Pearson’s *r*=0.841, 0.833-0.848) and SELECT (*r*=0.854, 0.851-0.857). The number of participants with PSA^G^>4 ng/mL, a commonly used threshold for diagnostic testing, increased from 0 to 24 in PCPT and 5 to 413 in SELECT **(Fig. 4e-f)**, reflecting the preferential trial selection of men with low PSA^8,37^.

### Impact of PSA-Related Bias on PGS Associations

In men of European ancestry UKB excluded from the PSA GWAS, there was a strong positive relationship between the 269-variant prostate cancer PGS (PGS_269_)^32^ and PGS_PSA_ in cases (β=0.190, *P*=2.3×10^-96^; n=11,568) and controls (β=0.236, *P*<10^-700^; n=152,884) **(Extended Data Fig. 5, Supplementary Table 13)**. Refitting PGS_269_ using weights corrected for index event bias (PGS_269_^adj^) substantially attenuated associations in cases (β_adj_=0.029, *P*=2.7×10^-3^) and controls (β_adj_=0.052, *P*=2.2×10^-89^).

**Fig. 5:**
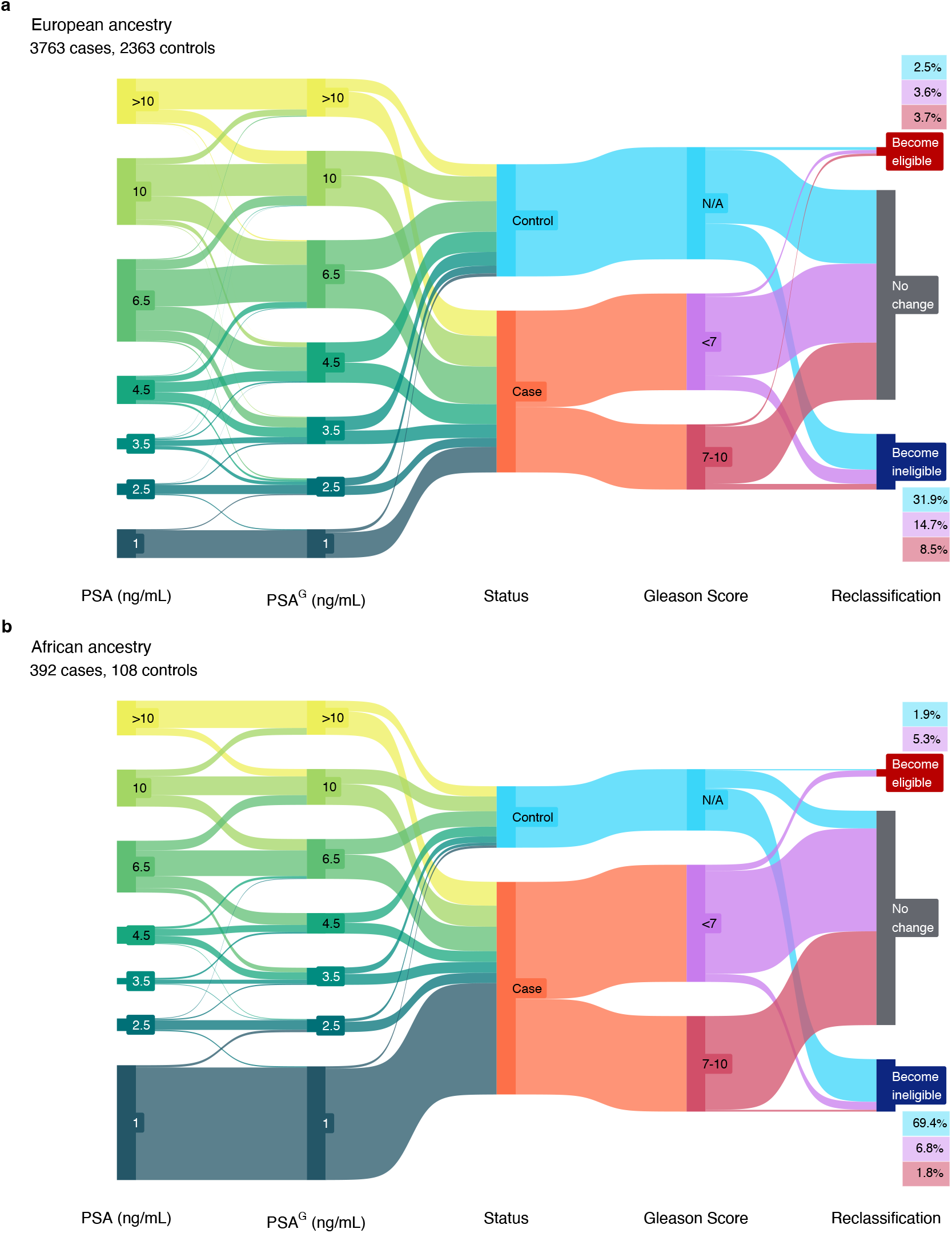
Genetically adjusted PSA influences biopsy eligibility. Flow diagrams illustrate changes in PSA values after genetic adjustment for participants in the Genetic Epidemiology Research in Adult Health and Aging (GERA) cohort, and subsequent reclassification at PSA thresholds used to recommend prostate biopsy. Genetic adjustment was applied to the last pre-biopsy PSA value to obtain PSA^G^. Analyses were performed separately in men of **a**, European and **b**, African ancestry. Size of the nodes and flows are proportional to the number of individuals in each category. Prostate cancer patients (cases) were stratified by Gleason score categories, where Gleason <7 represents potentially indolent disease. Gleason score is not applicable to men with a negative prostate biopsy (controls).

To further characterize the impact of this bias, we examined PGS_269_ associations with prostate cancer status in 3673 cases and 2363 biopsy-confirmed, European ancestry controls from GERA. PGS_269_^adj^ had a larger magnitude of association with prostate cancer (OR for top decile=3.63, 95% CI: 3.01-4.37) than PGS_269_ (OR=2.71, 2.28-3.21) and higher area under the curve (AUC: 0.685 vs. 0.677, *P*=3.91×10^-3^) **(Supplementary Table 14)**. The impact of bias correction was pronounced for Gleason ≥7 tumors (PGS_269_^adj^ AUC=0.692 vs.

PGS_269_ AUC=0.678, *P*=1.91×10^-3^), although these AUC estimates are inflated due to overlap with the GWAS used to develop PGS_26932_. In case-only analyses, PGS_PSA_ and PGS_269_ were inversely associated with Gleason score, illustrating how screening bias decreases the likelihood of identifying high-grade disease **(Supplementary Table 15)**. Compared to Gleason ≤6 tumors, an SD increase in PGS_PSA_ was inversely associated with Gleason 7 (OR=0.79, 0.76-0.83) and Gleason ≥8 disease (OR=0.71, 0.64-0.81). Patients in the top decile of PGS_269_ were approximately 30% less likely to have Gleason ≥8 (OR=0.72, 0.54-0.96) than Gleason ≤6 tumors, but this association was attenuated after bias correction (PGS_269_^adj^: OR=0.94, 0.75-1.17).

### Impact of Genetic Adjustment of PSA on Biopsy Eligibility

Among GERA participants who underwent prostate biopsy, we examined how adjustment using PGS_PSA_ reclassified individuals for biopsy recommendation at age-specific thresholds used by Kaiser Permanente: 40-49 years old=2.5 ng/ml; 50-59 years old=3.5 ng/ml; 60-69 years old=4.5 ng/ml; and 70-79 years old=6.5 ng/ml **(Methods)**. For men of European ancestry, mean PSA levels in men with a negative biopsy (7.2 ng/mL, n=2363) were higher than in men without prostate cancer who did not have a biopsy (1.5 ng/mL; n=24,811) **(Supplementary Table 16)**. Relative to all controls, where standardized 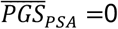, biopsied men were enriched for PSA-increasing alleles (cases: 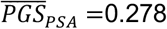; controls: 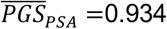). After genetic adjustment, 31.7% of biopsy-negative men were reclassified below the PSA level for recommending biopsy, and 2.5% became biopsy-eligible, resulting in a net reclassification of 29.3% (27.5% to 31.21%) **(Fig. 5a)**. Among 3673 cases PSA^G^ values below the biopsy referral threshold were more prevalent than upward adjustment, resulting in a net reclassification of -8.6% (-9.48% to -7.67%) **(Fig. 5a)**. Of the patients who became ineligible, most had Gleason <7 tumors (n=300, 72%; **Supplementary Table 16)**. In men of African ancestry, there were few changes in biopsy eligibility among p (n=392), with 3.1% reclassified upward and 4.6% downward **(Fig. 5b; Supplementary Table 16)**. Of 108 biopsy-negative controls, 75 (69.4%) were reclassified below the referral threshold based on PSA^G^, reflecting high enrichment for predisposition to PSA elevation (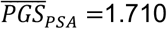). The overall net reclassification was positive, suggesting that PSA^G^ has some clinical utility in both populations.

### PSA Genetic Adjustment Improves Prostate Cancer Detection

The utility of PSA^G^, alone and in combination with PGS_269_, was first assessed in PCPT, where end-of-study biopsies were performed in all participants, effectively eliminating potential misclassification of prostate cancer status. Among 335 cases and 5548 controls, PGS_PSA_ was not associated with prostate cancer incidence (pooled: OR per SD=1.01, *P*=0.83), confirming it captures genetic determinants of non-cancer PSA variation. The magnitude of association for genetically adjusted baseline PSA^G^ with prostate cancer (OR per unit increase in log(ng/mL) =1.90, 95% CI: 1.56-2.31) was slightly larger than for PSA (OR=1.88, 1.55-2.29) in the European ancestry group **(Supplementary Table 17)**. The magnitude of association with prostate cancer was larger for PGS_269_^adj^ (pooled and European: OR per SD=1.57, 1.40-1.76) than for PGS_269_ without bias correction (pooled: OR=1.52, 1.36-1.70; European: OR=1.53, 1.36-1.72) **(Supplementary Table 17)**. The model with PGS_269_^adj^ and PSA^G^ achieved the best classification in the pooled (AUC=0.686) and European ancestry (AUC=0.688) populations, and outperformed PGS_269_^adj^ alone (pooled: AUC=0.656, *P*_AUC_=7.5×10^-4^; European: AUC=0.658, *P*_AUC_=1.4×10^-3^).

The benefit of genetically adjusting PSA was most evident for detection of aggressive prostate cancer, defined as Gleason ≥7, PSA ≥ 10 ng/mL, T3-T4 stage, and/or distant or nodal metastases. In PCPT, PSA^G^ conferred an approximately 3-fold risk increase (pooled: OR=2.87, 1.98-4.65, AUC=0.706; European: OR=2.99, 1.95-4.59, AUC=0.711) compared to PGS_269_^adj^ (pooled: OR=1.55, 1.23-1.95, AUC=0.651; European: OR=1.55, 1.22-1.96, AUC=0.657; **Fig. 6a; Supplementary Table 18)**. The model with PSA^G^ and PGS_269_^adj^ achieved AUC=0.726 (European: AUC=0.734) for aggressive tumors, but had lower discrimination for non-aggressive disease (pooled and European: AUC=0.681; **Supplementary Table 19)**. Among patients with prostate cancer, PSA^G^ (pooled: OR=2.06, 1.23-3.45) and baseline PSA (pooled: OR=1.81, 1.12-3.10) were associated with higher likelihood of aggressive compared to non-aggressive tumors, whereas PGS_269_ (pooled: OR=0.91, *P*=0.54) and PGS_269_^adj^ (OR=0.97, *P*=0.85) were not **(Supplementary Table 20)**.

**Fig. 6:**
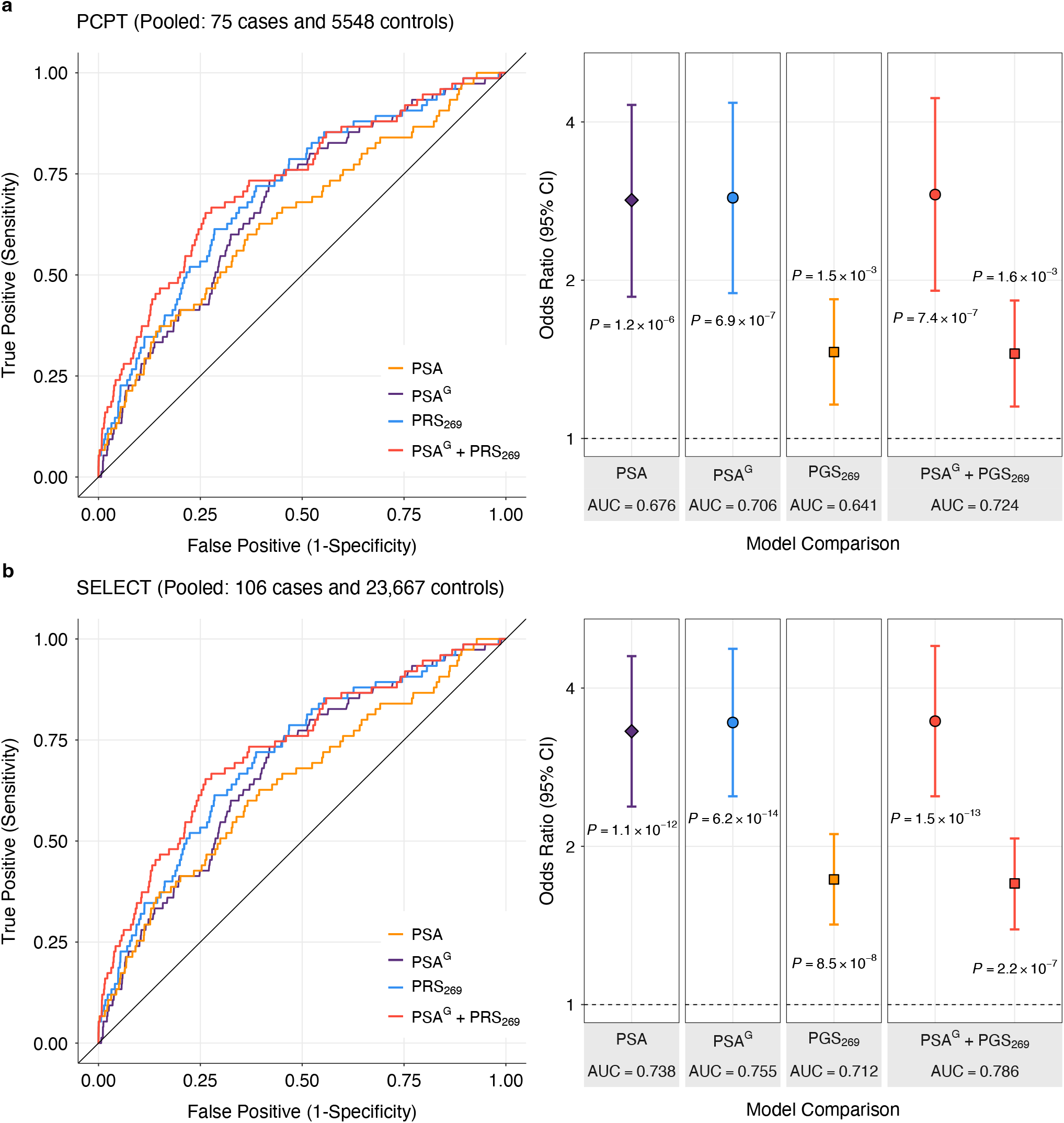
Genetic associations with aggressive prostate cancer. Comparison of models for aggressive disease, defined as Gleason score ≥7, PSA ≥ 10 ng/mL, T3-T4 stage, and/or distant or nodal metastases in the **a**, the Prostate Cancer Prevention Trial (PCPT) and **b**, Selenium and Vitamin E Cancer Prevention Trial (SELECT). The pooled study population includes all ancestry groups. Logistic regression models adjusted for baseline age, randomization arm, the top 10 population-specific genetic ancestry principal components, and proportions of African and East Asian genetic ancestry. Odds ratios (OR) and 95% confidence intervals were estimated per 1-unit increase in log(PSA ng/mL) and log(PSA^G^ ng/mL), and per standard deviation (SD) increase in the prostate cancer genetic risk score (PGS_269_) from Conti et al.^32^, which was standardized to achieve SD equal to 1. All p-values are two-sided. AUC is based on the full model with all covariates.

In SELECT, associations with risk of prostate cancer overall **(Supplementary Table 21)**, aggressive **(Fig. 6b; Supplementary Table 22)**, and non-aggressive disease **(Supplementary Table 23)** in the pooled and European ancestry analyses were comparable to PCPT. In men of East Asian ancestry, associations for PSA^G^ (OR=2.15, 0.82-5.62) were attenuated compared to PSA (OR=2.60, 1.03-6.54). This was also observed in men of African ancestry, although the effect size for PSA^G^ derived using PGS_128_ (OR=3.37, 2.38-4.78) was larger than for PSA^G^ based on PGS_CSx_ (OR=2.68, 1.94-3.69), consistent with the larger proportion of variation in PSA explained by PGS_128_ than PGS_CSx_ in this population. Models for prostate cancer including PSA^G^ were calibrated in the pooled and European ancestry individuals, while in the African ancestry subgroup, PSA^G^ inaccurately estimated risk in upper deciles **(Supplementary Fig. 3-6)**.

The largest improvement in discrimination from PSA^G^ (OR=3.81, 2.62-5.54; AUC=0.777) relative to PSA (OR=3.40, 2.34-4.93; AUC=0.742, *P*_AUC_=0.026) and to PGS_269_ (OR=1.76, 1.41-2.21; AUC=0.726; *P*_AUC_=0.057)_269_was for aggressive tumors in men of European ancestry (106 cases, 23,667 controls). In the pooled African ancestry population (18 cases, 2733 controls), PSA^G^ based on PGS_128_ (OR=2.96, 1.43-6.12), but not PGS_CSx_ (OR=2.48, 1.24-4.97), was more predictive than unadjusted PSA (OR=2.82, 1.33-5.99; **Supplementary Table 22)**. The best model for aggressive disease included PSA^G^ and PGS ^adj^ for pooled (AUC=0.788, 95% CI: 0.744-0.831) and European ancestry populations (AUC=0.804, 0.757-0.851), but for African ancestry individuals, unadjusted PSA and PGS_269_ without bias correction achieved the highest AUC of 0.828 (95% CI: 0.739-0.916). PSA^G^ was better calibrated than PSA in pooled and European ancestry groups, but not in African ancestry participants **(Supplementary Fig. 7-8)**.

## DISCUSSION

Serum PSA is the most widely used biomarker for prostate cancer detection, although concerns with specificity, and to a lesser degree sensitivity, have limited adoption of PSA testing for population-level screening. Leveraging PGS to personalize diagnostic biomarkers, such as PSA, provides a new avenue for translating GWAS discoveries into clinical practice. This concept, “de-Mendelization,” is essentially Mendelian randomization in reverse – subtracting the genetically-predicted component of trait variance instead of using it to estimate causal effects. De-Mendelization of non-causal predictive biomarkers can maximize disease-related signal and improve disease detection^38,39^. While previous work on PSA genetics^19^ and other biomarkers^38,40^ has alluded to the potential of genetic adjustment to produce clinically meaningful shifts in the PSA distribution, the value of this approach for reducing overdiagnosis and detecting aggressive disease has not been previously shown.

Risk stratified, personalized screening for prostate cancer will require parallel efforts to elucidate the genetic architecture of prostate cancer susceptibility and PSA variation in individuals without disease. Our GWAS advances these efforts by discovering 82 novel PSA-associated variants. The strongest novel signals map to genes involved in reproductive processes, potentially reflecting non-cancer function of PSA in liquefying seminal fluid. *TEX11* on Xq13.1 is preferentially expressed in male germ cells and early spermatocytes. *TEX11* mutations cause meiotic arrest and azoospermia, and this gene regulates homologous chromosome synapsis and double-strand DNA break repair^41^. *ODF3* encodes a component of sperm flagella fibers and has been linked to regulation of platelet count and volume^42^. Other novel loci contained genes involved in embryonic development, epigenetic regulation, and chromatin organization, including *DNMT3A, OTX1, CHD3, JARID2, HMGA1, HMGA2*, and *SUDS3. DNMT3A* is a methyltransferase that regulates imprinting and X-chromosome inactivation, and has been studied extensively in the context of height^43^, clonal hematopoiesis, and hematologic cancers^44^. *CHD3* is involved in chromatin remodeling during development and suppresses herpes simplex virus infection^45^. Multiple PSA-associated variants were in genes related to infection and immunity, including *HLA-A*; *ST6GAL1*, involved in IgG N-glycosylation^46^; *KLRG1*, which regulates NK cell function and IFN-γ production^47^; and *FUT2*, which affects ABO precursor H antigen presentation and confers susceptibility to viral and bacterial infections^48^.

Although our GWAS was restricted to men without prostate cancer, several cancer susceptibility genes were among the PSA-associated loci, including a pan-cancer risk variant in *TP53* (rs78378222)^49^ and signals in *TP63, GPC3*, and *THADA*. While we cannot rule out undiagnosed prostate cancer in our participants, its prevalence is unlikely to be high enough to produce appreciable bias. Pervasive pleiotropy and omnigenic architecture^50^ may explain the diverse functions of PSA loci implicated in inflammation, epigenetic regulation, and growth factor signaling. Even established tumor suppressor genes, like *TP53, GPC3*, and *THADA* have pleiotropic effects on obesity via dysregulation of cell growth and metabolism^51-53^. Furthermore, distinct p63 isoforms regulate epithelial and craniofacial development, as well as apoptosis of male germ cells and spermatogenesis^54,55^. Mutations in *GPC3* cause Simson-Golabi-Behmel syndrome, which is characterized by visceral and skeletal abnormalities and excess risk of embryonic tumors^56^.

Distinguishing variants that influence prostate cancer detection via PSA screening from genetic signals for prostate carcinogenesis has implications for deciphering biological mechanisms and developing risk prediction models. Prostate cancer detection depends on PSA testing, while PSA screening is influenced by genetic factors affecting constitutive PSA levels. The bias arising from this complex relationship may be substantial. Our findings suggest that bias-corrected effect sizes more accurately capture the contribution of GWAS-identified variants to prostate cancer risk, without conflating it with detection. Correction for PSA-related bias and subsequent improvement in PGS_269_ performance for detecting aggressive disease is an extension of de-Mendelization. Adjusting risk allele weights may be a more effective strategy than filtering out variants based on associations with PSA. Generally, the improvements in PSA^G^ and PGS_269_ are proportional to the extent of their de-noising of signals for PSA elevation unrelated to prostate cancer. The impact of bias correction was most pronounced in populations selected for high PSA, such as men who underwent prostate biopsy in GERA, but it was also observed in PCPT and SELECT, which enrolled men with low PSA.

Our investigation of index event bias has several limitations. The Dudbridge method assumes that direct genetic effects on PSA levels and prostate cancer susceptibility are uncorrelated, and violations of this assumption over-attribute shared genetic signals to selection bias^33^. Although SlopeHunter relaxes this assumption^35^, analyses of PGS_269_ suggest it under-corrects selection bias. SlopeHunter relies on clustering to distinguish PSA-specific from pleiotropic variants^35^, with small or poorly separated clusters resulting in unstable bias estimates. Disentangling genetic associations between PSA and prostate cancer with greater certainty will require experiments, such as CRISPR screens and massively parallel reporter assays.

Another limitation is that the reported magnitude of biopsy reclassification may be specific to GERA and Kaiser Permanente clinical guidelines and biased since GERA controls comprised 30% of the PSA discovery GWAS. Since it was unlikely for men with low PSA to be biopsied and most patients with prostate cancer already had PSA values at or above the biopsy referral cutoff, there were limited opportunities to increase biopsy eligibility in this population. Despite these limitations, our findings indicate that genetically adjusted PSA may reduce overdiagnosis and overtreatment, albeit accompanied by some undesirable loss of sensitivity. While reclassifying pro to not receive biopsy is concerning, most such reclassifications occurred among patients with non-aggressive disease, a group susceptible to overdiagnosis^57^.

Our PGS-based approach updates the first application of PSA genetic correction by Gudmundsson et al.^19^ while retaining straightforward calculation of the genetic correction factor. Increasing the specificity of an established, clinically useful biomarker is efficient and would have low adoption barriers. However, analytic choices, such as selecting an optimal PGS algorithm and reference population for obtaining mean PGS_PSA_, are not trivial. The choice of reference population affects the magnitude of correction and clinical decisions based on absolute PSA values. Furthermore, any new biomarker would require validation in real-world settings to identify populations who would benefit most and characterize barriers to implementation, such as physician familiarity with PGS and patient education about genetic testing. Genetically adjusted PSA should also be evaluated in conjunction with other procedures used for prostate cancer detection, such as targeted MRI, and explored as a criterion for refining selection of participants into screening trials.

Our study highlights the importance and challenge of developing a PGS that adequately performs across the spectrum of ancestry. Compared to PGS_128_, PGS_CSx_ did not improve performance in men of African ancestry. This may reflect the ‘meta’ estimation procedure, which does not require a separate dataset for hyperparameter tuning, but is less accurate^36^. GWAS efforts in larger and more diverse cohorts are under way and will expand the catalog of PSA-associated variants and increase their utility. Genetic adjustment using a PGS_PSA_ that does not explain a sufficiently high proportion of trait variation risks decreasing the accuracy of PSA screening.

Future research should assess whether genetically adjusted PSA levels improve prediction of prostate cancer mortality and investigate PSA-related biomarkers, such as the ratio of free to total PSA and pro-PSA, which may have higher specificity for prostate cancer detection^58,59^. Although PGS_PSA_ was associated with PSA doubling time and velocity, these metrics assess change between two time points and may not capture PSA trajectories meaningful for disease detection^60^. Clinical guidelines for PSA kinetics are also lacking in the context of prostate cancer screening. Regardless, we believe that genetic adjustment may improve the accuracy of any heritable PSA biomarker and may be a valuable addition to multi-omic biomarkers.

In summary, by detecting genetic variants associated with non-prostate cancer PSA variation, we developed a PGS_PSA_ that captures the contribution of common genetic variants to a man’s inherent PSA level. We showed that a straightforward calculation of genetically adjusted, personalized PSA levels using PGS_PSA_ provides clinically meaningful improvements in prostate cancer diagnostic characteristics. Moreover, genetic determinants of PSA provide an avenue for mitigating selection bias due to PSA screening in prostate cancer GWAS and improving disease prediction. These results illustrate a proof of concept for incorporating genetic factors into PSA screening for prostate cancer and expanding this approach to other diagnostic biomarkers.

## Data Availability

UK Biobank data are publicly available by request from https://www.ukbiobank.ac.uk. To maintain individuals' privacy, data on the GERA cohort are available by application to the Kaiser Permanente Research Bank (researchbank.kaiserpermanente.org).

## ACKNOWLEDGEMENTS

The Precision PSA study is supported by funding from the National Institutes of Health (NIH) National Cancer Institute (NCI) under award number R01CA241410 (JSW) and K99CA246076 (LK) and the Young Investigator Award from the Prostate Cancer Foundation (REG). Contributing studies were supported by research grants from the NIH National Institute of General Medical Sciences (NIGMS) under award number R01GM130791 (JDM); NIH/NCI Cancer Center Support Grant to Memorial Sloan Kettering Cancer Center (P30CA008748), MSKCC Specialized Programs of Research Excellence in Prostate Cancer (P50CA92629, HL), Swedish Cancer Society (Cancerfonden 20 1354 PjF, HL), and General Hospital in Malmö Foundation for Combating Cancer. This work was supported in part through the computational resources and staff expertise provided by Scientific Computing at the Icahn School of Medicine at Mount Sinai. Research reported in this paper was supported by the Office of Research Infrastructure of the National Institutes of Health under award number S10OD026880 and NIH/NCI funding (R01CA175491, R01CA244948; RJK).

The content is solely the responsibility of the authors and does not necessarily represent the official views of the National Institutes of Health.

## AUTHOR CONTRIBUTIONS

Concept and design: L.K, T.J.H., R.J.K., J.D.M., R.E.G., J.S.W. Acquisition, analysis or interpretation of data: L.K, T.J.H., Y.J., S.I.B., J.P.S, K.S., M.J.M, N.D.F., W.-Y.H., S.A.L, R.E., P.J.G., C.T., I.T., H.L, S. K.V.D.E, S.J.C., C.A.H., D.V.C., R.J.K., J.D.M., R.E.G., J.S.W. Drafting of the manuscript: L.K, T.J.H., R.E.G., J.S.W.

Critical revision of the manuscript for important intellectual content: L.K, T.J.H., Y.J., S.I.B., J.P.S, K.S., M.J.M, N.D.F., W.-Y.H., S.A.L, R.E., P.J.G., C.T., I.T., H.L, S. K.V.D.E, S.J.C., C.A.H., D.V.C., R.J.K., J.D.M., R.E.G., J.S.W.

## COMPETING INTERESTS

JSW is a non-employee, cofounder of Avail Bio. HL is named on a patent for intact PSA assays and a patent for a statistical method to detect prostate cancer that is licensed to and commercialized by OPKO Health. HL receives royalties from sales of the test and has stock in OPKO Health. All other authors have no competing interests.

## METHODS

Informed consent was obtained from all study participants. UK Biobank received ethics approval from the Research Ethics Committee (REC reference: 11/NW/0382) in accordance with the UK Biobank Ethics and Governance Framework. The research was conducted with approved access to UK Biobank data under application number 14105. The Vanderbilt Institutional Review Board approved the BioVU study. We used previously published PSA GWAS results from the GERA cohort by Hoffmann et al^17^. The original study was approved by the Kaiser Permanente Northern California Institutional Review Board and the University of California San Francisco Human Research Protection Program Committee on Human Research. The Malmö Diet and Cancer Study (MDCS) was approved by the local ethics committee. The PLCO study was approved by the institutional review board at each participating centre and the National Cancer Institute. The informed consent document signed by the PLCO study participants allows use of these data by investigators for discovery and hypothesis generation in the investigation of the genetic contributions to cancer and other adult diseases. Our study includes publicly posted genomic summary results from PLCO Atlas^61^. No IRB review is required for PLCO summary data use.

### Study Populations and Phenotyping

Genome-wide association analyses of PSA levels were conducted using germline genetic data derived from DNA extracted from non-prostatic tissues (e.g., blood, buccal swabs). Analyses were restricted to cis-gender men, defined as individuals of biological male sex and self-reported male gender identity, who have never been diagnosed with prostate cancer. Men with a history of surgical resections of the prostate were excluded in studies for which this information was available. To reduce potential for reverse causation, analyses were limited to PSA values ≤10 ng/mL, which corresponds to low-risk prostate cancer based on the D’Amico prostate cancer risk classification system^62^, and PSA>0.01 ng/mL, to ensure that individuals had a functional prostate not impacted by surgery or radiation.

The UK Biobank (UKB) is a population-based prospective cohort of over 500,000 individuals aged 40-69 years at enrollment in 2006-2010 with genetic and phenotypic data^63^. Health-related outcomes were ascertained via individual record linkage to national cancer and mortality registries and hospital in-patient encounters. PSA values were abstracted from primary care records for a subset of participants linked to genetic data. Field code mappings used to identify PSA values included any serum PSA measure except for free PSA or ratio of free to total PSA **(Supplementary Table 25)**.

The Kaiser Permanente Genetic Epidemiology Research on Adult Health and Aging (GERA) cohort used in this analysis has been previously described in Hoffmann et al^17^. Briefly, prostate cancer status was ascertained from the Kaiser Permanente Northern California Cancer Registry, the Kaiser Permanente Southern California Cancer Registry, or through review of clinical electronic health records. PSA levels were abstracted from Kaiser Permanente electronic health records from 1981 through 2015.

The Prostate, Lung, Colorectal and Ovarian (PLCO) Cancer Screening Trial is a completed randomized trial that enrolled approximately 155,000 participants between November 1993 and July 2001. PLCO was designed to determine the effects of screening on cancer-related mortality and secondary endpoints in men and women aged 55 to 74^64^. Men randomized to the screening arm of the trial underwent annual screening with PSA for six years and digital rectal exam (DRE) for four years^64^. These analyses were limited to men with a baseline PSA measurement who were randomized to the screening arm of the trial (N=29,524). Men taking finasteride at the time of PSA measurement were excluded from analysis.

The Vanderbilt University Medical Center BioVU resource is a synthetic derivative biobank linked to deidentified electronic health records^65^. Analyses were based on PSA levels that were measured as part of routine clinical care.

The Malmö Diet and Cancer Study (MDCS) is a population-based prospective cohort study that recruited men and women aged between 44 and 74 years old who were living in Malmö, Sweden between 1991 and 1996 to investigate the impact of diet on cancer risk and mortality^66^. These analyses included men from the MDCS who were not diagnosed with prostate cancer as of December 2014 and had available genotyping and baseline PSA measurements^66^.

PCPT is a completed phase III randomized, double-blind, placebo-controlled trial of finasteride for prostate cancer prevention that began in 1993^8^. PCPT randomly assigned 18,880 men aged 55 years or older who had a normal DRE and PSA level ≤3 ng/mL to either finasteride or placebo. For men with multiple prerandomization PSA values, the earliest value was selected. Cases included all histologically confirmed prostate cancers detected during the 7-year treatment period and tumors that were detected by the end-of-study prostate biopsy. Our analyses included the subset of PCPT participants that was genotyped on the Illumina Infinium Global Screening Array (GSAMD) 24v2-0 array.

SELECT is a completed phase III randomized, placebo-controlled trial of selenium (200 μg/day from L-selenomethionine) and/or vitamin E (400 IU/day of all rac-α-tocopheryl acetate) supplementation for prostate cancer prevention^37^. Between 2001 and 2004, 34,888 eligible participants were randomized. The minimum enrollment age was 50 years for African American men and 55 years for all other men^37^. Additional eligibility requirements included no prior prostate cancer diagnosis, ≤4 ng/mL of PSA in serum, and a DRE not suspicious for cancer. For men who had multiple pre-randomization PSA values, the earliest value was selected. Our analyses included a subset of SELECT participants genotyped on the Illumina Infinium Global Screening Array (GSAMD) 24v2-0 array.

### Quality Control and Genome-Wide Association Analyses

Standard genotyping and quality control (QC) procedures were implemented in each participating study. Prior to meta-analysis we applied variant-level QC filters that included low imputation quality (INFO<0.30), MAF<0.005, deviations from Hardy-Weinberg equilibrium (*P*_HWE_<1×10^-5^). Sample-level QC filtered based on discordant genetic sex and self-reported gender, call rate ≥97%, and removed one sample from each pair of first-degree relatives. GWAS phenotypes and adjustment covariates are reported in **Supplementary Table 26**. Genome-wide association analyses performed linear regression of log(PSA) as the outcome, using age and genetic ancestry principal components (PC’s) as the minimum set of covariates.

#### UK Biobank

Genotyping and imputation for the UK Biobank cohort have been previously described^63^. Briefly, participants were genotyped on the UKB Affymetrix Axiom array (89%) or the UK BiLEVE array (11%) with imputation performed using the Haplotype Reference Consortium (HRC) and the merged UK10K and 1000 Genomes (1000G) phase 3 reference panels. Genetic ancestry principal components (PCs) were computed using fastPCA based on a set of 407,219 unrelated samples and 147,604 genetic markers^63^. Association analyses in UKB were restricted to individuals of European ancestry based on self-report (“White”) and after excluding samples with any of the first two genetic ancestry PC’s outside of 5 standard deviations (SD) of the population mean, as previously descibed^49^. We removed samples with discordant self-reported and genetic sex, as well as one sample from each pair of first-degree relatives identified using KING^67^. Using a subset of genotyped autosomal variants with (MAF)≥0.01 and call rate ≥97%, we filtered samples with heterozygosity >5 SD from the mean. For participants with multiple PSA measurements, the median value PSA was used. Sensitivity analyses were conducted comparing this approach to a GWAS of individual-specific random effects derived from fitting a linear mixed model to repeated log(PSA) values.

#### Genetic Epidemiology Research on Adult Health and Aging (GERA)

Genotyping, imputation, and QC of the GERA cohort has been previously described^17,68,69^. Briefly, all men were genotyped for over 650,00 SNPs on four race/ethnicity-specific Affymetrix Axiom arrays that were optimized for individuals who self-identified as non-Hispanic white, Latino, East Asian, and African American, respectively^68,69^. Genotype QC procedures and imputation for the original GERA cohort were performed on an array-wise basis, as previously described^17,70^. Pre-phasing was done by SHAPEIT v2.5^71^ and imputation with IMPUTE2 v2.3.1^72^ using the 1000 Genomes Phase 3 release with 2504 samples. The top 10 genetic ancestry PC’s from Eigenstrat v4.2, were included in the linear model as ancestry covariates^73^. Analyses were conducted according to self-identified race/ethnicity groups. Residuals were computed from linear mixed models that were fit to repeated log(PSA) measures. This approach nearly identical to a long-term average, except it uses the median instead of the mean to handle any potential outlier PSA level values.

#### Prostate, Lung, Colorectal and Ovarian (PLCO) Atlas

Our study used GWAS summary statistics from the PLCO Atlas Project, a resource for multi-trait GWAS. Genotyping, QC, and imputation procedures for this resource are described by Machiela et al.^61^ The Atlas project combined genotyping data previously generated by high density arrays for 25,831 participants (OncoArray, Omni2.5 M, and OmniExpress) with a new round of genotyping using the Illumina Global Screening Array (GSA). For participants genotyped on multiple genotyping arrays (N = 1,192), data from only one array were retained, with the following prioritization: GSA > OncoArray > Omni2.5 M > OmniExpress (OmniX). Extensive quality control filtering was performed for subsequent imputation and association analyses. Iterative 80% and 95% sample- and variant-level call rate filters were applied to remove poorly genotyped samples and variants. Samples with greater than 20% estimated contamination based on VerifyIDintensity^74^ were also removed. Samples with discordant self-reported gender and genetically inferred sex were identified based on X chromosome method-of-moments F coefficient from PLINK, using 0.5 as the threshold (F coefficients are close to 0.0 for males and 1.0 for females). Heterozygosity outliers were detected using absolute values from PLINK method-of-moments F coefficients >0.2.

Genetic ancestry was determined using GRAF^75^ on a set of 10,000 pre-selected fingerprinting variants. Participants were assigned to 9 ancestral groups: “African”, “African American”, “East Asian”, “European”, “Hispanic1”, “Hispanic2”, “Other”, “Other Asian”, and “South Asian”. Hispanic1 included individuals of Dominican or Puerto Rican ancestry whereas Hispanic2 included individuals of Mexican or Latin American ancestry. For parsimony we merged “African” and “African American” into a “African American (Combined)” and “East Asian” and “Other Asian” into a “East Asian (Combined)”. Imputation was performed using the TOPMed 5b reference panel, which is accessible via the TOPMed Imputation Server hosted on the Michigan Imputation Server. Prior to imputation, variants with MAF≤0.01, missingness ≥0.05, and Hardy Weinberg deviations (*P*_HWE_β1×10^-6^) were removed. Genotyped data were aligned to reference datasets using a community-recommended script (HRC-1000G-check-bim.pl from https://www.well.ox.ac.uk/~wrayner/tools/) that was modified to support the TOPMed 5b reference panel using a pre-existing test imputation with 1000 Genomes subjects. Pre-phasing using phased reference data from TOPMed release 5b was conducted using EAGLE 2.4^76^. Imputation was conducted against the same reference panel using minimac4. GWAS was based on the first PSA value for each PLCO participant.

#### BioVU

Participants were identified using Vanderbilt University Medical Center’s (VUMC) BioVU resource, a DNA biobank comprising ∼270,000 individuals and linked to a de-identified electronic health record (EHR)^65^. All participants (n=8,074) were genotyped on Illumina’s Expanded Multi-Ethnic Genotyping Array (MEGA^EX^) platform. Genetic ancestries were assigned by running principal component analysis using SNPRelate^77^ on a set of pruned SNPs (Rsq<0.5, MAF≥0.1). Subjects were classified as being of European ancestry if their first two PCs were within 4 SDs of the median for the subjects reporting “White” as their race. Subjects were classified as being of African ancestry if their first two PCs were within 4 SDs of the median for subjects reporting their race as “Black”. All quality control procedures were performed using PLINK version 1.90. We removed one randomly selected sample out of each pair of related individuals (pi-hat≥0.2) identified using identity-by-descent. We excluded subjects with SNP missingness >3% or heterozygosity >5 SD from the mean. Prior to imputation, data were pre-processed using the HRC-1000G-check-bim.pl (from http://www.well.ox.ac.uk/~wrayner/tools/) and pre-phased using Eagle v2.4^76^. Genetic data was imputed on the Michigan Imputation Server using 1000 Genomes phase 3 version 5 as the reference panel. For men with multiple PSA measurements, the median PSA was used.

#### Malmö Diet and Cancer Study (MDCS)

Data from multiple batches of genotyping of 4069 MDCS participants using different Illumina Omni arrays were merged. For variants that appeared more than once under different names on the same Illumina array, those with the higher genotyping rate were retained. Indels, ambiguous palindromic (eg. A/T or C/G alleles) and multi-allelic variants were removed. Only SNPs that we could unambiguously map to 1000 Genomes phase 1 dataset were kept. Individuals with more than 10% missingness were removed. Next, SNPs with a missingness rate greater than 10% or deviation from Hardy-Weinberg equilibrium (*P*_HWE_<0.001) were removed. At this stage, the principal components of ancestry were computed. Individuals for whom the inferred sex based on X chromosome heterozygosity was not male, or for whom there were more than two genetic mismatches with 40 SNPs we had previously genotyped in these samples with targeted genotyping^66^ were excluded.

To assess genetic ancestry, data were combined with data from HapMap phase 3 for variants present in all genotyping batches. These SNPs were further filtered to have less than 0.01% missingness and LD-pruned (--indep-parwise 50 5 0.05). SMARTPCA in EIGENSOFT (https://github.com/chrchang/eigensoft) was run on the resulting 18,299 SNPs to generate the top 10 genetic ancestry PC’s. Analyses were restricted to individuals of European ancestry based on clustering with HapMap reference populations and exclusion of outliers with a Z-score on PC1 and PC2 greater than 5. Imputation was performed using the TOPMed 5b reference panel, which is accessible via the TOPMed Imputation Server hosted on the Michigan Imputation Server. Prior to imputation, the input file was aligned to the build37 reference genome on the basis of chromosome, position and alleles. A total of 847,133 SNPs that passed pre-imputation QC were uploaded to the imputation server. From the resulting imputed files, analyses were restricted to individuals without a prostate cancer diagnosis by December 31, 2014, with individual missingness less than 3%, a Z-score less than 5.0 for heterozygosity. Log(PSA) values were analyzed using robust linear regression with Tukey biweights. GWAS was performed using linear regression on the residuals were extracted from the fitted models.

#### Prostate Cancer Prevention Trial (PCPT) and Selenium and Vitamin E Cancer Prevention Trial (SELECT)

Participants from PCPT and SELECT were genotyped on the Illumina Infinium Global Screening Array (GSAMD) 24v2-0 array and underwent the same QC and imputation procedures. Genotyping calling and quality control were performed at the Center for Inherited Disease Research (CIDR) at Johns Hopkins After removal of samples that failed to produce valid output during initial processing and clustering, the completion rate was 0.9951 and 0.9959 in PCPT and SELECT, respectively. A two-stage filter by completion rate threshold of 0.8 for samples and 0.8 for variants, followed by 0.95 for samples and 0.95 for variants was performed. Samples with discordant self-reported gender and genetically inferred sex were identified based on X chromosome method-of-moments F coefficient from PLINK, using 0.5 as the threshold (F coefficients are close to 0.0 for males and 1.0 for females). Identity-by-descent (IBD) for all subject pairs were using PLINK and close (1^st^ and 2^nd^ degree) relatives were identified based on a threshold of 0.20. One randomly selected sample from pair of relatives was retained.

Ancestry was estimated using a set of LD-pruned markers and running SNPWEIGHTS^78^ with the reference panel provided containing the following populations: European, West African, and East Asian, with a threshold of 0.8 used for imputed ancestry designation. Subjects were assigned to a single ancestry group if the ancestry score was equal to or above 0.80 for just one group. Individuals were assigned to an admixed cluster if their ancestry score was >0.20 and <0.80 for only one group (eg: ADMIXED_AFR where AFR=0.75, EUR=0.17, EAS=8). Intermediate ancestry clusters included individuals with ancestry scores matching those criteria in multiple groups: 0.20<AFR_EUR<0.80 (eg: AFR=0.65, EUR=0.33) and 0.20<EAS_EUR<0.80 (eg: EUR=0.55, EAS=0.43). Autosomal heterozygosity was assessed using the method-of-moments F coefficient calculated within each ancestry cluster. Heterozygosity outliers were identified and excluded using a threshold of 0.10. Principal components analysis was performed with SMARTPCA in EIGENSOFT (https://github.com/chrchang/eigensoft) on a set of LD-pruned markers after splitting by ancestry cluster, to resolve more detailed population substructure. Genetic ancestry PC’s were not computed for small clusters (n<50) or individuals who failed other QC filters. For validation of PGS_PSA_ in PCPT and SELECT we combined ADMIXED_AFR and AFR_EUR and treated this as a single group with admixed AFR and EUR ancestry proportions (AFR/EUR). ADMIXED_EAS and EAS_EUR were also combined into a single cluster with admixed EAS and EUR ancestry (EAS/EUR).

To prepare genotype data for imputation with the TOPMed 5b reference panel, variants with MAF<0.001, call rate <98%, or evidence of deviation from Hardy-Weinberg equilibrium (*P*_HWE_<10^-6^) were removed. After these QC step, a total of 474,046 variants remained for PCPT and 491,015 variants were retained for SELECT. Prior to submitting the data to the TOPMed Imputation Server, files were pre-processed using the check-bim.pl script (http://www.well.ox.ac.uk/~wrayner/tools/). Next, chromosomal positions were lifted over from GRCh37/hg19 to GRCh38 and aligned against the TOPMed reference SNP list based on chromosome, position, and alleles to ensure that reference and alternate alleles were correct in the resulting VCF files.

### Heritability of PSA Levels Attributed to Common Variants

Heritability of PSA levels was estimated using individual-level data and GWAS summary statistics. UKB participants with available PSA and genetic data were analyzed using Linkage Disequilibrium Adjusted Kinships (LDAK) v5.1^24^ and GCTA v1.93^23^, following the approach previously implemented in the GERA cohort^17^. Genetic relationship matrices were filtered to ensure that no pairwise relationships with kinship estimates>0.05 remained. Heritability was estimated using common (MAF≥0.01) LD-pruned (*r*^2^<0.80) variants with imputation INFO>0.80. We implemented the LDAK-Thin model using the recommended GRM settings (INFO>0.95, LD *r*^2^<0.98 within 100 kb) and the same parameters as GCTA for comparison (LD *r*^2^<0.80, INFO>0.80). For both methods, sensitivity analyses were conducted using more stringent GRM settings (kinship=0.025, genotyped variants).

Summary statistics from GWAS results based on the same set of UKB participants (n=26,491) and from a European ancestry GWAS meta-analysis (n=85,824) were analyzed using LDAK, LD score regression (LDSR)^25^, and an extension of LDSR using a high-definition likelihood (HDL) approach^26^. For LDSR we used the default panel comprised of variants available in HapMap3 with weights computed in 1000 Genomes v3 EUR individuals and in-house LD scores computed in UKB European ancestry participants^49^. The baseline linkage disequilibrium (BLD)-LDAK model was fit using precomputed tagging files calculated in UKB GBR (white British) individuals for HapMap3 variants from the LDSR default panel. HDL analyses were conducted using the UKB-derived panel restricted to high-quality imputed HapMap3 variants^26^. All GWAS summary statistics had sufficient overlap with the reference panels, not exceeding the 1% missingness threshold for HDL and 5% missingness threshold for LDAK and LDSR.

### Genome-Wide Meta-Analysis

Each ancestral population was analyzed separately, and GWAS summary statistics were combined via metaanalysis **(Figure 1)**. We first used METAL^79^ to conduct an inverse-variance-weighted fixed-effects metaanalysis in each ancestry group and then meta-analyzed the ancestry-stratified results. Multi-ancestry metaanalysis results were processed using clumping to identify independent association signals by grouping variants based on linkage disequilibrium within specific windows. Clumps were formed around index variants with the lowest genome-wide significant (*P*<5×10^-8^) meta-analysis p-value. All other variants with LD *r*^2^ >0.01 within a ± 10Mb window were considered non-independent and assigned to that lead variant. Since over 90% of the meta-analysis consisted of individuals of European ancestry, clumping was performed using 1000 Genomes (1000G) Phase 3 EUR and UKB reference panels, which yielded concordant results. We confirmed that LD among the resulting lead variants did not exceed *r*^2^=0.05 using a merged 1000G ALL reference panel.

We first examined heterogeneity in the multi-ancestry fixed-effects meta-analysis results using Cochran’s Q statistic. To assess heterogeneity specifically due to ancestry we applied MR-MEGA^27^, a meta-regression approach for aggregating GWAS results across diverse populations. Summary statistics from each GWAS were meta-analyzed using MR-MEGA without combining by ancestry first. The MR-MEGA analysis was performed across four axes of genetic variation derived from pairwise allele frequency differences, based on the recommendation for separating major global ancestry groups. Index variants from the MR-MEGA analysis were selected using the same clumping parameters as described above (LD *r*^2^ <0.01 within ± 10Mb window), based on the merged 1000G ALL reference panel. For each variant, we report two heterogeneity p-values: one that is correlated with ancestry and accounted for in the meta-regression (*P*_Het-Anc_) and the residual heterogeneity that is not due to population genetic differences (*P*_Het-Res_).

### PSA Genetic Score (PGS_PSA_) Development and Validation

We implemented two strategies for generating a genetic score for PSA levels. In the first approach, we selected 128 variants that were genome-wide significant (*P*<5×10^-8^) in the multi-ancestry meta-analysis and were independent (LD *r*^2^ <0.01 within ± 10Mb window) in 1000G EUR and (LD *r*^2^<0.05) 1000G ALL populations (PGS_128_). Each variant in PGS_128_ was weighted by the meta-analysis effect size estimated using METAL. As an alternative strategy to clumping and thresholding, we fit a genome-wide score using the PRS-CSx algorithm^36^, which takes GWAS summary statistics from each ancestry group as inputs and estimates posterior SNP effect sizes under coupled continuous shrinkage priors across populations (PGS_CSx_). Analyses were conducted using pre-computed population-specific LD reference panels from the UKB, which included 1,287,078 HapMap3 variants that are available in both the UKB and 1000 Genomes Phase 3.

We calculated a single trans-ancestry PGS that can be applied to all participants in the target cohort, rather than optimizing a PGS within each ancestry group. This approach is more robust to differences in genetic ancestry assignments across studies and does not require separate testing and validation datasets for parameter tuning each ancestry group ^36^. To facilitate this type of analysis, PRS-CSx provides a --meta option that integrates population-specific posterior SNP effects using an inverse-variance-weighted meta-analysis in the Gibbs sampler^36^. The global shrinkage parameter was set to f=0.0001. PRS-CSx was run on the intersection of variants that were in the LD reference panel and had imputation quality (INFO>0.90), resulting in 1,058,163 variants in PCPT and 1,071,268 variants in SELECT. Since PRS-CSx only considers autosomes, chrX variants that were included in PGS_128_ were added to PGS_CSx_ separately, when output files from each chromosome produced by the PLINK --score command were concatenated.

The predictive performance of PGS_CSx_ and PGS_128_ was evaluated in two independent cancer prevention trials that were not included in the meta-analysis: PCPT and SELECT. Analyses were conducted in the pooled sample for each cohort, which included individuals of all ancestries who passed quality control filters **(see Supplementary Note)**. Ancestry-stratified analyses were conducted for clusters with n>50 with available genetic ancestry PC’s. Ancestry scores were computed with SNPWEIGHTS^78^. Individuals with ancestry scores ≥0.80 for a single group were assigned to clusters for predominantly European (EUR), West African (AFR) and East Asian (EAS) ancestry. Admixed individuals with intermediate ancestry scores for at least one group were assigned to separate clusters: 0.20<EUR/AFR<0.80 or 0.20<EUR/EAS<0.80. Pooled analyses were adjusted for 10 within-cluster PC’s and global ancestry proportions (AFR, EAS).

### Index Event Bias Analysis

Index event bias occurs when individuals are selected based on the occurrence of an event or specific criterion. This is analogous to the direct dependence of one phenotype on another, as in the commonly used example of cancer survival^34^. Due to unmeasured confounding, this dependence can induce correlations between previously independent risk factors among those selected^33,34^. Genetic effects on prostate cancer can be viewed as conditional on PSA levels, since elevated PSA typically triggers diagnostic investigation. Genetic factors resulting in higher constitutive PSA levels may also increase the likelihood of prostate cancer detection due to more frequent testing **(Figure 4)**. This selection mechanism could bias prostate cancer

GWAS associations by capturing both direct genetic effects on disease risk and selection-induced PSA signals. In the GWAS setting, methods using summary statistics have been developed to estimate and correct for this bias^33,35^. Although typically derived assuming a binary selection trait, these methods are still applicable to selection or adjustment based on quantitative phenotypes^33^. In this study, we conceptualized PSA variation as the selection trait and prostate cancer incidence as the outcome trait **(Figure 4)**.

We applied the method described in Dudbridge et al.^33^, which tests for index event bias and estimates the corresponding correction factor (*b*) by regressing genetic effects on the selection trait (PSA) against their effects on the subsequent trait (prostate cancer), with inverse variance weights: *w* = 1/(*SE*_*prCa*_)^2^. Summary statistics for prostate cancer were obtained from the most recent prostate cancer GWAS from the PRACTICAL consortium^32^. Sensitivity analyses were performed using SlopeHunter^35^, an extension of the Dudbridge approach that allows for direct genetic effects on the index trait and subsequent trait to be correlated. For both methods, analyses were conducted using relevant summary statistics and 127,906 variants pruned at the recommended threshold^33^ (LD *r*^2^<0.10 in 250 kb windows) with MAF ≥0.05 in the 1000G EUR reference panel. After merging the pruned 1000G variants with each set of summary statistics, variants with large effects, (|β|>0.20) on either log(PSA) or prostate cancer, were excluded. The resulting estimate (*b*), adjusted regression dilution using the SIMEX algorithm, was used as a correction factor to recover unbiased genetic effects for each variant: 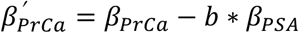, where β_*PSA*_ is the per-allele effect on log(PSA), and β_*prCa*_ is the log(OR) for prostate cancer.

The impact of the bias correction was assessed in three ways. First, genome-wide significant prostate cancer index variants were selected from the European ancestry PRACTICAL GWAS meta-analysis (85,554 cases and 91,972 controls) using clumping (LD *r*^2^<0.01 within 10 Mb)^32^. We tabulated the number of variants that remained associated at *P*<5×10^-8^ after bias correction. Next, we fit genetic scores for PSA and prostate cancer in men of European ancestry in UKB who were not included in the PSA or prostate cancer GWAS (11,568 prostate cancer cases and 152,884 controls). We compared the correlation between the PGS for PSA (PGS_PSA_), comprised of 128 lead variants, and the 269-variant prostate cancer risk score fit with original risk allele weights (PGS_269_) and with weights corrected for index event bias (PGS_269_^adj^). To allow adjustment for genetic ancestry PCs and genotyping array, associations between the two scores were estimated using linear regression models. Next, we examined associations for each genetic score (PGS_269_, PGS_269_^adj^, PGS_269_^adj-S^) with prostate cancer in a subset of GERA participants who underwent a biopsy. Since GERA controls were include in the PSA GWAS meta-analysis, AUC estimates and corresponding bootstrapped 95% confidence intervals were obtained using 10-fold cross-validation. We also examined PGS associations with Gleason score, a marker of disease aggressiveness, which was not available in the UK Biobank. Multinomial logistic regression models with Gleason score ≤6 (reference), 7, and ≥8 as the outcome were fit for each score in 4584 cases from the GERA cohort.

### Application of Genetically Adjusted PSA for Biopsy Referral and Prostate Cancer Detection

Genetically corrected PSA values were calculated for individual *i* as follows^17,19^:

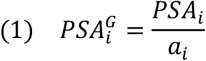

where *a*_*i*_ is a personalized adjustment factor derived from PGS_PSA_. Since genetic effects were estimated for log(PSA), *a*_*i*_ for correcting PSA in ng/mL was derived as:

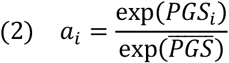

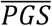 can be estimated in controls without prostate cancer or obtained from an external control population^17,19^. We see that *a*_*i*_ ***>*** 1 when an individual has a higher multiplicative increase in PSA than the sample average due to their genetic profile, resulting in a lower genetically adjusted PSA compared to the observed value 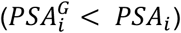.

We evaluated the potential utility of PGS_PSA_ in two clinical contexts. First, we quantified the impact of using 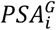 on biopsy referrals by examining reclassification at age-specific PSA thresholds used in the Kaiser Permanente health system. Analyses were conducted in GERA participants with information on biopsy date and outcome, comprised of prostate cancer cases not included in the PSA GWAS and controls that were part of the PSA GWAS. To use the same normalization factor for both cases and controls while mitigating bias due to control overlap with the PSA discovery GWAS, *a*_*i*_ for GERA participants was calculated by substituting 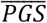 from out-of-sample UK Biobank controls (n=152,884). Upward classification resulting in biopsy eligibility occurred when 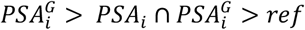, where *ref* was the biopsy referral threshold. Downward classification resulting in biopsy ineligibility was defined as: 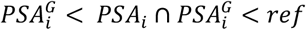. Net reclassification (NR) was summarized separately for cases and controls:

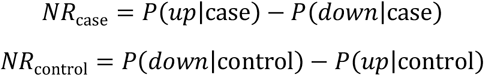

This is equivalent to tabulating the proportion of individuals in each biopsy eligibility category:

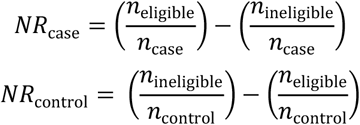

For each NR proportion, 95% confidence intervals were obtained using the normal approximation:

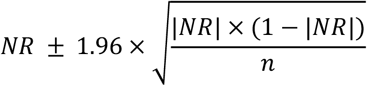

Next, we assessed the performance of risk prediction models for prostate cancer overall, aggressive prostate cancer, and non-aggressive prostate cancer in PCPT and SELECT. Since both studies were excluded from the PSA GWAS meta-analysis, *a*_*i*_ and 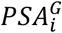 for PCPT and SELECT were calculated using 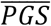 observed in each respective study. Consistent with the PGS_PSA_ validation analysis, pooled analyses included individuals of all ancestries who passed quality control filters. To facilitate ancestry-stratified analyses in SELECT, especially for aggressive disease, we combined AFR and AFR/EUR clusters into a single group (AFR pooled) and similarly pooled EAS and EAS/EUR (EAS pooled). Aggressive prostate cancer was defined as Gleason score ≥7, PSA ≥ 10 ng/mL, T3-T4 stage, and/or distant or nodal metastases. We compared AUC estimates for logistic regression models using the following predictors, alone and in combination: baseline PSA, genetically adjusted baseline PSA (PSA^G^) PGS_PSA_, prostate cancer risk score with original weights (PGS_269_)^32^ and weights corrected for index event bias (PGS_269_^adj^).

## DATA AVAILABILITY

UK Biobank data are publicly available by request from https://www.ukbiobank.ac.uk. To maintain individuals’ privacy, data on the GERA cohort are available by application to the Kaiser Permanente Research Bank (researchbank.kaiserpermanente.org). All PLCO genotype data is available in dbGaP18 under accession number phs001286.v2.p2 (https://identifiers.org/dbgap:phs001286.v2.p2). Companion phenotype data can be requested through the NCI Cancer Data Access System (CDAS) (https://cdas.cancer.gov/plco/). GWAS summary statistics are available directly from the PLCO Atlas GWAS Explorer website (https://exploregwas.cancer.gov/plco-atlas/) as well as accessed directly through API access (https://exploregwas.cancer.gov/plco-atlas/#/api-access). Genome-wide summary statistics for the PSA multi-ancestry meta-analysis and ancestry-stratified summary statistics for the development of the genomewide PSA polygenic score are available from: https://doi.org/10.5281/zenodo.7460134. Scoring files for fitting PSA polygenic scores are available from the PGS Catalog: www.pgscatalog.org/score/PGS003378/ and www.pgscatalog.org/score/PGS003379/

## CODE AVAILABILITY

Genome-wide association analyses were conducted using PLINK 2.0 version a3LM (https://www.coggenomics.org/plink/2.0/). Fixed-effects inverse-variance-weighted meta-analysis was performed with METAL using SCHEME STDERR (https://genome.sph.umich.edu/wiki/METAL_Documentation). Weights for the genome-wide polygenic score for PSA were estimated using PRS-CSx (https://github.com/getian107/PRScsx). Scripts for fitting polygenic scores, performing the index event bias analysis, and calculating genetically adjusted PSA values are available at: https://github.com/lkachuri/precision_PSA

## Notes

### Competing Interest Statement

John S. Witte is a non-employee, cofounder of Avail Bio. Hans Lilja is named on a patent for intact PSA assays and a patent for a statistical method to detect prostate cancer that is licensed to and commercialized by OPKO Health. HL receives royalties from sales of the test and has stock in OPKO Health.

### Author Declarations

The research was conducted with approved access to UK Biobank data under application number 14105 (PI: Witte). UK Biobank data are publicly available by request from https://www.ukbiobank.ac.uk. To maintain individuals' privacy, data on the GERA cohort are available by application to the Kaiser Permanente Research Bank (researchbank.kaiserpermanente.org). Informed consent was obtained from all study participants. UK Biobank received ethics approval from the Research Ethics Committee (REC reference: 11/NW/0382) in accordance with the UK Biobank Ethics and Governance Framework. The Kaiser Permanente Northern California Institutional Review Board and the University of California San Francisco Human Research Protection Program Committee on Human Research approved the GERA study. The institutional review board at the National Cancer Institute approved the PLCO study. The Vanderbilt Institutional Review Board approved the BioVU study. The Ethics Committee at Lund University approved the Malmo Diet & Cancer Study. The SELECT Coordinating Center institutional review board approved the SELECT study after its transition from a randomized control trial (clinicaltrials.gov NCT00006392) to an observational cohort. The PCPT was designed by investigators at the National Cancer Institute and SWOG (formerly the Southwest Oncology Group) and approved by institutional review boards at all sites.

### Summary of Updates

Corrected p-values in abstract and author affiliations

