## Supplementary Figures 1-8, Supplementary Tables 1-6, 8-26 for "Genetically Adjusted PSA Levels for Prostate Cancer Screening"

**Supplementary Figure 1: PSA values in men of European ancestry in the UK Biobank. a,** Violin plots and box plots visualize the distribution of PSA values across age groups. Lower and upper box plot hinges correspond to the first and third quartiles, or 25th and 75th percentiles, respectively. The upper and lower whiskers extend from each hinge to as a multiple of the inter-quartile range (IQR*1.5). **b,** Bar plots show the counts of UK Biobank participants with different numbers of PSA measurements, with the corresponding proportion of total GWAS sample size (N=26,491) accounted by each category.

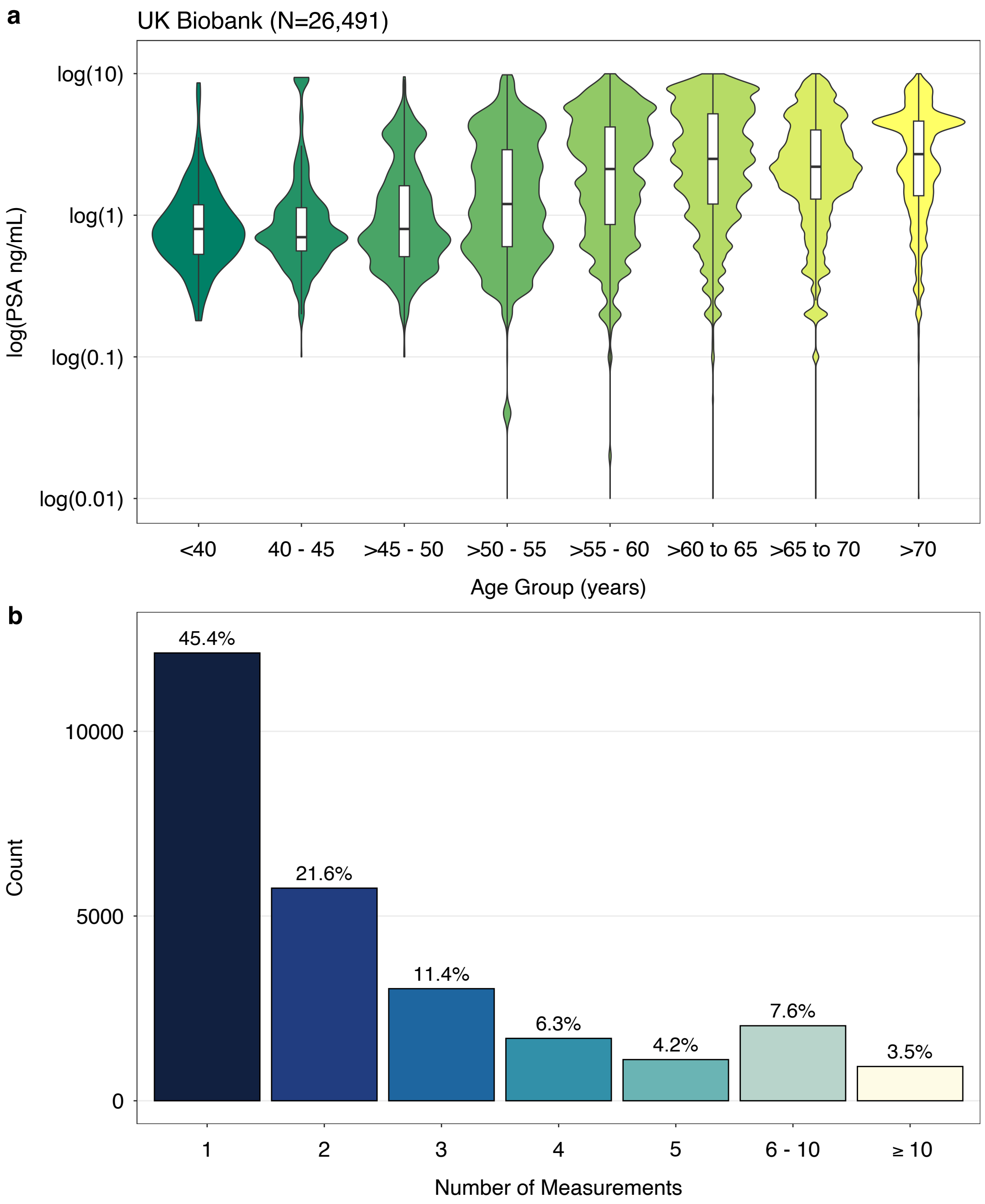

**Supplementary Figure 2: Cross-ancestry comparison of effect sizes for 128 PSA index variants.** Each panel compares effect sizes (β) in the European ancestry (EUR) inverse-variance-weighted fixed-effects meta-analysis (n=85,824) with effect sizes and corresponding 95% confidence intervals (CI) from **a,** African ancestry ($\beta_{AFR}$; n=3,509) **b,** Hispanic/Latino ($\beta_{HIS/LAT}$; n=3,098) **c,** East Asian ancestry ($\beta_{EAS}$; n=3,337) inverse-variance weighted fixed-effects meta-analyses. Index variants with $P<5\times10^{-8}$ were selected from the multi-ancestry GWAS meta-analysis (N=95,768) using linkage disequilibrium (LD) clumping (LD $r^2<0.01$ within 10Mb windows). Correlations between effect sizes were estimated using Spearman's rho (ρ) for all variants and excluding those in the 19q13.33 (*KLK3*) region. All p-values are two-sided.

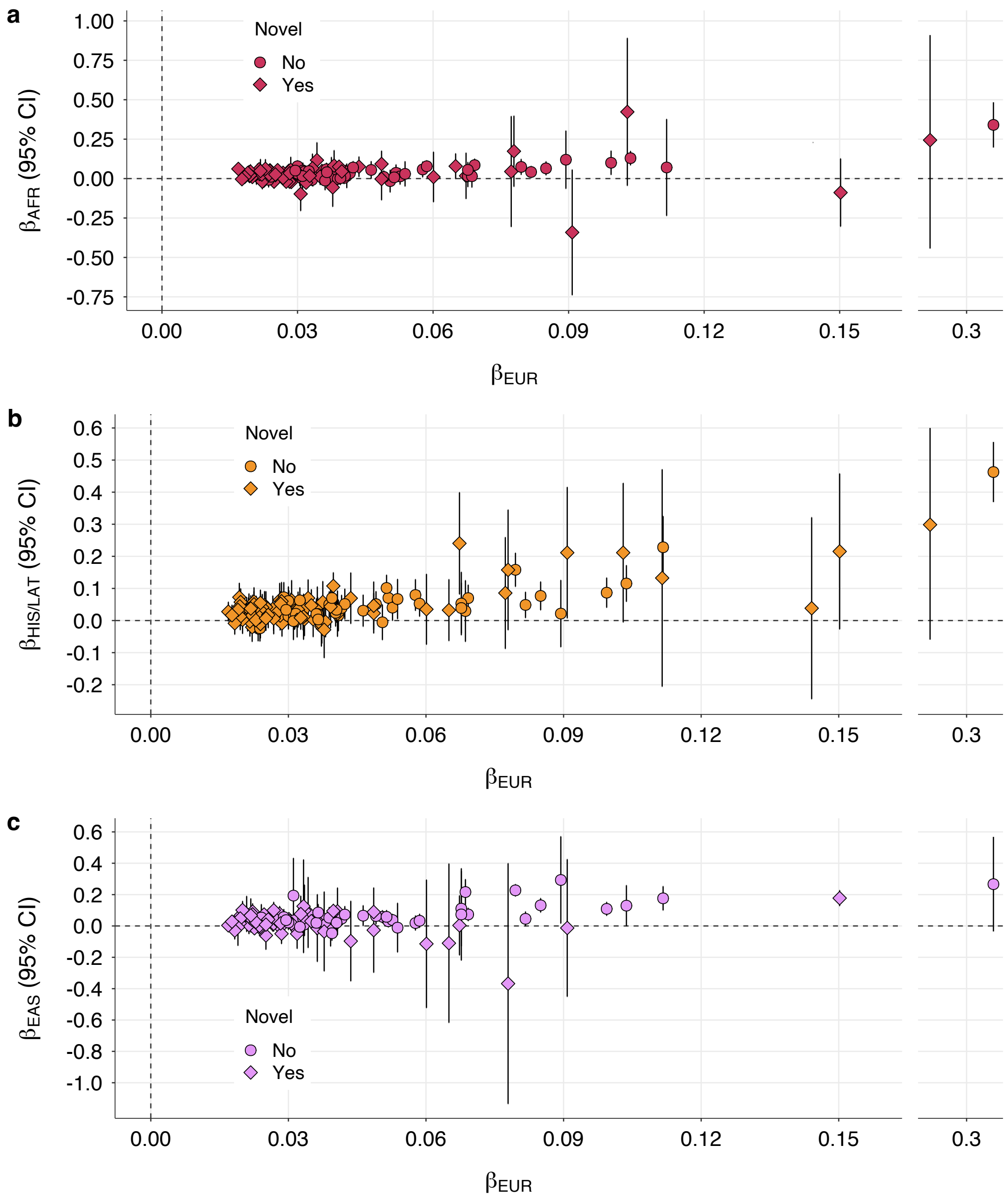

| | ρ (all variants) | p – value (all variants) | ρ (exclude 19q13.33) | p – value (exclude 19q13.33) |
|---|---|---|---|---|
| **AFR** | 0.273 | $1.95\times10^{-3}$ | 0.285 | $2.12\times10^{-3}$ |
| **HIS/LAT** | 0.479 | $1.14\times10^{-8}$ | 0.399 | $9.84\times10^{-6}$ |
| **EAS** | 0.166 | 0.069 | 0.097 | 0.31 |

**Supplementary Figure 3:** Calibration plots show the concordance between observed and predicted values generated from logistic regression models for overall incident prostate cancer (335 cases and 5548 controls) and aggressive prostate cancer (75 cases and 5548 controls) in the pooled, multi-ancestry population in the Prostate Cancer Prevention Trial (PCPT). Error bars show the 95% confidence intervals for the predictions within each bin.

PCPT: Prostate Cancer (Pooled)

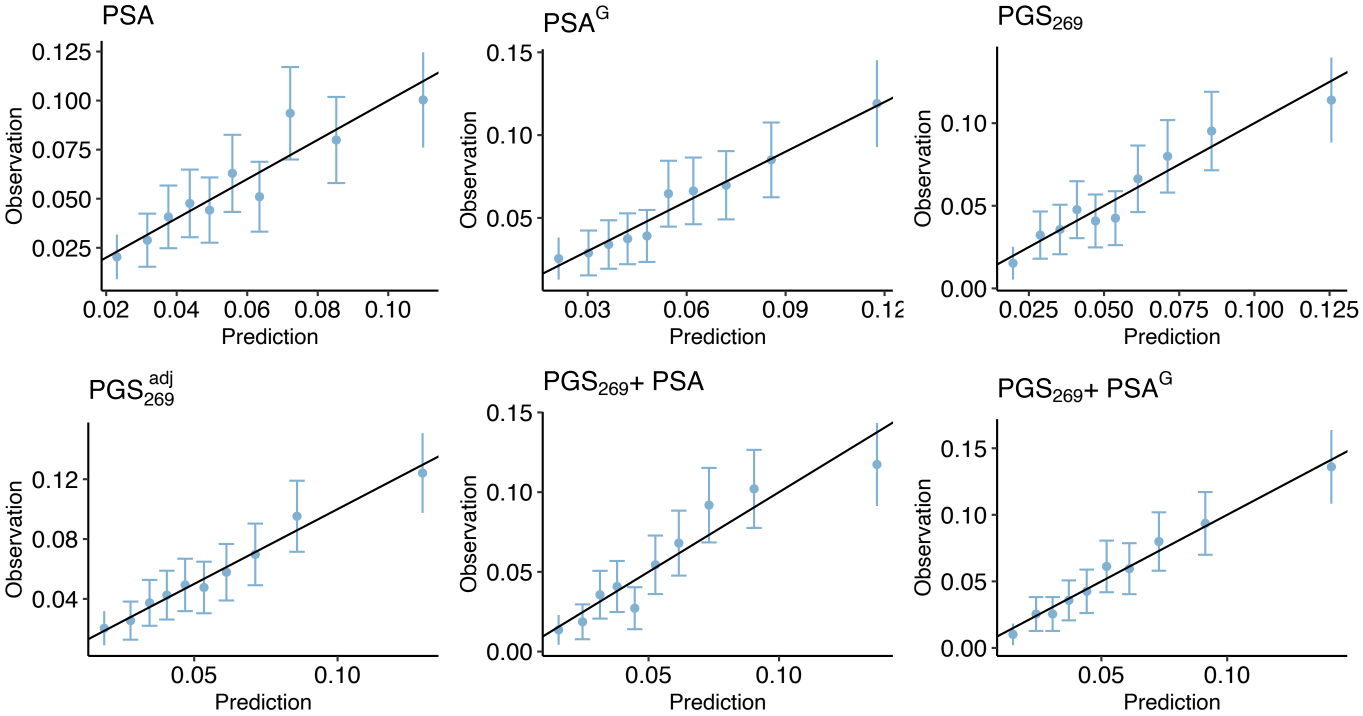

PCPT: Aggressive Prostate Cancer (Pooled)

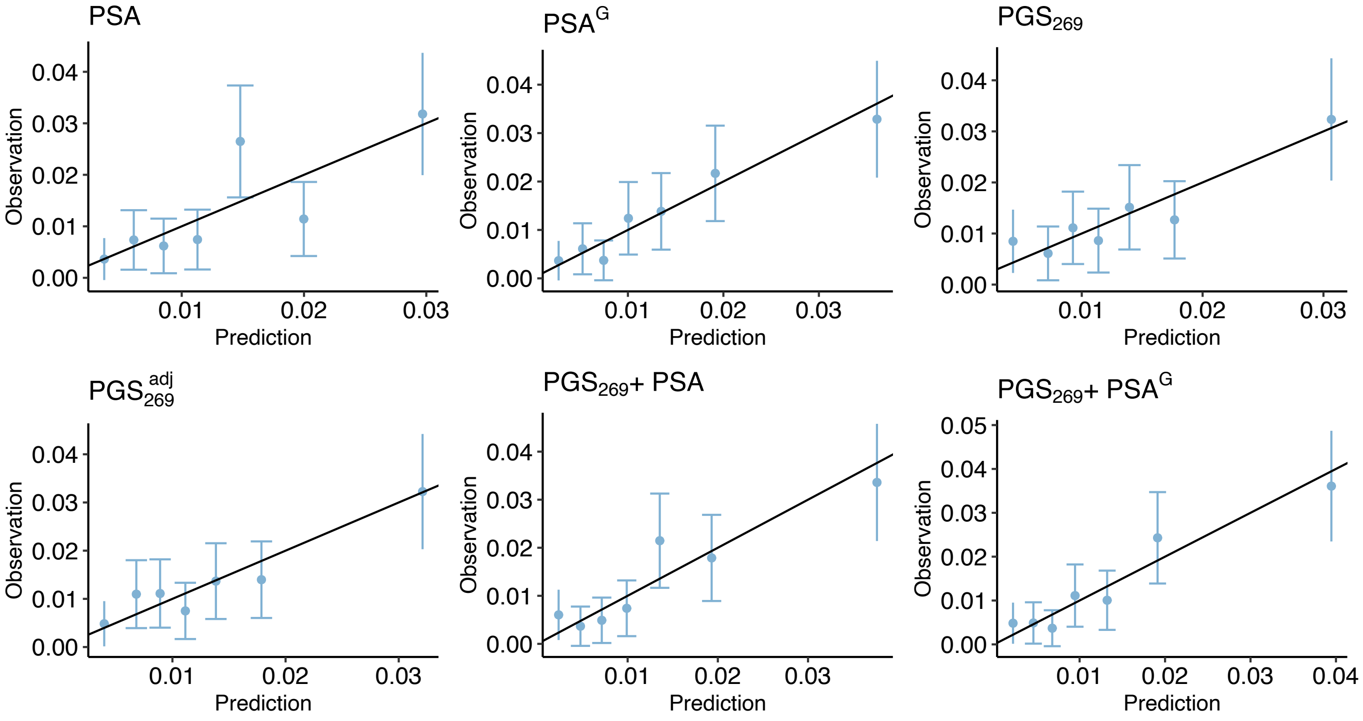

**Supplementary Figure 4:** Calibration plots show the concordance between observed and predicted values generated from logistic regression models for incident prostate cancer (323 cases and 5402 controls) and aggressive prostate cancer (71 cases and 5402 controls) in men of European ancestry in the Prostate Cancer Prevention Trial (PCPT). Error bars show the 95% confidence intervals for the predictions within each bin.

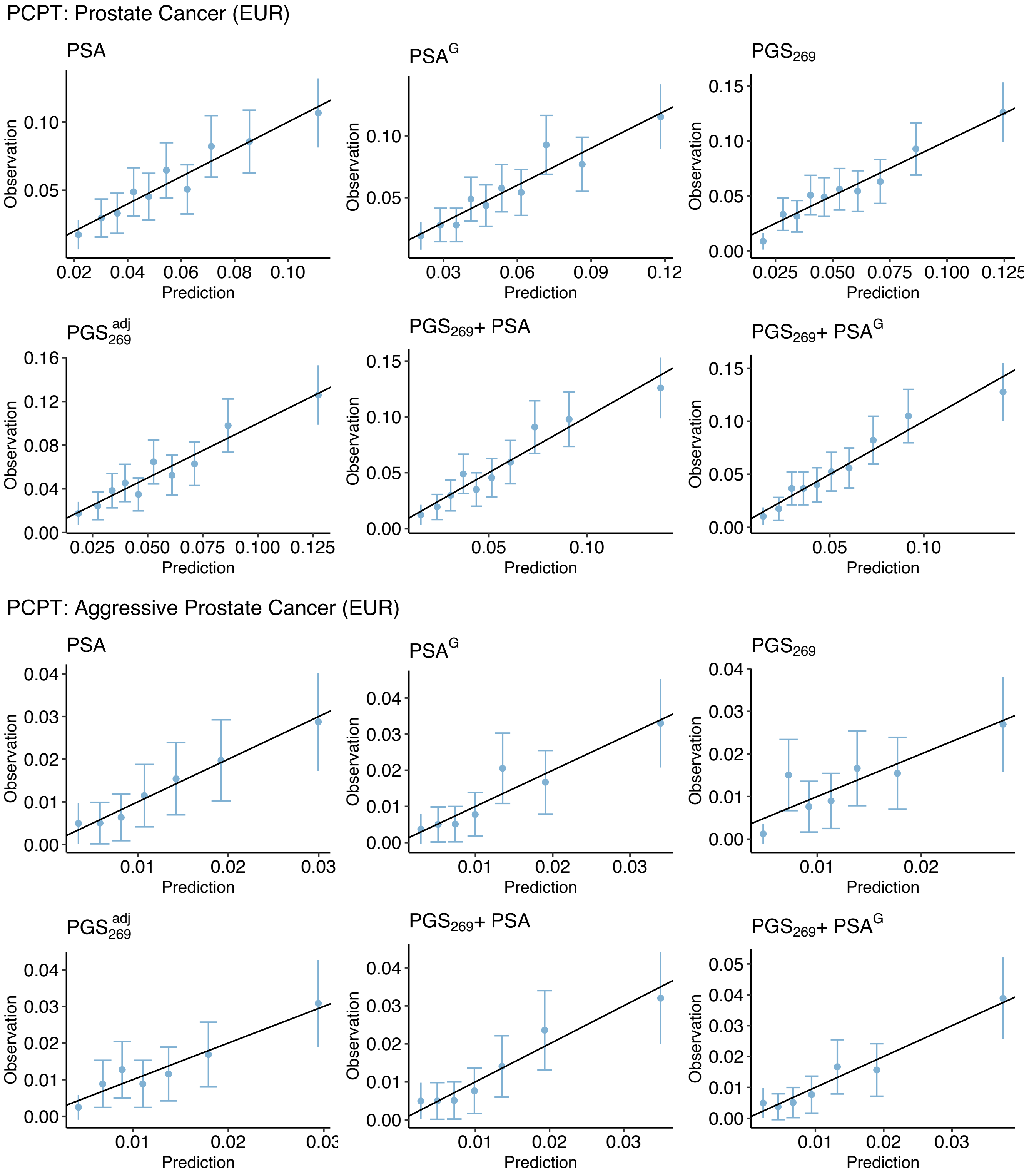

**Supplementary Figure 5:** Calibration plots show the concordance between observed and predicted values generated from logistic regression models for incident prostate cancer in the Selenium and Vitamin E Cancer Prevention Trial (SELECT). The pooled analysis includes 572 prostate cancer cases and 23,667 controls. European ancestry (EUR≥0.80) group includes 467 cases and 20,173 controls. Error bars show the 95% confidence intervals for the predictions within each bin.

SELECT: Prostate Cancer (Pooled)

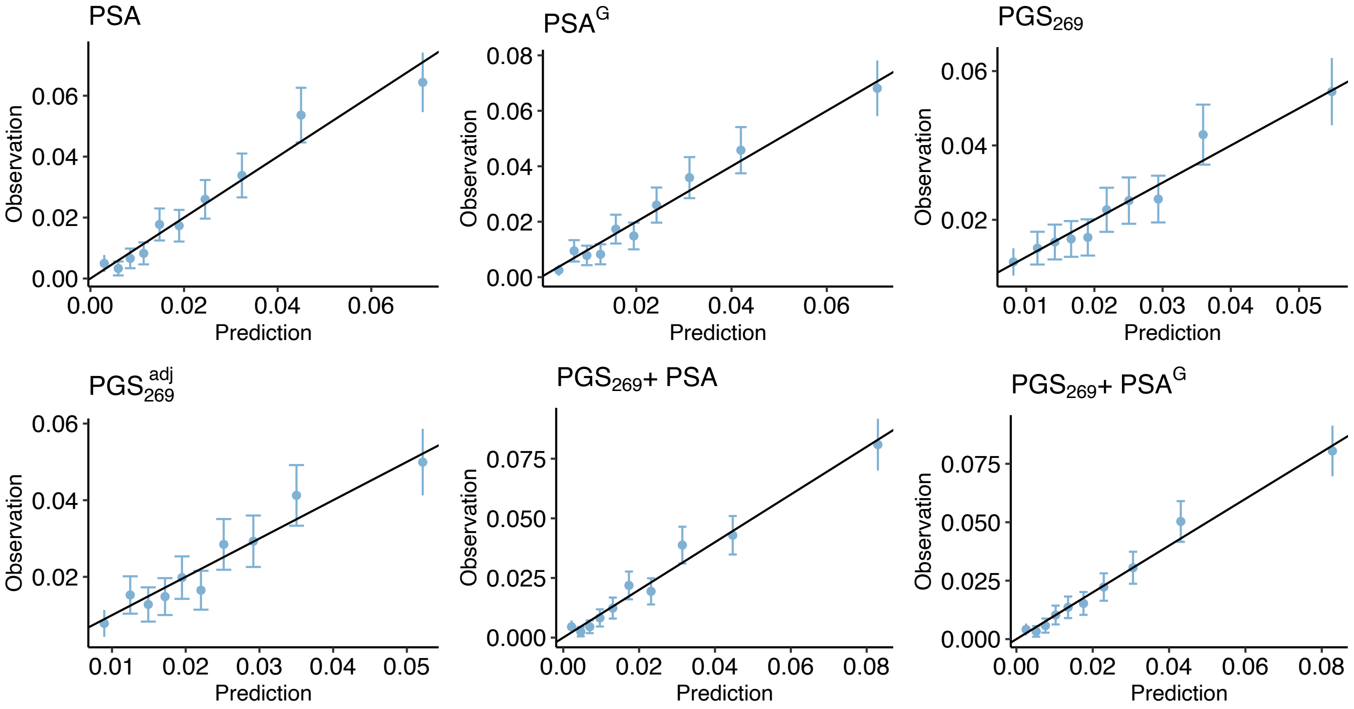

SELECT: Prostate Cancer (EUR)

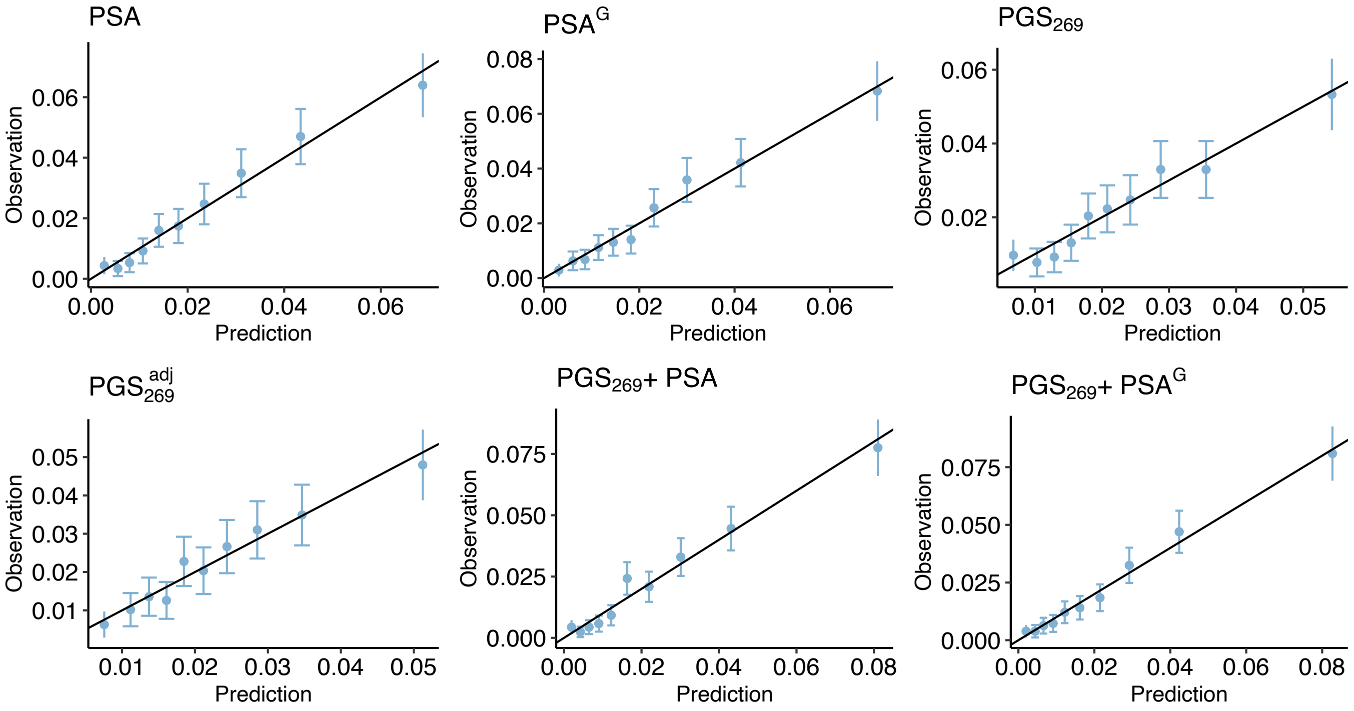

**Supplementary Figure 6:** Calibration plots show the concordance between observed and predicted values generated from logistic regression models for prostate cancer in the Selenium and Vitamin E Cancer Prevention Trial (SELECT). Pooled African ancestry group includes men of African ancestry (AFR≥0.80) and admixed African and European ancestry (0.20<AFR/EUR<0.80), for a total of 88 cases and 2733 controls. Error bars show the 95% confidence intervals for the predictions within each bin.

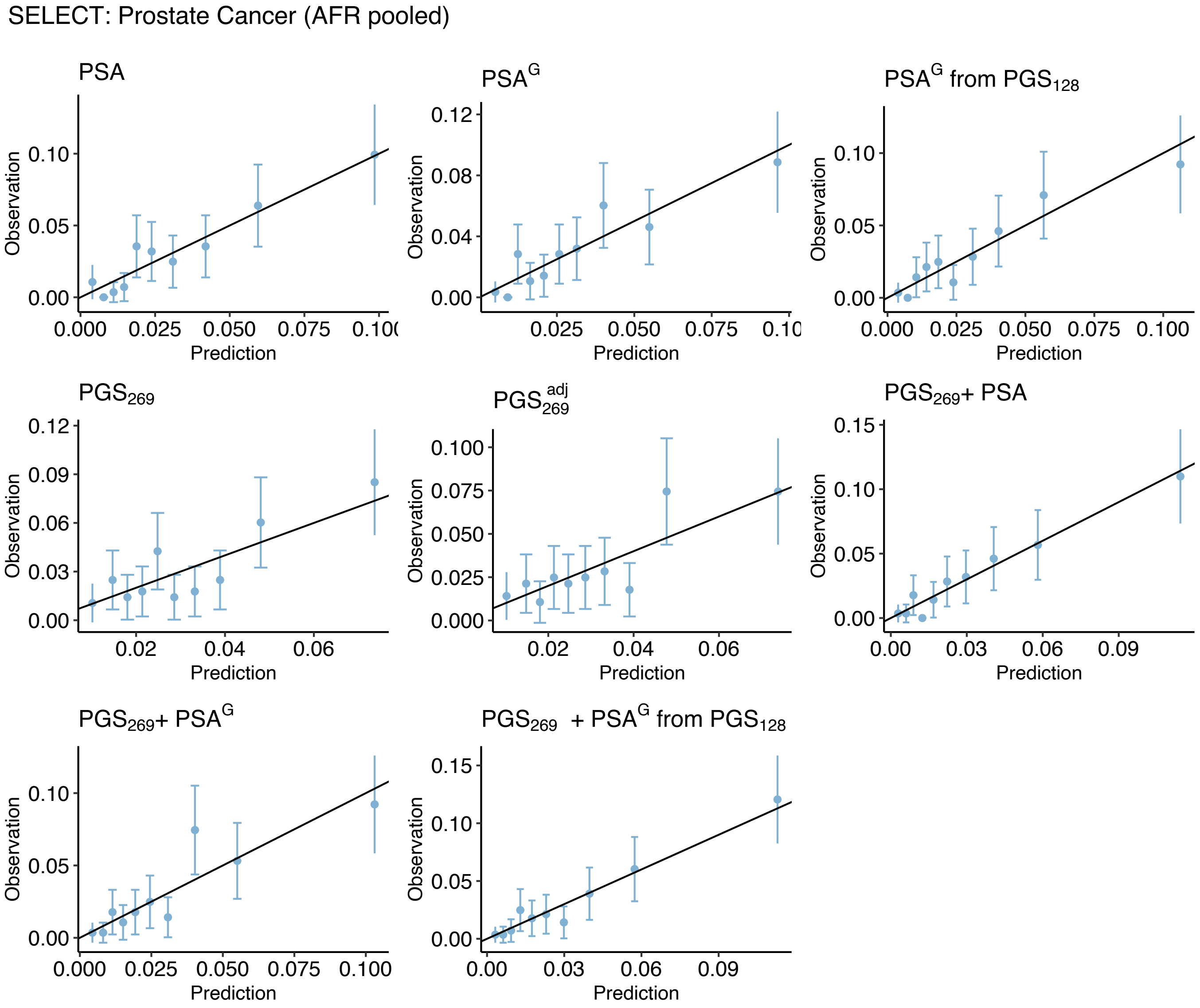

**Supplementary Figure 7:** Calibration plots show the concordance between observed and predicted values generated from logistic regression models for incident aggressive prostate cancer in the Selenium and Vitamin E Cancer Prevention Trial (SELECT). The pooled analysis included 106 cases and 23,667 controls. European ancestry (EUR≥0.80) group includes 85 cases and 20,173 controls. Error bars show the 95% confidence intervals for the predictions within each bin.

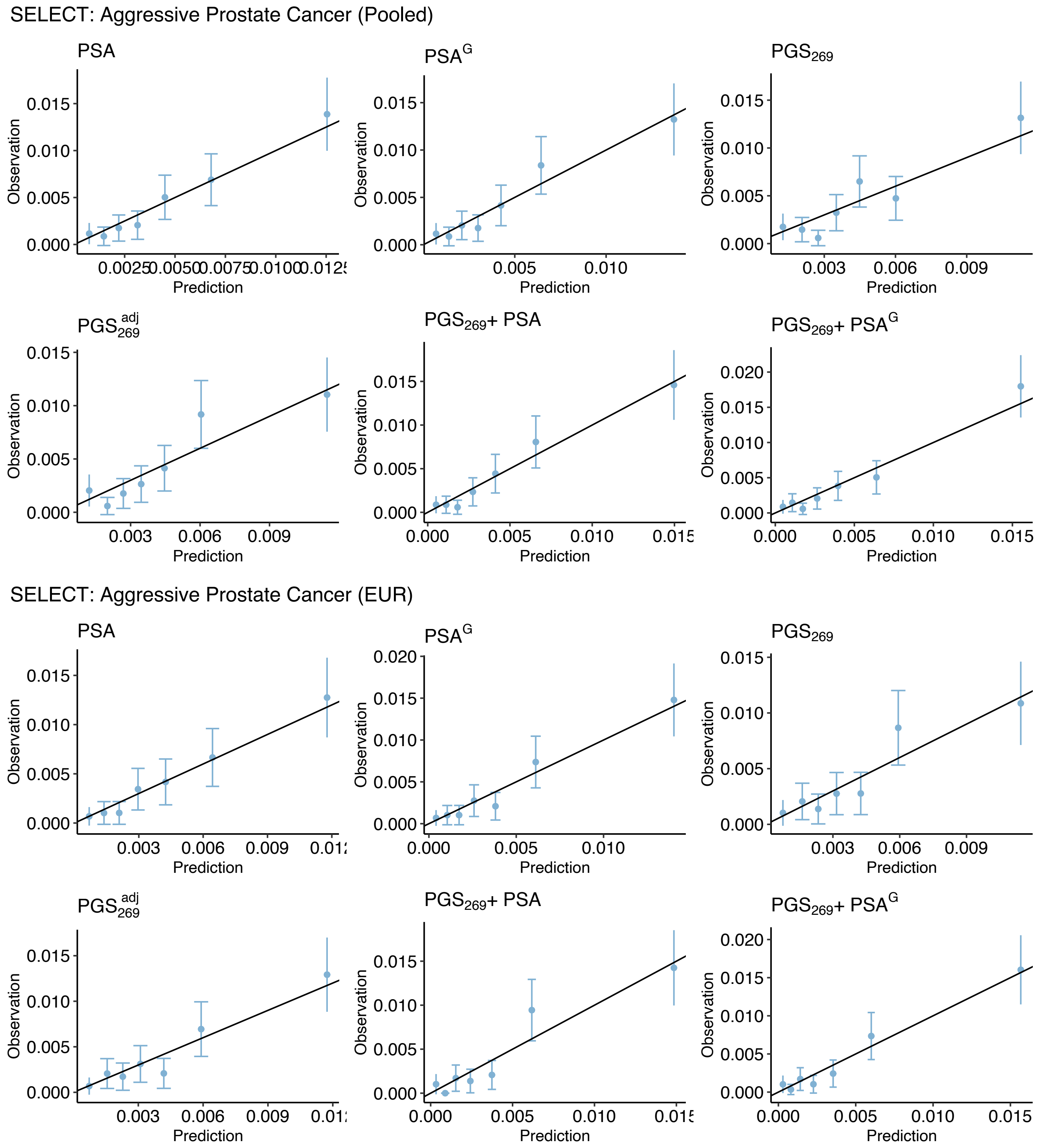

**Supplementary Figure 8:** Calibration plots show the concordance between observed and predicted values generated from logistic regression models for incident aggressive prostate cancer in the Selenium and Vitamin E Cancer Prevention Trial (SELECT). Pooled African ancestry subgroup includes men of African ancestry (AFR≥0.80) and admixed African and European ancestry (0.20<AFR/EUR<0.80), for a total of 18 cases and 2733 controls. Error bars show the 95% confidence intervals for the predictions within each bin. Error bars are absent for bins that had too few observations.

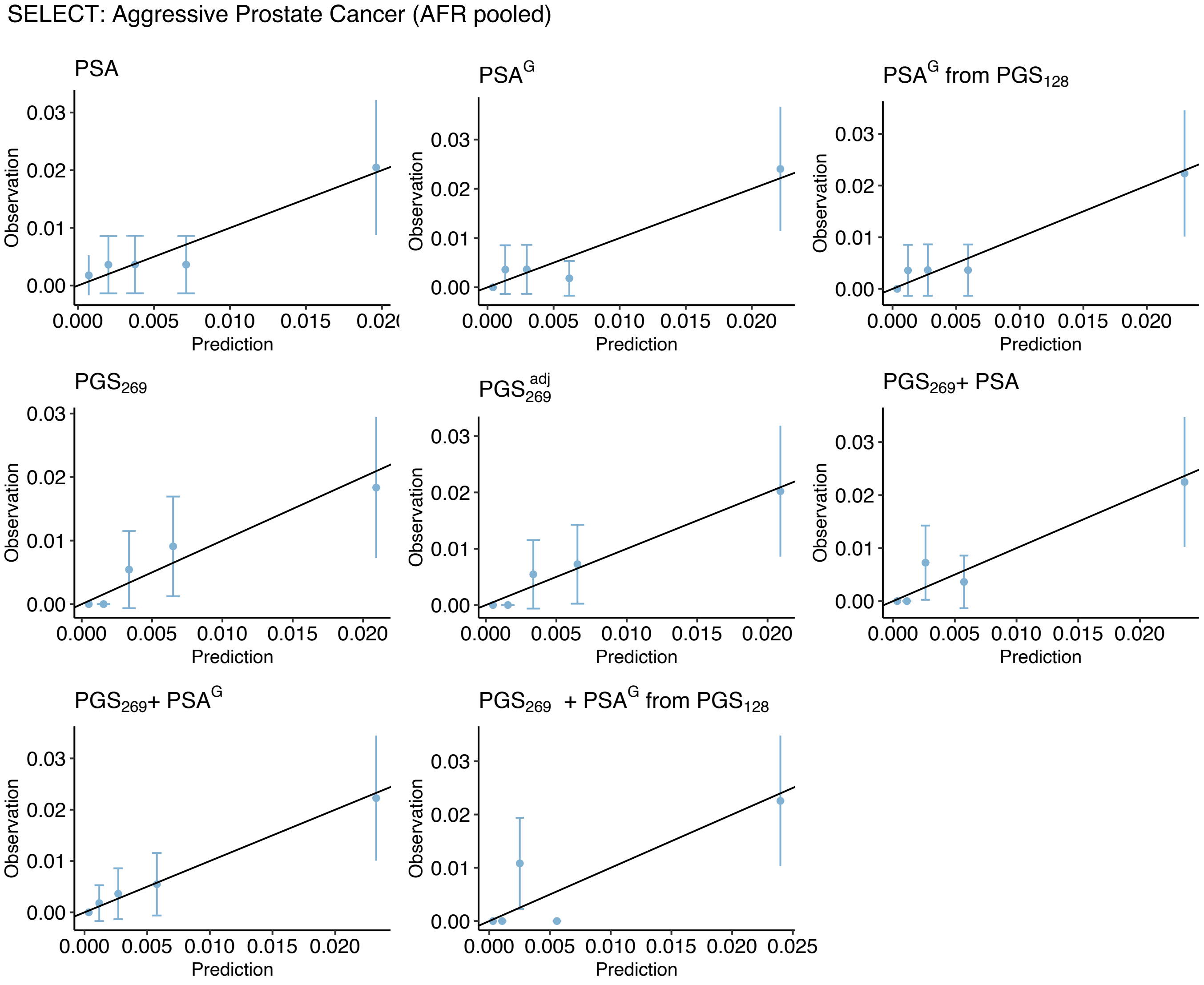

**Supplementary Table 1: Heritability ($h^2$) of PSA levels in men of European ancestry without prostate cancer.** Estimates are compared between heritability methods that use individual-level data and GWAS summary statistics, and between two approaches to modelling longitudinal PSA values: summarizing multiple PSA measurements per individual as the median compared to estimating the heritability of individual-specific random intercepts derived from a linear mixed model of longitudinal PSA values.

| Phenotype | Method | Genetic Relatedness Matrix Parameters: Kinship | LD $r^2$ | INFO | ($N_{SNP}$) | $h^2$ | (95% CI) |
|---|---|---|---|---|---|---|---|
| UKB median log(PSA) n=26,491 | GCTA | 0.025 | 0.80 | 0.80 | (2,109,931) | 0.405 | (0.353 - 0.456) |
| | | | 0.80 | Typed | (563,257) | 0.372 | (0.326 - 0.418) |
| | LDAK | 0.025 | 0.80 | 0.80 | (2,109,931) | 0.302 | (0.266 - 0.339) |
| | | | 0.98 | 0.95 | (2,629,389) | 0.244 | (0.213 - 0.276) |
| | | | 0.98 | Typed | (563,257) | 0.295 | (0.261 - 0.329) |
| | GCTA | 0.05 | 0.80 | 0.80 | (2,109,931) | 0.408 | (0.359 - 0.457) |
| | | | 0.80 | Typed | (563,257) | 0.376 | (0.332 - 0.420) |
| | LDAK | 0.05 | 0.80 | 0.80 | (2,109,931) | 0.296 | (0.261 - 0.331) |
| | | | 0.98 | 0.95 | (2,629,389) | 0.239 | (0.209 - 0.269) |
| | | | 0.98 | Typed | (563,257) | 0.288 | (0.256 - 0.320) |
| UKB random intercept n=26,491 | GCTA | 0.025 | 0.80 | 0.80 | (2,109,931) | 0.351 | (0.300 - 0.402) |
| | | | 0.80 | Typed | (563,257) | 0.327 | (0.280 - 0.373) |
| | LDAK | 0.025 | 0.80 | 0.80 | (2,109,931) | 0.270 | (0.233 - 0.307) |
| | | | 0.98 | 0.95 | (2,629,389) | 0.210 | (0.179 - 0.242) |
| | | | 0.98 | Typed | (563,257) | 0.255 | (0.221 - 0.289) |
| | GCTA | 0.05 | 0.80 | 0.80 | (2,109,931) | 0.358 | (0.309 - 0.407) |
| | | | 0.80 | Typed | (563,257) | 0.333 | (0.289 - 0.377) |
| | LDAK | 0.05 | 0.80 | 0.80 | (2,109,931) | 0.259 | (0.224 - 0.294) |
| | | | 0.98 | 0.95 | (2,629,389) | 0.207 | (0.178 - 0.237) |
| | | | 0.98 | Typed | (563,257) | 0.251 | (0.220 - 0.283) |

| Summary Statistics | Method | Reference Panel: Variants | LD Dataset | INFO | ($N_{SNP}$) | $h^2$ | (95% CI) |
|---|---|---|---|---|---|---|---|
| UKB median log(PSA) n=26,491 | LDAK | HapMap3 | UKB GBR | 0.30 | (1,127,342) | 0.385 | (0.310 - 0.460) |
| | | | | 0.30 | (1,127,337)[1] | 0.349 | (0.275 - 0.424) |
| | | | | 0.95 | (1,118,359)[1] | 0.350 | (0.275 - 0.425) |
| | HDL | HapMap3 | UKB GBR | 0.30 | (1,029,876) | 0.252 | (0.207 - 0.298) |
| | LDSR | HapMap3 | 1000G EUR | 0.30 | (1,133,218) | 0.206 | (0.154 - 0.257) |
| Meta-analysis log(PSA) n=85,824 | LDAK | HapMap3 | UKB GBR | 0.30 | (1,168,859) | 0.332 | (0.307 - 0.357) |
| | | | | 0.30 | (1,168,854)[1] | 0.300 | (0.287 - 0.311) |
| | HDL | HapMap3 | UKB EUR | 0.30 | (1,029,876) | 0.195 | (0.157 - 0.233) |
| | LDSR | HapMap3 | 1000G EUR | 0.30 | (1,136,374) | 0.188 | (0.151 - 0.225) |

[1] Excluding variants with large effect, using cutoff=0.01 for maximum variance explained

| | |
|---|---|
| INFO | Minimum imputation quality |
| HDL | High-definition likelihood method, as described in Ning et al. (2020)[26] |
| GBR | British white individuals |
| LDSR | LD score regression |

**Supplementary Table 2: Genome-wide association study (GWAS) of PSA levels in the UK Biobank (N=26,491).** Genome-wide significant ($P<5\times10^{-8}$) lead variants were identified using LD-based clumping at $r^2<0.01$ in ± 10Mb windows. All GWAS p-values are two-sided and derived from linear regression models.

| Region | Position (b37) | Nearest Gene | Variant ID | EA | OA | EAF | β[1] | (SE) | P | $P_{RI}$ |
|---|---|---|---|---|---|---|---|---|---|---|
| 1p22.3 | 88197017 | *PKN2-AS1* | rs12731909 | C | T | 0.484 | 0.041 | (0.007) | 9.12E-10 | 3.81E-09 |
| 2p21 | 43594483 | *THADA* | rs79625619 | T | A | 0.100 | -0.063 | (0.011) | 1.27E-08 | 3.48E-08 |
| 2p16.1 | 60762502 | *BCL11A* | rs2556378 | T | G | 0.161 | 0.058 | (0.009) | 1.45E-10 | 3.56E-10 |
| 2p15 | 63277843 | *OTX1* | rs58235267 | G | C | 0.484 | 0.038 | (0.007) | 1.50E-08 | 5.41E-07 |
| 3q27.3 | 186754957 | *ST6GAL1* | rs3936289 | C | T | 0.242 | 0.041 | (0.008) | 1.58E-07 | 7.07E-09 |
| 5p15.33 | 1341101 | *CLPTM1L* | rs31487 | C | G | 0.444 | -0.064 | (0.007) | 1.78E-21 | 6.11E-17 |
| 6p21.32 | 32682812 | *HLA-DQB1-AS1* | rs9275602 | A | C | 0.191 | -0.048 | (0.009) | 1.83E-08 | 9.64E-07 |
| 6p21.1 | 43710381 | *POLR1C* | rs10807290 | C | T | 0.475 | 0.045 | (0.007) | 2.21E-11 | 1.13E-07 |
| 7p15.2 | 27989768 | *JAZF1* | rs739704 | A | G | 0.253 | -0.043 | (0.008) | 2.02E-08 | 1.26E-04 |
| 8p21.2 | 23535219 | *NKX3-1* | rs13262167 | C | A | 0.414 | 0.067 | (0.007) | 1.01E-22 | 1.26E-19 |
| 8q24.21 | 128318755 | *CASC8 – PCAT1* | rs10107982 | C | T | 0.277 | -0.053 | (0.008) | 1.23E-12 | 6.15E-09 |
| 8q24.21 | 128413305 | *CASC8* | rs6983267 | T | G | 0.481 | -0.054 | (0.007) | 8.70E-16 | 2.45E-14 |
| 10q11.22 | 51549496 | *MSMB* | rs10993994 | T | C | 0.386 | 0.087 | (0.007) | 1.06E-36 | 4.87E-26 |
| 10q26.12 | 122633607 | *WDR11* | rs7080550 | G | C | 0.365 | 0.049 | (0.007) | 1.54E-12 | 2.91E-10 |
| 10q26.12 | 123048008 | *LINC01153* | rs10886900 | G | A | 0.231 | 0.103 | (0.008 | 1.75E-38 | 2.49E-32 |
| 10q26.13 | 123186164 | *RN7SKP167* | rs201552570 | C | CA | 0.052 | -0.118 | (0.015) | 7.02E-15 | 4.56E-13 |
| 10q26.13 | 123349324 | *FGFR2* | rs45631563 | T | A | 0.046 | 0.090 | (0.016) | 2.62E-08 | 3.30E-05 |
| 11p13 | 34770077 | *EHF – APIP* | rs10742324 | C | G | 0.291 | -0.054 | (0.007) | 3.54E-13 | 8.78E-10 |
| 12q24.21 | 115094260 | *TBX3* | rs11067228 | G | A | 0.451 | -0.045 | (0.007) | 1.65E-11 | 2.46E-09 |
| 13q14.3 | 51130374 | *DLEU1* | rs35681675 | TC | T | 0.389 | 0.042 | (0.007) | 1.37E-09 | 1.29E-08 |
| 17q12 | 36097775 | *HNF1B* | rs11263761 | G | A | 0.481 | -0.062 | (0.007) | 4.32E-20 | 8.87E-15 |
| 18q23 | 76765837 | *SALL3* | rs6506878 | T | G | 0.320 | -0.041 | (0.007) | 1.35E-08 | 6.75E-07 |
| 19q13.33 | 51323947 | *KLK1* | rs2659104 | A | G | 0.317 | 0.055 | (0.007) | 3.39E-14 | 9.62E-10 |
| 19q13.33 | 51340794 | *KLK15* | rs2659053 | A | G | 0.394 | 0.082 | (0.007) | 5.38E-33 | 1.83E-27 |
| 19q13.33 | 51360768 | *KLK3* | rs266877 | A | G | 0.048 | -0.094 | (0.016) | 2.29E-09 | 3.49E-09 |
| 19q13.33 | 51361757 | *KLK3* | rs17632542 | C | T | 0.077 | -0.375 | (0.012) | 1.69E-202 | 4.16E-155 |
| 19q13.33 | 51399496 | *KLKP1* | rs2659110 | T | C | 0.114 | -0.076 | (0.011) | 6.39E-13 | 9.41E-15 |
| 20q11.21-q11.22 | 32127671 | *CBFA2T2* | rs2150165 | T | A | 0.104 | 0.069 | (0.011) | 3.60E-10 | 4.72E-07 |
| Xp22.11 | 24065653 | *EIF2S3 – KLHL15* | rs12389566 | T | C | 0.331 | 0.033 | (0.005) | 1.73E-10 | 8.78E-08 |
| Xq13.1 | 70117073 | *TEX11* | rs186347618 | T | A | 0.368 | 0.039 | (0.005) | 7.89E-15 | 6.66E-13 |

[1] Per allele change in log(PSA)

Abbreviations:

| | |
|---|---|
| EA | Effect allele for which the change in log(PSA) is modelled |
| OA | Other allele |
| EAF | Effect allele frequency |
| $P_{RI}$ | P-value from the GWAS of individual-specific random intercepts estimated in |

**Supplementary Table 3: Association estimates for 128 index variants associated with PSA levels in the multi-ancestry meta-analysis (N=95,768).** Genome-wide significant ($P<5\times10^{-8}$) index variants were selected from fixed-effects, inverse-variance weighted meta-analysis results generated by METAL using linkage disequilibrium (LD) clumping at $r^2<0.01$ within ± 10Mb windows. Associations are compared between METAL ($P_{METAL}$), where heterogeneity is quantified using Cochran's Q statistic ($P_{Het-Q}$), and MR-MEGA ($P_{MEGA}$), an alternative approach to cross-ancestry meta-analysis that uses meta-regression to quantify heterogeneity due to ancestry ($P_{Het-Anc}$) separately from residual heterogeneity due to other factors ($P_{Het-Res}$). All p-values are two-sided.

| Region | Position (b37) | Variant ID | Effect Allele | Other Allele | Fixed-effects Meta-analysis (METAL) | | | | | Meta-Regression (MR-MEGA) | | | New[2] |
|---|---|---|---|---|---|---|---|---|---|---|---|---|---|
| | | | | | EAF | β[1] | SE | $P_{METAL}$ | $P_{Het-Q}$ | $P_{MEGA}$ | $P_{Het-Anc}$ | $P_{Het-Res}$ | |
| 1p32.3 | 51237409 | rs12569177 | C | G | 0.428 | 0.026 | (0.004) | 3.2E-13 | 0.89 | 3.5E-11 | 0.59 | 0.80 | yes |
| 1p22.3 | 88199778 | rs12131120 | T | A | 0.493 | 0.033 | (0.004) | 1.6E-19 | 0.38 | 3.9E-18 | 0.12 | 0.36 | no |
| 1p13.2 | 112264274 | rs2076591 | C | T | 0.216 | 0.032 | (0.004) | 2.6E-13 | 0.094 | 4.9E-12 | 0.10 | 0.77 | yes |
| 1q21.3 | 154868055 | rs4845681 | T | G | 0.412 | 0.022 | (0.004) | 7.7E-09 | 0.15 | 5.3E-07 | 0.74 | 0.13 | yes |
| 1q25.3 | 184191716 | rs12046452 | C | T | 0.622 | 0.021 | (0.004) | 9.0E-09 | 0.95 | 8.7E-07 | 0.95 | 0.19 | yes |
| 1q32.1 | 205632217 | rs71152447 | G | GGCCGAC AGCCCTTC TGCTGGCT CGGTGGG GCCCAGC | 0.047 | 0.163 | (0.024) | 9.9E-12 | 0.27 | 1.2E-09 | 0.80 | 0.025 | no |
| 1q32.1 | 205636334 | rs4951018 | C | A | 0.230 | 0.032 | (0.004) | 2.7E-13 | 0.52 | 2.5E-12 | 0.058 | 0.092 | no |
| 1q41 | 219694903 | rs12034581 | C | A | 0.495 | 0.025 | (0.004) | 2.5E-12 | 0.074 | 3.5E-11 | 0.14 | 0.49 | yes |
| 1q41 | 219954267 | rs6664688 | G | A | 0.216 | 0.025 | (0.005) | 3.9E-08 | 0.025 | 1.9E-07 | 0.084 | 1.2E-03 | yes |
| 2p24.2 | 18911674 | rs12710685 | G | C | 0.105 | 0.033 | (0.006) | 2.3E-08 | 0.26 | 1.7E-06 | 0.73 | 0.57 | yes |
| 2p24.1 | 20880833 | rs10193919 | T | C | 0.345 | 0.031 | (0.004) | 1.5E-15 | 0.84 | 3.7E-13 | 0.95 | 0.25 | yes |
| 2p23.3 | 25463871 | rs734693 | T | C | 0.715 | 0.023 | (0.004) | 3.9E-08 | 0.054 | 7.4E-08 | 0.040 | 0.37 | yes |
| 2p21 | 43072828 | rs1123695 | G | C | 0.659 | 0.031 | (0.004) | 3.3E-16 | 0.80 | 6.7E-14 | 0.89 | 0.81 | yes |
| 2p21 | 43618819 | rs11899863 | C | T | 0.902 | 0.045 | (0.006) | 1.7E-13 | 0.48 | 2.2E-11 | 0.56 | 0.078 | yes |
| 2p16.1 | 60759747 | rs2556375 | G | T | 0.176 | 0.057 | (0.005) | 1.8E-32 | 0.33 | 5.2E-30 | 0.54 | 0.42 | no |
| 2p15 | 63277843 | rs58235267 | G | C | 0.482 | 0.026 | (0.004) | 4.9E-13 | 0.014 | 1.1E-12 | 0.016 | 0.57 | yes |
| 2p13.2 | 72059504 | rs11679946 | T | C | 0.674 | 0.022 | (0.004) | 2.6E-08 | 0.23 | 1.3E-06 | 0.79 | 0.35 | yes |
| 2q33.1 | 198929083 | rs34388051 | A | G | 0.671 | 0.022 | (0.004) | 2.2E-08 | 0.62 | 1.4E-06 | 0.65 | 0.52 | yes |
| 3q13.2 | 113290793 | rs4682495 | A | C | 0.573 | 0.021 | (0.004) | 2.9E-09 | 0.12 | 1.6E-07 | 0.40 | 0.081 | no |
| 3q21.3 | 128067275 | rs11709611 | C | T | 0.120 | 0.036 | (0.006) | 2.0E-10 | 0.97 | 2.7E-08 | 0.98 | 0.90 | yes |
| 3q23 | 141115219 | rs1582874 | C | T | 0.442 | 0.030 | (0.004) | 1.0E-16 | 0.36 | 1.1E-14 | 0.37 | 0.79 | no |

| | | | | | | | | | | | | | |
|---|---|---|---|---|---|---|---|---|---|---|---|---|---|
| 3q25.33 | 160130592 | rs10673842 | TTATC | T | 0.522 | 0.023 | (0.004) | 2.2E-08 | 0.11 | 2.9E-07 | 0.15 | 0.46 | yes |
| 3q27.2 | 185246981 | rs4012714 | A | G | 0.579 | 0.025 | (0.004) | 1.1E-08 | 0.11 | 7.7E-08 | 0.11 | 0.81 | yes |
| 3q27.3 | 186746994 | rs12629450 | C | G | 0.171 | 0.031 | (0.005) | 2.6E-10 | 0.65 | 2.2E-08 | 0.58 | 0.69 | yes |
| 3q28 | 189482770 | rs6765473 | T | C | 0.881 | 0.037 | (0.006) | 3.2E-10 | 0.31 | 6.0E-09 | 0.23 | 0.73 | yes |
| 4p15.2 | 24410024 | rs4276269 | G | A | 0.263 | 0.023 | (0.004) | 2.0E-08 | 0.91 | 1.5E-06 | 0.82 | 0.59 | yes |
| 4q31.22 | 146890261 | rs9968429 | G | A | 0.370 | 0.032 | (0.004) | 6.3E-18 | 0.061 | 1.0E-16 | 0.056 | 0.29 | no |
| 4q32.1 | 157497921 | rs13115840 | A | G | 0.355 | 0.031 | (0.004) | 4.3E-17 | 0.70 | 2.7E-14 | 0.86 | 0.56 | no |
| 5p15.33 | 1285974 | rs7705526 | A | C | 0.331 | 0.023 | (0.004) | 1.2E-08 | 0.46 | 6.0E-07 | 0.47 | 0.97 | no |
| 5p15.33 | 1322087 | rs401681 | C | T | 0.560 | 0.059 | (0.004) | 7.0E-54 | 0.47 | 1.5E-52 | 0.12 | 0.84 | no |
| 5q14.3 | 92052005 | rs7732515 | A | T | 0.227 | 0.033 | (0.004) | 3.5E-14 | 0.52 | 5.3E-12 | 0.86 | 0.46 | yes |
| 5q35.2 | 172945918 | rs889017 | C | A | 0.230 | 0.024 | (0.004) | 2.6E-08 | 0.081 | 1.3E-07 | 0.074 | 0.19 | yes |
| 6p22.3 | 15463139 | rs926309 | T | G | 0.374 | 0.026 | (0.004) | 1.6E-12 | 0.81 | 1.4E-10 | 0.69 | 0.88 | yes |
| 6p22.3 | 16515154 | rs236961 | T | C | 0.353 | 0.026 | (0.004) | 8.6E-12 | 0.10 | 7.6E-11 | 0.10 | 0.44 | yes |
| 6p22.1 | 29915061 | rs2248162 | C | T | 0.666 | 0.025 | (0.004) | 3.6E-10 | 0.26 | 9.7E-09 | 0.24 | 0.36 | yes |
| 6p21.31 | 34212537 | rs551980123 | TG | T | 0.043 | 0.069 | (0.011) | 1.6E-10 | 0.14 | 2.1E-08 | 0.91 | 0.14 | yes |
| 6p21.1 | 43711981 | rs1535507 | T | C | 0.164 | 0.048 | (0.005) | 1.5E-22 | 0.063 | 1.4E-22 | 1.8E-03 | 0.044 | no |
| 6q22.33 | 129168057 | rs73583119 | A | T | 0.929 | 0.193 | (0.035) | 3.5E-08 | 0.12 | - | - | - | yes |
| 7p15.3 | 20909622 | rs204592 | G | A | 0.517 | 0.022 | (0.004) | 1.2E-09 | 0.24 | 1.9E-08 | 0.21 | 0.45 | yes |
| 7p15.3 | 21839896 | rs6956349 | T | C | 0.427 | 0.021 | (0.004) | 8.6E-09 | 0.78 | 6.1E-07 | 0.67 | 0.56 | yes |
| 7p15.2 | 27281792 | rs17437810 | A | G | 0.021 | 0.077 | (0.013) | 8.0E-09 | 0.98 | 8.5E-07 | 0.96 | 0.55 | yes |
| 7p15.2 | 27483166 | rs10250340 | A | G | 0.321 | 0.023 | (0.004) | 2.0E-09 | 0.047 | 1.4E-08 | 0.073 | 0.030 | yes |
| 7p15.2 | 27975919 | rs67152137 | G | C | 0.740 | 0.048 | (0.004) | 8.6E-31 | 0.29 | 1.9E-28 | 0.13 | 0.78 | no |
| 8p21.2 | 23437981 | rs2928681 | C | A | 0.413 | 0.031 | (0.004) | 3.5E-18 | 0.10 | 5.7E-17 | 0.045 | 0.66 | no |
| 8p21.2 | 23529521 | rs1160267 | G | A | 0.430 | 0.070 | (0.004) | 6.3E-83 | 0.89 | 9.6E-78 | 0.78 | 0.69 | no |
| 8q12.1 | 57138676 | rs36112366 | T | G | 0.886 | 0.037 | (0.006) | 7.3E-11 | 0.73 | 6.9E-09 | 0.60 | 0.84 | yes |
| 8q12.1 | 57834357 | rs4738555 | C | T | 0.434 | 0.031 | (0.004) | 3.2E-17 | 0.22 | 2.1E-16 | 0.028 | 0.76 | no |
| 8q21.13 | 81811566 | 8:81811566_CA_C | CA | C | 0.590 | 0.024 | (0.004) | 7.6E-11 | 0.13 | 9.1E-10 | 0.090 | 0.54 | yes |
| 8q23.1 | 108849527 | rs1494920 | T | C | 0.604 | 0.020 | (0.004) | 4.8E-08 | 0.84 | 1.7E-06 | 0.46 | 0.55 | yes |
| 8q24.21 | 127817460 | rs7843726 | G | C | 0.145 | 0.030 | (0.005) | 1.7E-08 | 0.073 | 2.0E-07 | 0.14 | 0.24 | yes |
| 8q24.21 | 128093297 | rs1016343 | T | C | 0.205 | 0.035 | (0.004) | 3.6E-15 | 0.67 | 9.2E-13 | 0.94 | 0.80 | yes |
| 8q24.21 | 128318755 | rs10107982 | T | C | 0.728 | 0.046 | (0.004) | 4.1E-30 | 0.85 | 6.9E-27 | 0.98 | 0.25 | no |

| | | | | | | | | | | | | | |
|---|---|---|---|---|---|---|---|---|---|---|---|---|---|
| 8q24.21 | 128413305 | rs6983267 | G | T | 0.512 | 0.051 | (0.004) | 2.6E-45 | 0.49 | 7.1E-43 | 0.45 | 0.31 | no |
| 9q22.32 | 99146751 | rs7020681 | C | T | 0.214 | 0.027 | (0.005) | 1.4E-09 | 6.1E-03 | 8.2E-10 | 5.6E-03 | 0.74 | yes |
| 9q33.1 | 118258080 | rs150402584 | T | TG | 0.129 | 0.035 | (0.006) | 9.0E-10 | 0.14 | 1.6E-08 | 0.17 | 0.37 | yes |
| 9q33.1 | 120475302 | rs4986790 | A | G | 0.942 | 0.065 | (0.008) | 7.1E-17 | 0.51 | 6.7E-15 | 0.51 | 0.73 | no |
| 9q33.2 | 123643426 | rs59482735 | T | TAA | 0.321 | 0.032 | (0.005) | 2.9E-12 | 0.23 | 2.2E-10 | 0.44 | 0.087 | no |
| 10p12.31 | 22363979 | rs3011633 | C | T | 0.221 | 0.040 | (0.004) | 5.9E-20 | 0.71 | 1.1E-17 | 0.36 | 0.81 | no |
| 10p12.2 | 23398448 | rs1857279 | C | G | 0.662 | 0.025 | (0.004) | 3.9E-11 | 0.90 | 5.3E-09 | 0.71 | 0.41 | yes |
| 10p12.1 | 28096754 | rs2815506 | A | G | 0.804 | 0.036 | (0.005) | 4.9E-15 | 0.55 | 3.2E-13 | 0.50 | 0.44 | no |
| 10q11.23 | 51549496 | rs10993994 | T | C | 0.402 | 0.086 | (0.004) | 7.3E-87 | 0.16 | 3.0E-84 | 0.20 | 0.68 | no |
| 10q25.2 | 111991733 | rs562417881 | G | GT | 0.840 | 0.035 | (0.006) | 5.6E-09 | 0.21 | 2.5E-07 | 0.43 | 0.36 | yes |
| 10q26.12 | 122631067 | rs9325569 | A | G | 0.340 | 0.051 | (0.004) | 9.9E-41 | 0.40 | 1.2E-38 | 0.39 | 0.72 | no |
| 10q26.12 | 123049264 | rs10886902 | C | T | 0.231 | 0.099 | (0.004) | 8.2E-118 | 0.92 | 6.2E-116 | 0.83 | 0.58 | no |
| 10q26.13 | 123135200 | rs10788173 | C | T | 0.403 | 0.021 | (0.004) | 2.9E-08 | 0.27 | 4.5E-07 | 0.19 | 0.86 | yes |
| 10q26.13 | 123185303 | rs10749415 | A | G | 0.920 | 0.108 | (0.008) | 1.8E-45 | 0.72 | 1.7E-42 | 0.75 | 0.42 | no |
| 10q26.13 | 123349324 | rs45631563 | T | A | 0.045 | 0.089 | (0.009) | 7.9E-23 | 0.28 | 4.5E-20 | 0.97 | 0.082 | no |
| 10q26.13 | 126737579 | rs3012065 | C | T | 0.320 | 0.022 | (0.004) | 2.2E-08 | 0.70 | 1.6E-06 | 0.76 | 0.99 | yes |
| 11p15.5 | 197557 | rs7103852 | G | A | 0.878 | 0.041 | (0.007) | 1.2E-09 | 0.86 | 7.2E-08 | 0.64 | 0.60 | yes |
| 11p13 | 34718279 | rs553481604 | G | GT | 0.697 | 0.031 | (0.005) | 1.7E-11 | 0.59 | 5.4E-10 | 0.21 | 7.4E-03 | no |
| 11p13 | 34780936 | rs10466455 | C | T | 0.397 | 0.038 | (0.004) | 8.0E-26 | 0.19 | 2.0E-23 | 0.20 | 0.80 | no |
| 11q13.5 | 76153279 | rs58015965 | G | A | 0.688 | 0.021 | (0.004) | 4.0E-08 | 0.84 | 1.9E-06 | 0.49 | 0.78 | yes |
| 11q22.2 | 102396607 | rs12285347 | C | T | 0.452 | 0.038 | (0.004) | 2.6E-25 | 0.17 | 5.5E-24 | 0.13 | 0.98 | no |
| 11q22.2 | 102438778 | rs7129424 | A | G | 0.559 | 0.024 | (0.004) | 2.5E-11 | 0.42 | 1.8E-09 | 0.54 | 0.52 | yes |
| 12p13.31 | 9222286 | rs1805664 | T | C | 0.334 | 0.022 | (0.004) | 9.2E-09 | 0.12 | 1.1E-07 | 0.18 | 0.96 | yes |
| 12q14.3 | 66393756 | rs74097857 | T | C | 0.872 | 0.033 | (0.006) | 1.2E-09 | 0.75 | 8.7E-08 | 0.54 | 0.24 | yes |
| 12q21.31 | 84994000 | rs61928422 | T | C | 0.169 | 0.027 | (0.005) | 1.1E-08 | 0.027 | 9.6E-09 | 0.015 | 0.27 | yes |
| 12q24.21 | 115099805 | rs10774760 | G | A | 0.504 | 0.042 | (0.004) | 2.4E-30 | 0.98 | 3.3E-27 | 0.84 | 0.78 | no |
| 12q24.21 | 115155460 | rs11067251 | G | T | 0.584 | 0.022 | (0.004) | 1.9E-09 | 0.10 | 1.4E-08 | 0.077 | 0.96 | yes |
| 12q24.23 | 118855432 | rs1045542 | G | A | 0.907 | 0.047 | (0.006) | 1.2E-13 | 0.69 | 1.8E-11 | 0.76 | 0.47 | yes |
| 13q14.3 | 51087443 | rs202346 | A | C | 0.253 | 0.044 | (0.004) | 1.8E-27 | 0.38 | 4.0E-26 | 0.025 | 0.53 | no |
| 13q14.3 | 51165912 | rs12869529 | A | G | 0.960 | 0.064 | (0.009) | 9.4E-12 | 0.80 | 9.6E-10 | 0.69 | 0.031 | yes |
| 13q14.3 | 51445360 | rs9563006 | A | G | 0.443 | 0.024 | (0.004) | 5.6E-11 | 0.81 | 1.9E-10 | 0.034 | 0.78 | no |

| | | | | | | | | | | | | | |
|---|---|---|---|---|---|---|---|---|---|---|---|---|---|
| 13q22.1 | 74055303 | rs1361057 | C | T | 0.261 | 0.023 | (0.004) | 3.3E-08 | 0.30 | 1.6E-06 | 0.53 | 0.43 | yes |
| 13q32.1 | 95799006 | rs61965887 | A | G | 0.062 | 0.059 | (0.008) | 3.7E-14 | 0.74 | 2.5E-12 | 0.51 | 0.57 | yes |
| 14q13.3 | 37240591 | rs712329 | G | A | 0.432 | 0.021 | (0.004) | 2.0E-08 | 0.031 | 1.2E-07 | 0.083 | 0.22 | yes |
| 14q32.13 | 94838142 | rs112635299 | G | T | 0.979 | 0.106 | (0.013) | 1.2E-15 | 0.26 | 3.4E-14 | 0.19 | 0.36 | yes |
| 14q32.13 | 95104745 | rs72695000 | C | T | 0.825 | 0.054 | (0.005) | 7.4E-28 | 0.76 | 4.5E-26 | 0.19 | 0.12 | no |
| 15q22.31 | 66941399 | rs8038722 | G | A | 0.499 | 0.020 | (0.004) | 3.2E-08 | 0.68 | 3.9E-06 | 1.00 | 0.65 | yes |
| 16p13.3 | 4318206 | rs251739 | G | A | 0.506 | 0.033 | (0.004) | 6.6E-20 | 0.13 | 2.0E-17 | 0.46 | 0.13 | no |
| 16p13.13 | 11201428 | rs9933507 | C | T | 0.430 | 0.023 | (0.004) | 3.8E-10 | 0.84 | 5.2E-08 | 0.90 | 0.74 | yes |
| 16q23.1 | 77507275 | rs7206309 | C | T | 0.316 | 0.039 | (0.004) | 1.6E-22 | 0.53 | 3.5E-20 | 0.54 | 0.56 | no |
| 16q24.1 | 86554784 | rs13334022 | T | C | 0.109 | 0.042 | (0.006) | 3.4E-11 | 0.17 | 5.7E-10 | 0.14 | 0.45 | yes |
| 17p13.3 | 1994071 | rs3760230 | C | G | 0.433 | 0.020 | (0.004) | 2.8E-08 | 0.63 | 1.7E-06 | 0.58 | 0.56 | yes |
| 17p13.1 | 7571752 | rs78378222 | G | T | 0.012 | 0.112 | (0.018) | 2.8E-10 | 0.90 | 3.6E-08 | 0.86 | 0.85 | yes |
| 17p13.1 | 7793367 | rs34009884 | C | CTG | 0.104 | 0.035 | (0.006) | 1.1E-08 | 0.20 | 1.7E-07 | 0.26 | 0.29 | yes |
| 17p12 | 12320741 | rs7219536 | T | C | 0.495 | 0.025 | (0.004) | 5.7E-12 | 0.23 | 2.0E-10 | 0.30 | 0.60 | yes |
| 17p12 | 12556260 | rs9889266 | C | A | 0.574 | 0.023 | (0.004) | 8.9E-10 | 0.62 | 4.4E-08 | 0.65 | 0.97 | yes |
| 17q12 | 36099952 | rs10908278 | A | T | 0.525 | 0.052 | (0.004) | 2.1E-46 | 0.017 | 2.8E-44 | 0.16 | 0.017 | no |
| 18q23 | 76777252 | rs71279357 | A | T | 0.725 | 0.035 | (0.005) | 1.8E-12 | 0.80 | 1.3E-10 | 0.57 | 0.97 | yes |
| 19q12 | 32055024 | rs11084590 | C | T | 0.691 | 0.035 | (0.004) | 3.4E-18 | 0.39 | 2.4E-16 | 0.48 | 0.91 | no |
| 19q13.32 | 46229711 | rs6509234 | G | T | 0.822 | 0.028 | (0.005) | 7.7E-09 | 0.066 | 1.0E-07 | 0.18 | 0.28 | yes |
| 19q13.33 | 49206462 | rs681343 | T | C | 0.483 | 0.020 | (0.004) | 4.4E-08 | 0.70 | 2.4E-06 | 0.65 | 0.70 | yes |
| 19q13.33 | 51225544 | rs890863 | G | A | 0.656 | 0.023 | (0.004) | 1.1E-08 | 0.44 | 3.1E-07 | 0.43 | 0.22 | yes |
| 19q13.33 | 51323501 | rs1054713 | A | G | 0.324 | 0.037 | (0.004) | 4.8E-22 | 5.9E-04 | 2.1E-22 | 8.7E-04 | 0.076 | no |
| 19q13.33 | 51340794 | rs2659053 | A | G | 0.405 | 0.078 | (0.004) | 5.4E-94 | 0.035 | 2.3E-92 | 5.2E-03 | 0.031 | no |
| 19q13.33 | 51349841 | rs113920094 | C | T | 0.007 | 0.144 | (0.025) | 6.5E-09 | 0.072 | 3.4E-08 | 0.076 | 0.98 | yes |
| 19q13.33 | 51360398 | rs374546878 | GA | G | 0.762 | 0.165 | (0.023) | 1.7E-12 | 0.12 | - | - | - | yes |
| 19q13.33 | 51361757 | rs17632542 | T | C | 0.924 | 0.368 | (0.007) | 3.2e-638 | 0.21 | 7.0e-641 | 2.8E-15 | 4.4E-09 | no |
| 19q13.33 | 51365466 | rs2735837 | G | A | 0.871 | 0.098 | (0.006) | 7.2E-58 | 1.5E-12 | 6.7E-67 | 3.6E-12 | 0.51 | no |
| 19q13.33 | 51373744 | rs12983994 | T | C | 0.050 | 0.071 | (0.008) | 1.1E-17 | 1.3E-03 | 3.2E-19 | 2.0E-04 | 0.32 | no |
| 19q13.33 | 51378275 | rs112103380 | C | G | 0.008 | 0.212 | (0.024) | 1.7E-18 | 0.89 | 5.3E-16 | 0.88 | 0.38 | yes |
| 19q13.33 | 51383072 | rs198978 | G | T | 0.642 | 0.044 | (0.004) | 8.4E-31 | 1.5E-03 | 5.1E-30 | 0.034 | 0.077 | yes |
| 19q13.41 | 51421334 | rs141135092 | C | G | 0.976 | 0.091 | (0.015) | 1.2E-09 | 0.11 | 1.9E-09 | 0.025 | 0.43 | yes |

| | | | | | | | | | | | | | |
|---|---|---|---|---|---|---|---|---|---|---|---|---|---|
| 19q13.41 | 51441425 | rs80122351 | G | T | 0.804 | 0.030 | (0.005) | 1.2E-10 | 0.37 | 5.2E-09 | 0.56 | 7.5E-03 | no |
| 20q11.21 | 31950845 | rs291671 | G | A | 0.114 | 0.053 | (0.006) | 1.2E-18 | 0.23 | 1.3E-16 | 0.31 | 3.1E-03 | yes |
| 21q22.2 | 40289167 | rs2836750 | T | G | 0.401 | 0.027 | (0.004) | 1.6E-12 | 0.46 | 4.3E-11 | 0.22 | 0.97 | yes |
| 22q13.31 | 45996298 | rs13268 | A | G | 0.976 | 0.080 | (0.013) | 1.6E-10 | 0.45 | 1.4E-08 | 0.69 | 0.43 | yes |
| Xp22.31 | 8912815 | rs112062538 | C | T | 0.261 | 0.029 | (0.003) | 4.7E-19 | 0.57 | 5.8E-18 | 0.10 | 0.71 | yes |
| Xp22.2 | 16844831 | rs62586587 | G | A | 0.614 | 0.031 | (0.003) | 3.6E-25 | 0.56 | 1.9E-23 | 0.054 | 0.027 | no |
| Xp22.13 | 17831188 | rs200615037 | TTG | T | 0.950 | 0.066 | (0.007) | 3.5E-22 | 0.87 | 2.8E-20 | 0.34 | 0.021 | no |
| Xp22.11 | 24109619 | rs7065158 | A | G | 0.311 | 0.036 | (0.003) | 1.1E-29 | 0.080 | 1.6E-27 | 0.099 | 0.25 | no |
| Xp11.22 | 51242364 | rs5987424 | A | G | 0.384 | 0.017 | (0.003) | 2.1E-08 | 0.16 | 3.4E-07 | 0.15 | 0.41 | yes |
| Xp11.21 | 55936822 | rs10855058 | A | G | 0.322 | 0.019 | (0.003) | 7.6E-09 | 0.054 | 2.5E-08 | 0.048 | 0.20 | yes |
| Xq13.1 | 68996186 | rs2520386 | G | A | 0.735 | 0.023 | (0.003) | 4.2E-11 | 0.64 | 1.3E-09 | 0.18 | 0.50 | yes |
| Xq13.1 | 70125103 | rs62608084 | A | G | 0.334 | 0.032 | (0.003) | 1.7E-24 | 0.43 | 4.8E-25 | 3.8E-03 | 0.089 | yes |
| Xq26.2 | 132935739 | rs4829762 | T | C | 0.704 | 0.024 | (0.003) | 5.9E-12 | 0.59 | 3.7E-10 | 0.35 | 0.90 | yes |
| Xq26.3 | 133703924 | rs5930651 | T | C | 0.665 | 0.017 | (0.003) | 1.0E-08 | 0.62 | 4.3E-07 | 0.43 | 0.99 | yes |

[1] Per allele change in log(PSA)

[2] Variants that were independent of previously reported PSA associations at LD $r^2$<0.01 within ±10 Mb were considered novel

Abbreviations:

EAF Effect allele frequency

Note: MR-MEGA results are not available for variants that were present in too few studies

**Supplementary Table 4: Population-specific association estimates for 128 index variants associated with PSA in the multi-ancestry meta-analysis (N=95,768).** For each variant, effect sizes (β) and corresponding two-sided p-values are reported from an inverse-variance-weighted fixed-effects meta-analysis performed within each ancestry group using METAL.

| Region | Position (b37) | Variant ID | Effect Allele | Other Allele | EUR (N=85,824) | | | AFR (N=3,509) | | | HIS/LAT (N=3,098) | | | EAS (N=3,337) | | | New[2] |
|---|---|---|---|---|---|---|---|---|---|---|---|---|---|---|---|---|---|
| | | | | | EAF | β[1] | $P_{METAL}$ | EAF | Effect | $P_{METAL}$ | EAF | β[1] | $P_{METAL}$ | EAF | β[1] | $P_{METAL}$ | |
| 1p32.3 | 51237409 | rs12569177 | C | G | 0.431 | 0.027 | 2.78E-12 | 0.565 | 0.023 | 0.21 | 0.362 | 0.036 | 0.088 | 0.215 | 0.011 | 0.62 | yes |
| 1p22.3 | 88199778 | rs12131120 | T | A | 0.478 | 0.033 | 2.13E-18 | 0.862 | 0.018 | 0.50 | 0.565 | 0.025 | 0.24 | 0.923 | 0.089 | 0.011 | no |
| 1p13.2 | 112264274 | rs2076591 | C | T | 0.207 | 0.028 | 1.45E-09 | 0.398 | 0.050 | 8.07E-03 | 0.177 | 0.064 | 0.019 | 0.184 | 0.078 | 1.58E-03 | yes |
| 1q21.3 | 154868055 | rs4845681 | T | G | 0.393 | 0.023 | 1.84E-09 | 0.533 | 0.014 | 0.45 | 0.535 | -0.024 | 0.24 | 0.852 | 0.023 | 0.39 | yes |
| 1q25.3 | 184191716 | rs12046452 | C | T | 0.622 | 0.021 | 4.05E-08 | 0.797 | 0.026 | 0.26 | 0.639 | 0.030 | 0.17 | 0.503 | 0.013 | 0.50 | yes |
| 1q32.1 | 205632217 | rs71152447 | G | GGCCGACA GCCCTTCT GCTGGCTC GGTGGGG CCCAGC | 0.013 | 0.112 | 5.05E-03 | 0.010 | 0.071 | 0.65 | 0.051 | 0.228 | 3.42E-06 | 0.078 | 0.176 | 5.46E-06 | no |
| 1q32.1 | 205636334 | rs4951018 | C | A | 0.224 | 0.033 | 5.03E-13 | 0.132 | 0.007 | 0.82 | 0.321 | 0.036 | 0.099 | 0.340 | 0.008 | 0.70 | no |
| 1q41 | 219694903 | rs12034581 | C | A | 0.477 | 0.025 | 6.82E-11 | 0.821 | 0.013 | 0.60 | 0.622 | 0.005 | 0.80 | 0.655 | 0.073 | 2.49E-04 | yes |
| 1q41 | 219954267 | rs6664688 | G | A | 0.185 | 0.019 | 7.72E-05 | 0.393 | 0.041 | 0.028 | 0.277 | 0.073 | 1.36E-03 | 0.454 | 0.057 | 2.89E-03 | yes |
| 2p24.2 | 18911674 | rs12710685 | G | C | 0.098 | 0.037 | 6.92E-09 | 0.135 | 0.008 | 0.76 | 0.118 | -0.019 | 0.55 | 0.178 | 0.026 | 0.31 | yes |
| 2p24.1 | 20880833 | rs10193919 | T | C | 0.347 | 0.031 | 2.07E-14 | 0.118 | 0.010 | 0.73 | 0.327 | 0.043 | 0.047 | 0.437 | 0.029 | 0.13 | yes |
| 2p23.3 | 25463871 | rs734693 | T | C | 0.745 | 0.023 | 2.68E-07 | 0.504 | -0.015 | 0.42 | 0.691 | 0.059 | 8.65E-03 | 0.352 | 0.042 | 0.041 | yes |
| 2p21 | 43072828 | rs1123695 | G | C | 0.662 | 0.030 | 4.07E-14 | 0.824 | 0.031 | 0.19 | 0.725 | 0.048 | 0.036 | 0.442 | 0.043 | 0.026 | yes |
| 2p21 | 43618819 | rs11899863 | C | T | 0.900 | 0.044 | 6.10E-12 | 0.910 | 0.075 | 0.021 | 0.935 | 0.070 | 0.086 | 0.995 | -0.097 | 0.46 | yes |
| 2p16.1 | 60759747 | rs2556375 | G | T | 0.172 | 0.058 | 8.88E-30 | 0.172 | 0.058 | 0.020 | 0.221 | 0.080 | 1.36E-03 | 0.221 | 0.019 | 0.43 | no |
| 2p15 | 63277843 | rs58235267 | G | C | 0.476 | 0.029 | 6.68E-14 | 0.453 | -0.022 | 0.24 | 0.512 | -0.009 | 0.67 | 0.679 | 0.045 | 0.030 | yes |
| 2p13.2 | 72059504 | rs11679946 | T | C | 0.680 | 0.022 | 4.67E-08 | 0.764 | 0.024 | 0.28 | 0.718 | -0.021 | 0.37 | 0.431 | 0.040 | 0.039 | yes |
| 2q33.1 | 198929083 | rs34388051 | A | G | 0.679 | 0.021 | 3.04E-07 | 0.820 | 0.042 | 0.10 | 0.594 | 0.008 | 0.71 | 0.470 | 0.038 | 0.052 | yes |
| 3q13.2 | 113290793 | rs4682495 | A | C | 0.580 | 0.024 | 5.95E-10 | 0.571 | 0.012 | 0.52 | 0.612 | -0.024 | 0.24 | 0.339 | 0.008 | 0.69 | no |
| 3q21.3 | 128067275 | rs11709611 | C | T | 0.116 | 0.036 | 1.25E-09 | 0.192 | 0.036 | 0.12 | 0.109 | 0.042 | 0.19 | 0.011 | -0.013 | 0.90 | yes |
| 3q23 | 141115219 | rs1582874 | C | T | 0.440 | 0.031 | 2.00E-16 | 0.796 | 0.042 | 0.063 | 0.405 | 0.013 | 0.52 | 0.233 | -0.003 | 0.90 | no |
| 3q25.33 | 160130592 | rs10673842 | TTATC | T | 0.530 | 0.025 | 2.41E-08 | 0.579 | -0.019 | 0.37 | 0.460 | 0.048 | 0.028 | 0.271 | 0.001 | 0.97 | yes |
| 3q27.2 | 185246981 | rs4012714 | A | G | 0.574 | 0.028 | 8.53E-10 | 0.560 | -0.018 | 0.43 | 0.589 | 0.016 | 0.51 | 0.730 | -0.009 | 0.73 | yes |

| | | | | | | | | | | | | | | | | | |
|---|---|---|---|---|---|---|---|---|---|---|---|---|---|---|---|---|---|
| 3q27.3 | 186746994 | rs12629450 | C | G | 0.169 | 0.032 | 3.92E-10 | 0.139 | 0.020 | 0.48 | 0.185 | 0.034 | 0.23 | 0.231 | 0.003 | 0.91 | yes |
| 3q28 | 189482770 | rs6765473 | T | C | 0.881 | 0.034 | 9.06E-09 | 0.967 | 0.117 | 0.040 | 0.845 | 0.070 | 0.017 | 0.986 | 0.086 | 0.45 | yes |
| 4p15.2 | 24410024 | rs4276269 | G | A | 0.265 | 0.023 | 8.32E-08 | 0.255 | 0.010 | 0.65 | 0.255 | 0.031 | 0.19 | 0.222 | 0.028 | 0.22 | yes |
| 4q31.22 | 146890261 | rs9968429 | G | A | 0.361 | 0.030 | 2.25E-14 | 0.438 | 0.078 | 2.49E-05 | 0.358 | 0.014 | 0.51 | 0.506 | 0.039 | 0.037 | no |
| 4q32.1 | 157497921 | rs13115840 | A | G | 0.355 | 0.031 | 3.01E-15 | 0.172 | 0.051 | 0.040 | 0.363 | 0.013 | 0.54 | 0.457 | 0.035 | 0.063 | no |
| 5p15.33 | 1285974 | rs7705526 | A | C | 0.334 | 0.024 | 5.30E-08 | 0.200 | -0.008 | 0.74 | 0.309 | 0.023 | 0.30 | 0.386 | 0.045 | 0.052 | no |
| 5p15.33 | 1322087 | rs401681 | C | T | 0.562 | 0.059 | 2.19E-49 | 0.417 | 0.078 | 2.35E-05 | 0.581 | 0.053 | 9.58E-03 | 0.672 | 0.033 | 0.13 | no |
| 5q14.3 | 92052005 | rs7732515 | A | T | 0.224 | 0.034 | 2.45E-13 | 0.331 | 0.045 | 0.023 | 0.184 | 0.004 | 0.89 | 0.020 | 0.099 | 0.23 | yes |
| 5q35.2 | 172945918 | rs889017 | C | A | 0.213 | 0.022 | 1.59E-06 | 0.550 | 0.020 | 0.28 | 0.189 | 0.029 | 0.27 | 0.162 | 0.091 | 5.38E-04 | yes |
| 6p22.3 | 15463139 | rs926309 | T | G | 0.385 | 0.027 | 4.98E-12 | 0.275 | 0.037 | 0.080 | 0.290 | 0.012 | 0.60 | 0.187 | 0.014 | 0.56 | yes |
| 6p22.3 | 16515154 | rs236961 | T | C | 0.347 | 0.028 | 5.18E-12 | 0.415 | -0.020 | 0.29 | 0.442 | 0.025 | 0.23 | 0.367 | 0.035 | 0.079 | yes |
| 6p22.1 | 29915061 | rs2248162 | C | T | 0.659 | 0.025 | 1.14E-09 | 0.700 | 0.022 | 0.27 | 0.782 | 0.042 | 0.092 | 0.956 | -0.061 | 0.19 | yes |
| 6p21.31 | 34212537 | rs551980123 | TG | T | 0.044 | 0.067 | 8.96E-10 | 0.023 | 0.017 | 0.81 | 0.019 | 0.240 | 2.96E-03 | 0.015 | 0.004 | 0.97 | yes |
| 6p21.1 | 43711981 | rs1535507 | T | C | 0.156 | 0.051 | 6.93E-22 | 0.067 | -0.016 | 0.66 | 0.155 | -0.005 | 0.85 | 0.325 | 0.059 | 4.24E-03 | no |
| 6q22.33 | 129168057 | rs73583119 | A | T | 6E-04 | - | - | 0.926 | 0.181 | 3.98E-07 | 0.995 | 0.452 | 7.66E-03 | 1E-04 | - | - | yes |
| 7p15.3 | 20909622 | rs204592 | G | A | 0.532 | 0.020 | 1.47E-07 | 0.269 | 0.027 | 0.19 | 0.493 | 0.041 | 0.041 | 0.350 | 0.056 | 4.82E-03 | yes |
| 7p15.3 | 21839896 | rs6956349 | T | C | 0.438 | 0.021 | 4.41E-08 | 0.117 | 0.018 | 0.53 | 0.298 | 0.029 | 0.18 | 0.027 | 0.077 | 0.18 | yes |
| 7p15.2 | 27281792 | rs17437810 | A | G | 0.021 | 0.077 | 1.21E-08 | 0.006 | 0.045 | 0.80 | 0.015 | 0.086 | 0.33 | NA | NA | NA | yes |
| 7p15.2 | 27483166 | rs10250340 | A | G | 0.323 | 0.024 | 3.00E-09 | 0.353 | 0.053 | 6.51E-03 | 0.284 | -0.011 | 0.63 | 0.277 | -0.017 | 0.44 | yes |
| 7p15.2 | 27975919 | rs67152137 | G | C | 0.764 | 0.049 | 2.56E-28 | 0.707 | 0.011 | 0.60 | 0.601 | 0.051 | 0.012 | 0.154 | 0.063 | 0.017 | no |
| 8p21.2 | 23437981 | rs2928681 | C | A | 0.418 | 0.033 | 4.82E-18 | 0.443 | -0.009 | 0.64 | 0.338 | 0.012 | 0.59 | 0.282 | 0.048 | 0.024 | no |
| 8p21.2 | 23529521 | rs1160267 | G | A | 0.424 | 0.069 | 2.53E-72 | 0.696 | 0.085 | 2.55E-05 | 0.448 | 0.070 | 9.02E-04 | 0.315 | 0.073 | 4.22E-04 | no |
| 8q12.1 | 57138676 | rs36112366 | T | G | 0.884 | 0.038 | 2.64E-10 | 0.971 | 0.028 | 0.61 | 0.894 | 0.058 | 0.080 | 0.914 | 0.005 | 0.89 | yes |
| 8q12.1 | 57834357 | rs4738555 | C | T | 0.445 | 0.032 | 1.28E-16 | 0.325 | 0.045 | 0.023 | 0.401 | 0.021 | 0.31 | 0.223 | -0.013 | 0.56 | no |
| 8q21.13 | 81811566 | 8:81811566_CA_C | CA | C | 0.588 | 0.022 | 2.25E-08 | 0.732 | 0.059 | 4.97E-03 | 0.468 | 0.015 | 0.45 | 0.619 | 0.053 | 6.25E-03 | yes |
| 8q23.1 | 108849527 | rs1494920 | T | C | 0.619 | 0.019 | 1.02E-06 | 0.513 | 0.035 | 0.059 | 0.569 | 0.027 | 0.20 | 0.372 | 0.024 | 0.23 | yes |
| 8q24.21 | 127817460 | rs7843726 | G | C | 0.139 | 0.031 | 2.98E-08 | 0.036 | -0.097 | 0.074 | 0.126 | 0.063 | 0.047 | 0.261 | 0.015 | 0.50 | yes |
| 8q24.21 | 128093297 | rs1016343 | T | C | 0.201 | 0.035 | 4.99E-14 | 0.215 | 0.047 | 0.036 | 0.155 | 0.004 | 0.89 | 0.289 | 0.032 | 0.12 | yes |
| 8q24.21 | 128318755 | rs10107982 | T | C | 0.720 | 0.046 | 5.80E-28 | 0.874 | 0.055 | 0.049 | 0.790 | 0.031 | 0.22 | 0.908 | 0.066 | 0.048 | no |
| 8q24.21 | 128413305 | rs6983267 | G | T | 0.508 | 0.052 | 1.49E-42 | 0.865 | 0.037 | 0.17 | 0.569 | 0.071 | 5.77E-04 | 0.384 | 0.030 | 0.12 | no |

| | | | | | | | | | | | | | | | | |
|---|---|---|---|---|---|---|---|---|---|---|---|---|---|---|---|---|
| 9q22.32 | 99146751 | rs7020681 | C | T | 0.196 | 0.032 | 1.93E-11 | 0.543 | -0.026 | 0.17 | 0.166 | 0.002 | 0.95 | 0.041 | -0.050 | 0.30 | yes |
| 9q33.1 | 118258080 | rs150402584 | T | TG | 0.100 | 0.037 | 3.91E-09 | 0.163 | 0.003 | 0.90 | 0.133 | -0.014 | 0.63 | 0.381 | 0.054 | 5.67E-03 | yes |
| 9q33.1 | 120475302 | rs4986790 | A | G | 0.942 | 0.068 | 4.77E-17 | 0.925 | 0.015 | 0.66 | 0.958 | 0.053 | 0.29 | 0.992 | 0.110 | 0.37 | no |
| 9q33.2 | 123643426 | rs59482735 | T | TAA | 0.326 | 0.029 | 1.86E-09 | 0.250 | 0.038 | 0.14 | 0.296 | 0.073 | 2.22E-03 | 0.263 | 0.057 | 0.026 | no |
| 10p12.31 | 22363979 | rs3011633 | C | T | 0.224 | 0.041 | 1.81E-19 | 0.061 | 0.018 | 0.65 | 0.212 | 0.016 | 0.52 | 0.198 | 0.033 | 0.18 | no |
| 10p12.2 | 23398448 | rs1857279 | C | G | 0.672 | 0.024 | 1.70E-09 | 0.708 | 0.028 | 0.18 | 0.639 | 0.041 | 0.054 | 0.393 | 0.026 | 0.18 | yes |
| 10p12.1 | 28096754 | rs2815506 | A | G | 0.823 | 0.036 | 2.13E-13 | 0.718 | 0.059 | 3.78E-03 | 0.810 | 0.021 | 0.43 | 0.596 | 0.023 | 0.24 | no |
| 10q11.23 | 51549496 | rs10993994 | T | C | 0.391 | 0.085 | 2.66E-76 | 0.600 | 0.065 | 3.12E-03 | 0.377 | 0.077 | 6.52E-04 | 0.493 | 0.132 | 6.74E-09 | no |
| 10q25.2 | 111991733 | rs562417881 | G | GT | 0.856 | 0.038 | 3.19E-09 | 0.934 | 0.082 | 0.094 | 0.712 | -0.004 | 0.87 | 0.706 | 0.013 | 0.61 | yes |
| 10q26.12 | 122631067 | rs9325569 | A | G | 0.339 | 0.053 | 4.42E-40 | 0.147 | 0.014 | 0.59 | 0.420 | 0.040 | 0.049 | 0.413 | 0.038 | 0.051 | no |
| 10q26.12 | 123049264 | rs10886902 | C | T | 0.231 | 0.099 | 6.5E-110 | 0.064 | 0.100 | 9.36E-03 | 0.247 | 0.087 | 2.33E-04 | 0.264 | 0.109 | 6.06E-07 | no |
| 10q26.13 | 123135200 | rs10788173 | C | T | 0.397 | 0.023 | 9.00E-09 | 0.580 | 0.017 | 0.43 | 0.407 | 0.003 | 0.88 | 0.391 | -0.015 | 0.47 | yes |
| 10q26.13 | 123185303 | rs10749415 | A | G | 0.944 | 0.104 | 2.93E-34 | 0.789 | 0.130 | 1.02E-08 | 0.850 | 0.115 | 6.13E-05 | 0.969 | 0.130 | 0.049 | no |
| 10q26.13 | 123349324 | rs45631563 | T | A | 0.046 | 0.089 | 2.66E-22 | 0.013 | 0.120 | 0.20 | 0.042 | 0.022 | 0.68 | 0.009 | 0.293 | 0.038 | no |
| 10q26.13 | 126737579 | rs3012065 | C | T | 0.321 | 0.022 | 1.28E-07 | 0.331 | 0.024 | 0.24 | 0.319 | 0.004 | 0.85 | 0.262 | 0.041 | 0.066 | yes |
| 11p15.5 | 197557 | rs7103852 | G | A | 0.914 | 0.041 | 2.10E-08 | 0.597 | 0.043 | 0.029 | 0.912 | 0.022 | 0.56 | 0.959 | 0.093 | 0.22 | yes |
| 11p13 | 34718279 | rs553481604 | G | GT | 0.703 | 0.030 | 1.27E-09 | 0.741 | 0.032 | 0.20 | 0.626 | 0.061 | 7.30E-03 | 0.612 | 0.022 | 0.35 | no |
| 11p13 | 34780936 | rs10466455 | C | T | 0.400 | 0.040 | 9.24E-26 | 0.387 | 0.004 | 0.84 | 0.305 | 0.024 | 0.27 | 0.380 | 0.023 | 0.24 | no |
| 11q13.5 | 76153279 | rs58015965 | G | A | 0.679 | 0.021 | 1.18E-07 | 0.785 | 0.011 | 0.61 | 0.771 | 0.036 | 0.15 | 0.871 | 0.005 | 0.85 | yes |
| 11q22.2 | 102396607 | rs12285347 | C | T | 0.455 | 0.039 | 6.15E-25 | 0.552 | -0.002 | 0.90 | 0.373 | 0.050 | 0.016 | 0.095 | 0.053 | 0.12 | no |
| 11q22.2 | 102438778 | rs7129424 | A | G | 0.547 | 0.023 | 3.16E-09 | 0.735 | 0.031 | 0.13 | 0.560 | 0.050 | 0.015 | 0.779 | 0.046 | 0.043 | yes |
| 12p13.31 | 9222286 | rs1805664 | T | C | 0.341 | 0.020 | 9.17E-07 | 0.202 | 0.044 | 0.056 | 0.340 | 0.057 | 7.18E-03 | 0.089 | 0.067 | 0.047 | yes |
| 12q14.3 | 66393756 | rs74097857 | T | C | 0.870 | 0.033 | 3.59E-09 | 0.957 | 0.065 | 0.16 | 0.912 | 0.012 | 0.74 | 0.995 | 0.126 | 0.41 | yes |
| 12q21.31 | 84994000 | rs61928422 | T | C | 0.166 | 0.027 | 1.50E-07 | 0.203 | 0.005 | 0.84 | 0.224 | 0.006 | 0.80 | 0.151 | 0.100 | 1.76E-04 | yes |
| 12q24.21 | 115099805 | rs10774760 | G | A | 0.498 | 0.042 | 1.50E-27 | 0.811 | 0.034 | 0.15 | 0.477 | 0.039 | 0.057 | 0.462 | 0.047 | 0.017 | no |
| 12q24.21 | 115155460 | rs11067251 | G | T | 0.573 | 0.022 | 6.83E-09 | 0.816 | -0.019 | 0.43 | 0.565 | 0.019 | 0.37 | 0.864 | 0.074 | 8.65E-03 | yes |
| 12q24.23 | 118855432 | rs1045542 | G | A | 0.907 | 0.049 | 7.35E-14 | 0.981 | -0.003 | 0.97 | 0.887 | 0.023 | 0.48 | 0.991 | -0.027 | 0.85 | yes |
| 13q14.3 | 51087443 | rs202346 | A | C | 0.254 | 0.042 | 2.07E-22 | 0.296 | 0.070 | 4.36E-04 | 0.233 | 0.051 | 0.033 | 0.146 | 0.073 | 6.78E-03 | no |
| 13q14.3 | 51165912 | rs12869529 | A | G | 0.961 | 0.065 | 5.11E-11 | 0.944 | 0.079 | 0.053 | 0.953 | 0.032 | 0.51 | 0.998 | -0.110 | 0.67 | yes |
| 13q14.3 | 51445360 | rs9563006 | A | G | 0.443 | 0.024 | 2.58E-10 | 0.506 | 0.012 | 0.53 | 0.436 | 0.027 | 0.19 | 0.054 | 0.054 | 0.21 | no |
| 13q22.1 | 74055303 | rs1361057 | C | T | 0.254 | 0.022 | 6.42E-07 | 0.225 | 0.029 | 0.20 | 0.249 | -0.003 | 0.89 | 0.430 | 0.052 | 6.87E-03 | yes |

| | | | | | | | | | | | | | | | | | |
|---|---|---|---|---|---|---|---|---|---|---|---|---|---|---|---|---|---|
| 13q32.1 | 95799006 | rs61965887 | A | G | 0.063 | 0.060 | 1.94E-14 | 0.016 | 0.010 | 0.90 | 0.035 | 0.035 | 0.53 | 0.004 | -0.114 | 0.58 | yes |
| 14q13.3 | 37240591 | rs712329 | G | A | 0.444 | 0.020 | 1.69E-07 | 0.324 | 0.010 | 0.60 | 0.329 | 0.002 | 0.92 | 0.138 | 0.101 | 4.41E-04 | yes |
| 14q32.13 | 94838142 | rs112635299 | G | T | 0.979 | 0.103 | 7.78E-15 | 0.997 | 0.423 | 0.076 | 0.990 | 0.211 | 0.056 | 8E-04 | - | - | yes |
| 14q32.13 | 95104745 | rs72695000 | C | T | 0.821 | 0.054 | 4.10E-27 | 0.943 | 0.030 | 0.46 | 0.887 | 0.067 | 0.033 | 0.982 | -0.011 | 0.89 | no |
| 15q22.31 | 66941399 | rs8038722 | G | A | 0.502 | 0.019 | 3.55E-07 | 0.337 | 0.016 | 0.42 | 0.465 | 0.044 | 0.032 | 0.632 | 0.017 | 0.39 | yes |
| 16p13.3 | 4318206 | rs251739 | G | A | 0.506 | 0.033 | 2.67E-17 | 0.358 | 0.048 | 0.013 | 0.488 | 0.063 | 1.86E-03 | 0.719 | -0.006 | 0.81 | no |
| 16p13.13 | 11201428 | rs9933507 | C | T | 0.431 | 0.022 | 6.58E-09 | 0.519 | 0.026 | 0.16 | 0.374 | 0.015 | 0.46 | 0.344 | 0.039 | 0.052 | yes |
| 16q23.1 | 77507275 | rs7206309 | C | T | 0.286 | 0.041 | 5.52E-22 | 0.445 | 0.018 | 0.35 | 0.480 | 0.034 | 0.093 | 0.761 | 0.021 | 0.36 | no |
| 16q24.1 | 86554784 | rs13334022 | T | C | 0.089 | 0.039 | 1.04E-08 | 0.307 | 0.076 | 2.11E-04 | 0.097 | 0.043 | 0.20 | 0.048 | -0.031 | 0.54 | yes |
| 17p13.3 | 1994071 | rs3760230 | C | G | 0.423 | 0.020 | 3.79E-07 | 0.518 | 0.021 | 0.27 | 0.363 | 0.012 | 0.56 | 0.725 | 0.047 | 0.030 | yes |
| 17p13.1 | 7571752 | rs78378222 | G | T | 0.012 | 0.112 | 4.08E-10 | 0.001 | - | - | 0.004 | 0.133 | 0.44 | 8E-04 | - | - | yes |
| 17p13.1 | 7793367 | rs34009884 | C | CTG | 0.106 | 0.038 | 1.49E-09 | 0.025 | -0.056 | 0.37 | 0.057 | -0.029 | 0.52 | 0.011 | -0.035 | 0.79 | yes |
| 17p12 | 12320741 | rs7219536 | T | C | 0.509 | 0.024 | 3.05E-10 | 0.262 | 0.048 | 0.023 | 0.461 | 0.055 | 7.92E-03 | 0.094 | -0.006 | 0.84 | yes |
| 17p12 | 12556260 | rs9889266 | C | A | 0.590 | 0.024 | 8.26E-10 | 0.489 | 0.027 | 0.14 | 0.430 | 0.011 | 0.61 | 0.381 | 0.000 | 0.98 | yes |
| 17q12 | 36099952 | rs10908278 | A | T | 0.513 | 0.051 | 9.85E-42 | 0.688 | 0.008 | 0.71 | 0.552 | 0.102 | 8.41E-07 | 0.713 | 0.058 | 6.32E-03 | no |
| 18q23 | 76777252 | rs71279357 | A | T | 0.729 | 0.035 | 1.31E-11 | 0.769 | 0.012 | 0.64 | 0.733 | 0.047 | 0.062 | 0.602 | 0.031 | 0.20 | yes |
| 19q12 | 32055024 | rs11084590 | C | T | 0.682 | 0.036 | 6.87E-19 | 0.894 | -0.013 | 0.68 | 0.775 | 0.020 | 0.41 | 0.926 | 0.020 | 0.60 | no |
| 19q13.32 | 46229711 | rs6509234 | G | T | 0.817 | 0.029 | 1.18E-08 | 0.940 | 0.056 | 0.18 | 0.866 | 0.060 | 0.047 | 0.897 | -0.050 | 0.13 | yes |
| 19q13.33 | 49206462 | rs681343 | T | C | 0.488 | 0.019 | 5.94E-07 | 0.489 | 0.036 | 0.051 | 0.380 | 0.033 | 0.11 | 0.024 | -0.008 | 0.90 | yes |
| 19q13.33 | 51225544 | rs890863 | G | A | 0.663 | 0.023 | 3.92E-08 | 0.879 | 0.062 | 0.043 | 0.632 | 0.001 | 0.96 | 0.389 | 0.026 | 0.22 | yes |
| 19q13.33 | 51323501 | rs1054713 | A | G | 0.330 | 0.040 | 1.38E-22 | 0.256 | 0.003 | 0.90 | 0.275 | 0.070 | 2.37E-03 | 0.226 | -0.046 | 0.050 | no |
| 19q13.33 | 51340794 | rs2659053 | A | G | 0.391 | 0.082 | 4.35E-92 | 0.449 | 0.042 | 0.026 | 0.547 | 0.049 | 0.018 | 0.615 | 0.046 | 0.049 | no |
| 19q13.33 | 51349841 | rs113920094 | C | T | 0.007 | 0.144 | 1.28E-08 | 0.002 | 1.008 | 0.011 | 0.009 | 0.038 | 0.79 | 0.001 | - | - | yes |
| 19q13.33 | 51360398 | rs374546878 | GA | G | 0.999 | 0.150 | 0.17 | 0.988 | -0.089 | 0.42 | 0.992 | 0.215 | 0.082 | 0.728 | 0.177 | 1.58E-12 | yes |
| 19q13.33 | 51361757 | rs17632542 | T | C | 0.923 | 0.366 | 1.1E-605 | 0.982 | 0.341 | 3.30E-06 | 0.954 | 0.463 | 3.83E-22 | 0.994 | 0.267 | 0.083 | no |
| 19q13.33 | 51365466 | rs2735837 | G | A | 0.915 | 0.080 | 5.26E-31 | 0.830 | 0.074 | 3.20E-03 | 0.827 | 0.158 | 3.13E-09 | 0.579 | 0.227 | 4.73E-32 | no |
| 19q13.33 | 51373744 | rs12983994 | T | C | 0.048 | 0.069 | 6.53E-15 | 0.071 | 0.015 | 0.68 | 0.046 | 0.030 | 0.54 | 0.056 | 0.216 | 1.39E-07 | no |
| 19q13.33 | 51378275 | rs112103380 | C | G | 0.008 | 0.210 | 7.27E-18 | 0.002 | 0.244 | 0.49 | 0.005 | 0.299 | 0.10 | 7E-04 | - | - | yes |
| 19q13.33 | 51383072 | rs198978 | G | T | 0.650 | 0.040 | 1.01E-23 | 0.362 | 0.045 | 0.020 | 0.646 | 0.108 | 4.47E-07 | 0.759 | 0.095 | 1.64E-05 | yes |
| 19q13.41 | 51421334 | rs141135092 | C | G | 0.976 | 0.091 | 1.67E-09 | 0.995 | -0.341 | 0.092 | 0.984 | 0.211 | 0.042 | 0.996 | -0.013 | 0.95 | yes |
| 19q13.41 | 51441425 | rs80122351 | G | T | 0.800 | 0.031 | 6.08E-11 | 0.938 | 0.016 | 0.70 | 0.863 | -0.002 | 0.95 | 0.989 | 0.194 | 0.11 | no |

| | | | | | | | | | | | | | | | | | |
|---|---|---|---|---|---|---|---|---|---|---|---|---|---|---|---|---|---|
| 20q11.21 | 31950845 | rs291671 | G | A | 0.100 | 0.049 | 4.46E-14 | 0.048 | 0.093 | 0.028 | 0.077 | 0.046 | 0.23 | 0.299 | 0.089 | 2.04E-05 | yes |
| 21q22.2 | 40289167 | rs2836750 | T | G | 0.380 | 0.026 | 1.51E-10 | 0.524 | 0.053 | 5.27E-03 | 0.548 | 0.018 | 0.40 | 0.774 | 0.041 | 0.080 | yes |
| 22q13.31 | 45996298 | rs13268 | A | G | 0.975 | 0.078 | 8.33E-10 | 0.992 | 0.173 | 0.13 | 0.986 | 0.158 | 0.099 | 0.999 | -0.368 | 0.35 | yes |
| Xp22.31 | 8912815 | rs112062538 | C | T | 0.263 | 0.028 | 4.40E-17 | 0.271 | 0.034 | 0.041 | 0.185 | 0.052 | 7.38E-03 | 0.247 | 0.017 | 0.34 | yes |
| Xp22.2 | 16844831 | rs62586587 | G | A | 0.631 | 0.030 | 5.80E-21 | 0.544 | 0.051 | 7.33E-04 | 0.470 | 0.033 | 0.041 | 0.252 | 0.037 | 0.067 | no |
| Xp22.13 | 17831188 | rs200615037 | TTG | T | 0.954 | 0.068 | 3.69E-21 | 0.897 | 0.055 | 0.027 | 0.960 | 0.039 | 0.33 | 0.989 | 0.074 | 0.62 | no |
| Xp22.11 | 24109619 | rs7065158 | A | G | 0.318 | 0.037 | 6.75E-28 | 0.138 | 0.040 | 0.15 | 0.250 | 0.003 | 0.85 | 0.150 | 0.086 | 1.32E-03 | no |
| Xp11.22 | 51242364 | rs5987424 | A | G | 0.394 | 0.018 | 5.99E-09 | 0.287 | 0.010 | 0.54 | 0.267 | -0.009 | 0.62 | 0.132 | -0.029 | 0.30 | yes |
| Xp11.21 | 55936822 | rs10855058 | A | G | 0.317 | 0.017 | 7.13E-07 | 0.310 | 0.061 | 3.17E-04 | 0.448 | 0.028 | 0.071 | 0.304 | 0.004 | 0.83 | yes |
| Xq13.1 | 68996186 | rs2520386 | G | A | 0.730 | 0.022 | 8.09E-10 | 0.911 | 0.048 | 0.20 | 0.820 | 0.037 | 0.069 | 0.979 | 0.067 | 0.22 | yes |
| Xq13.1 | 70125103 | rs62608084 | A | G | 0.342 | 0.033 | 4.91E-25 | 0.195 | 0.017 | 0.35 | 0.212 | 0.009 | 0.64 | 0.019 | 0.076 | 0.20 | yes |
| Xq26.2 | 132935739 | rs4829762 | T | C | 0.725 | 0.025 | 1.63E-11 | 0.522 | 0.026 | 0.092 | 0.795 | 0.007 | 0.73 | 0.462 | 0.008 | 0.58 | yes |
| Xq26.3 | 133703924 | rs5930651 | T | C | 0.679 | 0.018 | 3.97E-08 | 0.292 | -0.004 | 0.84 | 0.610 | 0.017 | 0.31 | 0.653 | 0.028 | 0.087 | yes |

[1] Per allele change in log(PSA)

[2] Variants that were independent of previously reported PSA associations at LD $r^2$<0.01 within ±10 Mb were considered novel

Abbreviations:

EAF Effect allele frequency

**Supplementary Table 5: Association estimates for 119 index variants associated with PSA levels detected with MR-MEGA.** Genome-wide significant ($P<5\times10^{-8}$) index variants were selected from the multi-ancestry meta-analysis performed with MR-MEGA using LD-based clumping at $r^2<0.01$ within ± 10Mb windows. MR-MEGA is an alternative approach to cross-ancestry meta-analysis that uses meta-regression to quantify heterogeneity due to ancestry ($P_{Het\text{-}Anc}$) separately from residual heterogeneity due to other factors ($P_{Het\text{-}Res}$). All p-values are two-sided. For comparison, association p-values based on the fixed-effects multi-ancestry meta-analysis using METAL ($P_{METAL}$) are provided.

| Region | Position (b37) | Variant ID | Effect Allele | Other Allele | Meta-Regression (MR-MEGA) | | | | | | | Meta-analysis |
|---|---|---|---|---|---|---|---|---|---|---|---|---|
| | | | | | $EAF_{MEGA}$ | $P_{MEGA}$ | $P_{Het\text{-}Anc}$ | $P_{Het\text{-}Res}$ | N | $N_{study}$ | Direction | $P_{METAL}$ |
| 1p32.3 | 51021867 | rs147696085 | A | G | 0.088 | 2.62E-09 | 0.51 | 0.16 | 95768 | 12 | -++-++++++++ | 1.75E-11 |
| 1p32.3 | 51480258 | rs7555006 | G | A | 0.439 | 7.11E-11 | 0.25 | 0.68 | 95768 | 12 | -++++--+++++ | 6.64E-13 |
| 1p22.3 | 88182718 | rs10493801 | G | A | 0.511 | 4.41E-20 | 0.016 | 0.16 | 69277 | 11 | +---?-+--+-- | 6.11E-21 |
| 1p13.2 | 112264274 | rs2076591 | C | T | 0.212 | 1.86E-11 | 0.18 | 0.70 | 95768 | 12 | +++++++++++<br>+ | 2.64E-13 |
| 1q32.1 | 205632217 | rs71152447 | GGCCGACAGC<br>CCTTCTGCTG<br>GCTCGGTGGG<br>GCCCAGC | G | 0.980 | 4.24E-10 | 0.19 | 0.11 | 44597 | 8 | ?-?-?+?----- | 9.91E-12 |
| 1q32.1 | 205636334 | rs4951018 | C | A | 0.227 | 7.05E-12 | 0.081 | 0.077 | 95768 | 12 | +++++--+++++ | 2.75E-13 |
| 1q41 | 219694903 | rs12034581 | A | C | 0.500 | 1.38E-10 | 0.24 | 0.39 | 95768 | 12 | +----+--+--- | 2.46E-12 |
| 2p24.1 | 20878153 | rs9306895 | C | T | 0.353 | 4.15E-13 | 0.67 | 0.32 | 95768 | 12 | +++++-++++++ | 1.79E-15 |
| 2p23.3 | 25463871 | rs734693 | T | C | 0.720 | 4.72E-08 | 0.018 | 0.66 | 95768 | 12 | +++++--+++++ | 3.88E-08 |
| 2p21 | 43072828 | rs1123695 | C | G | 0.338 | 2.87E-13 | 0.95 | 0.73 | 95768 | 12 | ---------+-- | 3.25E-16 |
| 2p21 | 43618819 | rs11899863 | T | C | 0.095 | 8.79E-11 | 0.71 | 0.050 | 95768 | 12 | +---------++ | 1.72E-13 |
| 2p16.1 | 60759747 | rs2556375 | T | G | 0.825 | 1.67E-29 | 0.47 | 0.45 | 95768 | 12 | ---------+-- | 1.75E-32 |
| 2p16.1 | 60790985 | rs1510478 | A | G | 0.161 | 2.82E-13 | 0.22 | 0.47 | 95768 | 12 | +------+---- | 2.22E-15 |
| 2p15 | 63277843 | rs58235267 | G | C | 0.483 | 2.88E-12 | 0.024 | 0.56 | 95768 | 12 | -++++-----++ | 4.92E-13 |
| 3q23 | 141115219 | rs1582874 | C | T | 0.444 | 5.11E-14 | 0.53 | 0.70 | 95768 | 12 | +++++++++--+ | 1.01E-16 |
| 3q27.3 | 186763665 | rs59663889 | A | T | 0.194 | 4.31E-08 | 0.45 | 0.19 | 95768 | 12 | +++++++-++-- | 4.50E-10 |
| 3q28 | 189482770 | rs6765473 | C | T | 0.114 | 1.26E-08 | 0.25 | 0.78 | 94949 | 11 | +----------? | 3.25E-10 |
| 3q28 | 191387768 | rs114388981 | T | A | 0.983 | 4.40E-08 | 1.29E-08 | 0.14 | 38447 | 8 | ?+???++-++-- | 0.78 |
| 4q31.22 | 146890261 | rs9968429 | G | A | 0.369 | 2.12E-16 | 0.053 | 0.34 | 95768 | 12 | +++++++++-++ | 6.34E-18 |
| 4q32.1 | 157491375 | rs17034927 | T | C | 0.356 | 8.03E-14 | 0.96 | 0.46 | 95768 | 12 | +++++++++++<br>+ | 1.22E-16 |
| 5p15.33 | 1287194 | rs2853677 | A | G | 0.584 | 2.29E-22 | 0.56 | 0.87 | 95768 | 12 | ------+----- | 7.91E-25 |

| | | | | | | | | | | | | |
|---|---|---|---|---|---|---|---|---|---|---|---|---|
| 5p15.33 | 1322087 | rs401681 | T | C | 0.437 | 1.17E-51 | 0.20 | 0.78 | 95768 | 12 | ------------ | 6.98E-54 |
| 5p15.33 | 1366007 | rs152133 | A | G | 0.368 | 1.21E-12 | 0.19 | 0.65 | 95768 | 12 | +++++-+-++-+ | 8.33E-15 |
| 5q14.3 | 92052005 | rs7732515 | A | T | 0.221 | 7.56E-12 | 0.56 | 0.62 | 94949 | 11 | ++++++++-++? | 3.53E-14 |
| 5q15 | 96727294 | rs291812 | T | C | 0.468 | 1.00E-08 | 2.24E-04 | 0.99 | 95768 | 12 | +++++---++++ | 9.30E-07 |
| 6p22.3 | 15230743 | rs112266013 | A | G | 0.132 | 4.90E-10 | 6.20E-04 | 0.88 | 95768 | 12 | ------------ | 9.85E-09 |
| 6p22.3 | 16515154 | rs236961 | T | C | 0.353 | 1.84E-10 | 0.12 | 0.43 | 95768 | 12 | +++++--+++++ | 8.57E-12 |
| 6p22.1 | 29915061 | rs2248162 | C | T | 0.675 | 2.08E-08 | 0.26 | 0.36 | 95768 | 12 | -++++++-+--+ | 3.60E-10 |
| 6p21.31 | 34237109 | rs575339229 | C | G | 0.976 | 1.64E-09 | 0.040 | 0.57 | 69887 | 7 | ?-?---?--?-? | 2.79E-10 |
| 6p21.1 | 43710348 | rs6920449 | C | T | 0.839 | 1.63E-22 | 1.29E-03 | 0.054 | 95768 | 12 | ------++++-- | 8.05E-22 |
| 7p15.3 | 20909622 | rs204592 | G | A | 0.515 | 3.82E-08 | 0.22 | 0.47 | 95768 | 12 | +++++++-++++ | 1.18E-09 |
| 7p15.2 | 27483166 | rs10250340 | A | G | 0.321 | 4.66E-08 | 0.13 | 0.018 | 95768 | 12 | +++++++-+--- | 2.03E-09 |
| 7p15.2 | 27975919 | rs67152137 | C | G | 0.264 | 8.12E-28 | 0.16 | 0.78 | 95768 | 12 | ------++---- | 8.57E-31 |
| 8p21.2 | 23470583 | rs7004327 | A | G | 0.440 | 1.25E-18 | 0.70 | 0.27 | 95768 | 12 | -----+-----+ | 7.77E-22 |
| 8p21.2 | 23529521 | rs1160267 | A | G | 0.569 | 9.76E-77 | 0.89 | 0.58 | 95768 | 12 | ------------ | 6.29E-83 |
| 8q12.1 | 57132317 | rs145612921 | G | GC | 0.108 | 1.79E-08 | 0.62 | 0.73 | 94949 | 11 | ------+----? | 1.18E-10 |
| 8q12.1 | 57834357 | rs4738555 | T | C | 0.569 | 9.74E-16 | 0.058 | 0.67 | 95768 | 12 | ---------++- | 3.20E-17 |
| 8q21.13 | 81811566 | 8:81811566_CA_C | C | CA | 0.409 | 3.37E-09 | 0.16 | 0.44 | 95768 | 12 | +--------+-+ | 7.64E-11 |
| 8q24.21 | 128093297 | rs1016343 | T | C | 0.203 | 1.90E-12 | 0.75 | 0.88 | 95768 | 12 | ++++++++++++ | 3.59E-15 |
| 8q24.21 | 128228847 | rs74631227 | G | A | 0.040 | 1.87E-09 | 0.055 | 0.029 | 94949 | 11 | ------+--++? | 2.02E-10 |
| 8q24.21 | 128318755 | rs10107982 | C | T | 0.266 | 1.94E-27 | 0.17 | 0.78 | 95768 | 12 | ---------+-- | 4.07E-30 |
| 8q24.21 | 128413305 | rs6983267 | T | G | 0.481 | 5.42E-42 | 0.62 | 0.22 | 95768 | 12 | ------+----- | 2.55E-45 |
| 8q24.21 | 128455167 | rs28645077 | T | A | 0.354 | 4.50E-09 | 0.64 | 0.60 | 95768 | 12 | ++++++-+++++ | 1.23E-11 |
| 9q22.32 | 99146751 | rs7020681 | C | T | 0.203 | 2.71E-09 | 0.012 | 0.68 | 95768 | 12 | +++++----+-- | 1.40E-09 |
| 9q33.1 | 118258187 | rs9409165 | G | A | 0.119 | 1.57E-08 | 0.050 | 0.55 | 95768 | 12 | +++++-+---++ | 2.46E-09 |
| 9q33.1 | 120473180 | rs10983756 | T | C | 0.057 | 1.46E-15 | 2.30E-03 | 0.88 | 94949 | 11 | +----+++---? | 3.55E-15 |
| 9q33.2 | 123642351 | rs1609810 | T | C | 0.658 | 1.41E-10 | 0.10 | 0.24 | 95768 | 12 | +--+-----+-- | 8.89E-12 |
| 10p12.31 | 21855753 | rs141268071 | G | T | 0.019 | 1.97E-08 | 0.011 | 0.63 | 91496 | 9 | ++++++?+--?? | 2.23E-08 |
| 10p12.31 | 22308015 | rs10828285 | A | G | 0.713 | 3.76E-12 | 0.53 | 0.96 | 95768 | 12 | ------+----- | 2.21E-14 |
| 10p12.31 | 22363979 | rs3011633 | C | T | 0.217 | 3.96E-17 | 0.43 | 0.80 | 95768 | 12 | ++++++-++++- | 5.92E-20 |
| 10p12.31 | 22590653 | rs79800006 | A | T | 0.022 | 2.56E-11 | 2.08E-03 | 0.032 | 93960 | 10 | +++++++?+--? | 1.01E-10 |

| | | | | | | | | | | | | |
|---|---|---|---|---|---|---|---|---|---|---|---|---|
| 10p12.1 | 28096754 | rs2815506 | A | G | 0.811 | 8.67E-13 | 0.50 | 0.44 | 95768 | 12 | +++++++++-++ | 4.88E-15 |
| 10q11.23 | 51549496 | rs10993994 | T | C | 0.402 | 2.31E-83 | 0.26 | 0.66 | 69887 | 7 | ?+?+++?++?+? | 7.31E-87 |
| 10q11.23 | 51551900 | rs57592785 | C | CG | 0.846 | 1.67E-08 | 0.23 | 0.57 | 69887 | 7 | ?+?+++?++?+? | 5.00E-10 |
| 10q26.12 | 122667022 | rs1530116 | G | T | 0.337 | 7.75E-38 | 0.47 | 0.65 | 95768 | 12 | +++++++-++++ | 1.07E-40 |
| 10q26.12 | 122926366 | rs59421178 | A | G | 0.163 | 2.64E-08 | 3.58E-03 | 0.12 | 95768 | 12 | +++-+---+++- | 8.51E-08 |
| 10q26.12 | 122933134 | rs12354663 | T | C | 0.098 | 1.00E-14 | 0.44 | 0.79 | 95768 | 12 | ++++++++++-- | 2.87E-17 |
| 10q26.12 | 123009023 | rs144869102 | A | G | 0.015 | 1.91E-09 | 0.43 | 0.91 | 94014 | 10 | ++++++?---+? | 1.35E-11 |
| 10q26.12 | 123025352 | rs61872992 | C | A | 0.422 | 1.16E-21 | 0.93 | 0.51 | 95768 | 12 | ------+----- | 3.15E-25 |
| 10q26.12 | 123049264 | rs10886902 | C | T | 0.227 | 5.53E-115 | 0.82 | 0.54 | 95768 | 12 | ++++++++++++ | 8.16E-118 |
| 10q26.12 | 123095366 | rs7914065 | T | C | 0.253 | 8.09E-24 | 4.69E-03 | 0.46 | 95768 | 12 | ++++++--++-+ | 5.01E-24 |
| 10q26.13 | 123185303 | rs10749415 | A | G | 0.937 | 1.28E-41 | 0.87 | 0.32 | 95768 | 12 | +++++++++++- | 1.83E-45 |
| 10q26.13 | 123349324 | rs45631563 | T | A | 0.043 | 3.42E-20 | 0.37 | 0.20 | 94949 | 11 | +++++++-+-+? | 7.94E-23 |
| 11p13 | 34676842 | rs896856 | T | G | 0.475 | 6.99E-18 | 0.33 | 0.059 | 95768 | 12 | -+++++-++++- | 2.34E-20 |
| 11p13 | 34780936 | rs10466455 | C | T | 0.396 | 1.10E-22 | 0.33 | 0.71 | 95768 | 12 | ++++++-++-++ | 8.02E-26 |
| 11q22.2 | 102396607 | rs12285347 | C | T | 0.443 | 1.68E-23 | 0.14 | 1.00 | 95768 | 12 | +++++++-++++ | 2.61E-25 |
| 11q22.2 | 102438778 | rs7129424 | A | G | 0.563 | 3.67E-09 | 0.50 | 0.54 | 95768 | 12 | +++++++++++- | 2.50E-11 |
| 12q21.31 | 84994000 | rs61928422 | T | C | 0.169 | 3.34E-08 | 0.031 | 0.20 | 95768 | 12 | -++++-++-+++ | 1.11E-08 |
| 12q24.21 | 115094260 | rs11067228 | G | A | 0.441 | 2.69E-27 | 0.19 | 0.99 | 95768 | 12 | ------------ | 1.25E-29 |
| 12q24.21 | 115136391 | rs1463888 | G | T | 0.337 | 7.38E-14 | 1.43E-03 | 0.79 | 95768 | 12 | -----+++---- | 2.66E-13 |
| 12q24.23 | 118601003 | rs35506792 | C | T | 0.093 | 3.12E-11 | 0.081 | 0.70 | 94949 | 11 | +----+++--+? | 2.01E-12 |
| 13q14.3 | 51087443 | rs202346 | A | C | 0.251 | 1.37E-25 | 0.034 | 0.54 | 95768 | 12 | ++++++++++++ | 1.82E-27 |
| 13q14.3 | 51165912 | rs12869529 | G | A | 0.039 | 3.42E-09 | 0.81 | 0.018 | 94949 | 11 | ---+---+-++? | 9.40E-12 |
| 13q14.3 | 51194405 | rs573666 | T | C | 0.340 | 3.17E-17 | 0.16 | 0.12 | 95768 | 12 | -+++++--+++- | 8.44E-19 |
| 13q14.3 | 51446114 | rs12429206 | G | A | 0.651 | 2.04E-10 | 1.53E-03 | 0.48 | 95768 | 12 | ------++-+-+ | 2.01E-09 |
| 13q32.1 | 95799006 | rs61965887 | A | G | 0.059 | 7.89E-12 | 0.58 | 0.53 | 94949 | 11 | ++++++--++-? | 3.69E-14 |
| 14q13.3 | 37240591 | rs712329 | A | G | 0.575 | 4.42E-08 | 0.022 | 0.56 | 95768 | 12 | ---+-+--+--- | 2.01E-08 |
| 14q32.13 | 94838142 | rs112635299 | T | G | 0.020 | 9.68E-14 | 0.23 | 0.33 | 90125 | 7 | ------??-??? | 1.21E-15 |
| 14q32.13 | 95097556 | rs8023057 | G | A | 0.166 | 1.00E-28 | 0.19 | 0.062 | 68458 | 10 | ----?-++--+? | 3.23E-31 |
| 14q32.13 | 95138671 | rs12891726 | C | T | 0.571 | 3.81E-11 | 0.63 | 0.38 | 95768 | 12 | ++++++-+++++ | 9.05E-14 |
| 16p13.3 | 4304905 | rs2386890 | T | A | 0.334 | 1.05E-10 | 0.52 | 0.59 | 95768 | 12 | -++++++++-++ | 3.11E-13 |

| | | | | | | | | | | | | |
|---|---|---|---|---|---|---|---|---|---|---|---|---|
| 16p13.3 | 4318206 | rs251739 | G | A | 0.507 | 7.24E-18 | 0.092 | 0.41 | 95768 | 12 | -++++++++--+ | 6.63E-20 |
| 16q23.1 | 77320605 | rs12921392 | G | A | 0.590 | 1.52E-08 | 0.42 | 0.90 | 95768 | 12 | ------------ | 1.89E-10 |
| 16q23.1 | 77507275 | rs7206309 | T | C | 0.685 | 1.44E-19 | 0.61 | 0.51 | 95768 | 12 | -------+---- | 1.61E-22 |
| 16q24.1 | 86547001 | rs56404456 | T | C | 0.350 | 1.43E-09 | 0.58 | 0.071 | 69277 | 11 | ++++?+++++-+ | 5.93E-12 |
| 17p12 | 12296124 | rs28444256 | G | C | 0.122 | 1.70E-08 | 1.42E-03 | 0.53 | 95768 | 12 | -+++++++++-+ | 2.10E-07 |
| 17p12 | 12323258 | rs2721828 | A | G | 0.502 | 4.33E-11 | 0.017 | 0.74 | 95768 | 12 | ----------++ | 1.81E-11 |
| 17q12 | 36099840 | rs11651755 | C | T | 0.491 | 6.54E-44 | 0.025 | 0.016 | 95768 | 12 | -----++----- | 3.94E-45 |
| 18q23 | 76777252 | rs71279357 | A | T | 0.726 | 4.66E-10 | 0.71 | 0.94 | 69887 | 7 | ?+?+++?++?+? | 1.83E-12 |
| 19q12 | 32059290 | rs7252125 | T | C | 0.339 | 3.18E-13 | 0.013 | 0.79 | 95768 | 12 | -----+++-++- | 1.74E-13 |
| 19q12 | 32104979 | rs11084596 | C | T | 0.386 | 1.14E-25 | 0.52 | 0.92 | 69277 | 11 | ----?-+----- | 4.17E-29 |
| 19q13.32 | 46229711 | rs6509234 | G | T | 0.826 | 2.22E-08 | 0.030 | 0.79 | 95768 | 12 | -++++++-++-- | 7.70E-09 |
| 19q13.33 | 51264558 | rs183959820 | A | C | 0.021 | 1.13E-38 | 1.37E-03 | 1.78E-04 | 91442 | 9 | ------+?-+?? | 1.16E-38 |
| 19q13.33 | 51280907 | rs1122124 | G | C | 0.420 | 1.08E-11 | 0.24 | 0.057 | 95768 | 12 | +++++-+-++-+ | 1.63E-13 |
| 19q13.33 | 51326106 | rs2659058 | T | C | 0.681 | 4.63E-27 | 1.66E-04 | 0.028 | 95768 | 12 | -----+-+--++ | 1.28E-25 |
| 19q13.33 | 51349090 | rs266849 | A | G | 0.794 | 7.39e-316 | 1.44E-05 | 0.035 | 95768 | 12 | ++++++++++++ | 5.70e-319 |
| 19q13.33 | 51349781 | rs112343212 | T | C | 0.026 | 2.09E-28 | 0.026 | 0.72 | 93960 | 10 | +++++++?+-+? | 1.07E-29 |
| 19q13.33 | 51356929 | rs193199179 | T | C | 0.008 | 3.34E-12 | 0.039 | 0.62 | 92643 | 8 | ++++++??+?-? | 3.86E-13 |
| 19q13.33 | 51361382 | rs61752561 | A | G | 0.037 | 7.51E-51 | 0.35 | 0.18 | 94949 | 11 | ----------+? | 2.12E-54 |
| 19q13.33 | 51361757 | rs17632542 | C | T | 0.072 | 4.1e-641 | 1.02E-16 | 1.82E-07 | 94949 | 11 | -----------? | 3.15E-638 |
| 19q13.33 | 51364649 | rs17632611 | C | A | 0.065 | 9.10E-63 | 0.59 | 0.88 | 95768 | 12 | ++++++++++++ | 1.05E-66 |
| 19q13.33 | 51383072 | rs198978 | T | G | 0.357 | 2.77E-29 | 0.062 | 0.054 | 95768 | 12 | ------------ | 8.38E-31 |
| 19q13.33 | 51399309 | rs112035754 | G | A | 0.103 | 1.26E-17 | 0.060 | 0.022 | 95768 | 12 | -------+---+ | 3.79E-19 |
| 19q13.41 | 51401797 | rs8104538 | G | A | 0.309 | 1.13E-08 | 0.16 | 0.84 | 95768 | 12 | -----++--++- | 7.21E-10 |
| 19q13.41 | 51440765 | rs2739403 | A | T | 0.179 | 1.02E-10 | 0.11 | 4.06E-03 | 94949 | 11 | +------+++-? | 2.53E-12 |
| 20q11.21 | 31950845 | rs291671 | A | G | 0.896 | 2.98E-16 | 0.28 | 2.87E-03 | 95768 | 12 | -------+-+-- | 1.19E-18 |
| 20q11.21 | 31966698 | rs2065703 | T | C | 0.146 | 3.76E-11 | 0.45 | 0.23 | 95768 | 12 | +--------+-- | 2.72E-13 |
| 20q11.22 | 32397556 | rs117119427 | T | C | 0.078 | 1.12E-10 | 0.14 | 0.040 | 94949 | 11 | -+++++++-++? | 4.00E-12 |
| 21q22.2 | 40289167 | rs2836750 | G | T | 0.595 | 1.73E-10 | 0.35 | 0.94 | 95768 | 12 | ------------ | 1.59E-12 |
| 22q13.31 | 45996298 | rs13268 | G | A | 0.023 | 3.82E-08 | 0.74 | 0.37 | 94949 | 11 | +---------+? | 1.63E-10 |

| | | | | | | | | | | | | |
|---|---|---|---|---|---|---|---|---|---|---|---|---|
| Xp22.31 | 8912815 | rs112062538 | C | T | 0.235 | 1.10E-17 | 0.086 | 0.84 | 95768 | 12 | ++++++++++++ | 4.74E-19 |
| Xp22.2 | 16864303 | rs5969728 | A | G | 0.378 | 1.01E-23 | 0.012 | 0.33 | 95768 | 12 | ---------+-- | 4.09E-24 |
| Xp22.13 | 17831188 | rs200615037 | T | TTG | 0.043 | 1.35E-19 | 0.47 | 0.012 | 94567 | 10 | ---------?-? | 3.54E-22 |
| Xp22.11 | 23996164 | rs2520225 | A | G | 0.538 | 9.67E-10 | 0.046 | 0.31 | 95768 | 12 | -----+--+--- | 1.98E-10 |
| Xp22.11 | 24109619 | rs7065158 | G | A | 0.635 | 9.49E-27 | 0.18 | 0.18 | 95768 | 12 | --------+--- | 1.11E-29 |
| Xq13.1 | 69025920 | rs2804355 | T | A | 0.612 | 1.74E-09 | 2.87E-03 | 0.52 | 95768 | 12 | +++++---++-- | 6.91E-09 |
| Xq13.1 | 69767264 | rs5936916 | T | A | 0.478 | 2.55E-08 | 0.12 | 0.48 | 95768 | 12 | ---------+-- | 2.75E-09 |
| Xq13.1 | 70125103 | rs62608084 | A | G | 0.293 | 2.84E-24 | 9.37E-03 | 0.054 | 94949 | 11 | +++++++++-+? | 1.66E-24 |
| Xq26.2 | 132942023 | rs1908817 | C | G | 0.665 | 5.49E-10 | 0.35 | 0.69 | 95768 | 12 | +++++++++-++ | 6.19E-12 |

Abbreviations:

$EAF_{MEGA}$ Average effect allele frequency weighted by the sample size of each input file

**Supplementary Table 6: Population-specific association estimates for variants that reached genome-wide significance using MR-MEGA but not METAL.** Out of the 119 independent index variants detected by MR-MEGA, four variants did not reach $P<5\times10^{-8}$ in the fixed effect multi-ancestry meta-analysis using METAL. For each variant, effect sizes (β) and corresponding two-sided p-values are reported from the inverse-variance-weighted fixed-effects meta-analysis performed within each ancestry group using METAL.

| Regional position | | Variant ID | Effect Allele | Other Allele | EUR (N=85,824) | | | AFR (N=3,509) | | | HIS/LAT (N=3,098) | | | EAS (N=3,337) | | | New[2] |
|---|---|---|---|---|---|---|---|---|---|---|---|---|---|---|---|---|---|
| | | | | | EAF | β[1] | $P_{METAL}$ | EAF | β[1] | $P_{METAL}$ | EAF | β[1] | $P_{METAL}$ | EAF | β[1] | $P_{METAL}$ | |
| 3q28 | 191387768 | rs114388981 | A | T | 0.003 | -0.031 | 0.62 | 0.126 | -0.019 | 0.49 | 0.011 | -0.453 | 3.9E-06 | 0.029 | 0.225 | 1.0E-04 | yes |
| 5q15 | 96727294 | rs291812 | T | C | 0.486 | 0.016 | 2.3E-05 | 0.133 | -0.026 | 0.35 | 0.474 | 0.010 | 0.64 | 0.350 | 0.097 | 1.2E-06 | yes |
| 10q26.12 | 122926366 | rs59421178 | A | G | 0.162 | 0.028 | 1.7E-07 | 0.159 | -0.045 | 0.080 | 0.170 | 0.020 | 0.47 | 0.210 | 0.067 | 5.5E-03 | yes |
| 17p12 | 12296124 | rs28444256 | C | G | 0.891 | -0.029 | 3.0E-06 | 0.697 | -0.034 | 0.095 | 0.859 | -0.108 | 2.4E-04 | 0.746 | 0.018 | 0.42 | yes |

1 Per allele change in log(PSA)

2 Variants that were independent of previously reported PSA associations, including those identified in the fixed-effects meta-analysis, at LD $r^2<0.01$ were considered novel

Abbreviations:

EAF Effect allele frequency

**Supplementary Table 8: Associations with prostate cancer risk for 128 index variants associated with PSA in the multi-ancestry meta-analysis.** Odds ratios (OR) and p-values for prostate cancer were obtained from summary statistics from the PRACTICAL consortium GWAS by Conti et al[32]. All p-values are two-sided and derived from a meta-analysis of European ancestry studies (85,554 cases and 91,972 controls) and multi-ancestry meta-analysis (107,247 cases and 127,006). For each variant, bias-adjusted effect sizes ($OR_{adj}$) and p-values ($P_{adj}$) were calculated using the Dudbridge method[33]. Two-sided bias-adjusted p-values were calculated from a chi-squared test statistic based on bias-corrected effect sizes and standard errors. For comparison, the effect of each variant on PSA levels is also presented, with two-sided p-values derived from the inverse-variance-weighted fixed-effect multi-ancestry meta-analysis of 95,768 men using METAL ($P_{METAL}$).

| Region | Position (b37) | Variant ID | PSA-increasing Allele | $P_{METAL}$ | Prostate Cancer (European Ancestry) | | | | Prostate Cancer (Multi-Ancestry) | | | |
|---|---|---|---|---|---|---|---|---|---|---|---|---|
| | | | | | Original GWAS | | Bias Corrected[1] | | Original GWAS | | Bias Corrected[2] | |
| | | | | | OR | P | $OR_{adj}$ | $P_{adj}$ | OR | P | $OR_{adj}$ | $P_{adj}$ |
| 1p32.3 | 51237409 | rs12569177 | C | 3.2E-13 | 1.04 | 3.6E-07 | 1.01 | 0.33 | 1.03 | 4.5E-05 | 1.00 | 0.87 |
| 1p22.3 | 88199778 | rs12131120 | T | 1.6E-19 | 1.04 | 1.7E-08 | 1.00 | 0.62 | 1.04 | 2.0E-09 | 1.01 | 0.49 |
| 1p13.2 | 112264274 | rs2076591 | C | 2.6E-13 | 1.02 | 0.012 | 0.99 | 0.23 | 1.02 | 0.031 | 0.98 | 0.056 |
| 1q21.3 | 154868055 | rs4845681 | T | 7.7E-09 | 1.05 | 2.5E-12 | 1.03 | 9.6E-04 | 1.05 | 8.5E-11 | 1.02 | 9.2E-03 |
| 1q25.3 | 184191716 | rs12046452 | C | 9.0E-09 | 1.04 | 3.2E-06 | 1.01 | 0.15 | 1.03 | 9.9E-06 | 1.01 | 0.36 |
| 1q32.1 | 205632217 | rs71152447 | G | 9.9E-12 | 1.00 | 0.96 | 0.83 | 6.2E-04 | 1.02 | 0.51 | 0.86 | 4.9E-04 |
| 1q32.1 | 205636334 | rs4951018 | C | 2.7E-13 | 1.05 | 6.2E-07 | 1.01 | 0.40 | 1.05 | 1.1E-10 | 1.02 | 0.096 |
| 1q41 | 219694903 | rs12034581 | C | 2.5E-12 | 1.00 | 0.91 | 0.97 | 1.1E-03 | 1.00 | 0.56 | 0.98 | 2.3E-03 |
| 1q41 | 219954267 | rs6664688 | G | 3.9E-08 | 0.97 | 5.9E-03 | 0.95 | 5.1E-07 | 0.99 | 0.26 | 0.96 | 1.0E-04 |
| 2p24.2 | 18911674 | rs12710685 | G | 2.3E-08 | 1.00 | 0.94 | 0.96 | 9.3E-03 | 1.01 | 0.39 | 0.97 | 0.025 |
| 2p24.1 | 20880833 | rs10193919 | T | 1.5E-15 | 1.07 | 1.4E-19 | 1.04 | 5.8E-05 | 1.08 | 4.3E-30 | 1.05 | 2.1E-08 |
| 2p23.3 | 25463871 | rs734693 | T | 3.9E-08 | 1.01 | 0.16 | 0.99 | 0.20 | 1.01 | 0.46 | 0.98 | 0.029 |
| 2p21 | 43072828 | rs1123695 | G | 3.3E-16 | 1.04 | 5.3E-08 | 1.01 | 0.42 | 1.04 | 1.4E-09 | 1.01 | 0.32 |
| 2p21 | 43618819 | rs11899863 | C | 1.7E-13 | 1.06 | 1.0E-05 | 1.01 | 0.72 | 1.05 | 9.9E-06 | 1.00 | 0.81 |
| 2p16.1 | 60759747 | rs2556375 | G | 1.8E-32 | 0.98 | 0.079 | 0.92 | 2.3E-12 | 0.98 | 0.029 | 0.92 | 4.3E-15 |
| 2p15 | 63277843 | rs58235267 | G | 4.9E-13 | 1.12 | 6.6E-54 | 1.09 | 9.8E-24 | 1.12 | 5.4E-72 | 1.09 | 8.5E-31 |
| 2p13.2 | 72059504 | rs11679946 | T | 2.6E-08 | 1.01 | 0.088 | 0.99 | 0.26 | 1.01 | 0.16 | 0.99 | 0.093 |
| 2q33.1 | 198929083 | rs34388051 | A | 2.2E-08 | 1.00 | 0.71 | 0.98 | 0.022 | 1.01 | 0.22 | 0.99 | 0.078 |
| 3q13.2 | 113290793 | rs4682495 | A | 2.9E-09 | 1.09 | 1.7E-28 | 1.06 | 5.7E-12 | 1.08 | 2.6E-28 | 1.05 | 1.7E-10 |
| 3q21.3 | 128067275 | rs11709611 | C | 2.0E-10 | 1.10 | 4.1E-18 | 1.06 | 1.2E-05 | 1.10 | 1.6E-19 | 1.05 | 1.2E-05 |
| 3q23 | 141115219 | rs1582874 | C | 1.0E-16 | 1.05 | 1.8E-09 | 1.01 | 0.21 | 1.04 | 1.6E-10 | 1.01 | 0.19 |

| | | | | | | | | | | | | |
|---|---|---|---|---|---|---|---|---|---|---|---|---|
| 3q25.33 | 160130592 | rs10673842 | TTATC | 2.2E-08 | 1.03 | 9.6E-06 | 1.01 | 0.46 | 1.03 | 6.2E-05 | 1.00 | 0.83 |
| 3q27.2 | 185246981 | rs4012714 | A | 1.1E-08 | 1.02 | 0.021 | 0.99 | 0.30 | 1.02 | 0.032 | 0.99 | 0.19 |
| 3q27.3 | 186746994 | rs12629450 | C | 2.6E-10 | 1.01 | 0.32 | 0.98 | 0.039 | 1.01 | 0.27 | 0.98 | 0.024 |
| 3q28 | 189482770 | rs6765473 | T | 3.2E-10 | 1.01 | 0.61 | 0.97 | 0.013 | 1.01 | 0.51 | 0.97 | 0.017 |
| 4p15.2 | 24410024 | rs4276269 | G | 2.0E-08 | 0.99 | 0.24 | 0.96 | 3.1E-04 | 0.98 | 0.041 | 0.96 | 4.7E-06 |
| 4q31.22 | 146890261 | rs9968429 | G | 6.3E-18 | 1.02 | 0.011 | 0.98 | 0.058 | 1.03 | 6.0E-05 | 0.99 | 0.28 |
| 4q32.1 | 157497921 | rs13115840 | A | 4.3E-17 | 1.01 | 0.061 | 0.98 | 0.018 | 1.02 | 5.9E-03 | 0.98 | 0.056 |
| 5p15.33 | 1285974 | rs7705526 | A | 1.2E-08 | 0.94 | 1.6E-12 | 0.92 | 5.3E-19 | 0.94 | 1.3E-16 | 0.92 | 1.5E-23 |
| 5p15.33 | 1322087 | rs401681 | C | 7.0E-54 | 1.04 | 4.0E-06 | 0.97 | 4.2E-04 | 1.02 | 0.011 | 0.95 | 1.9E-09 |
| 5q14.3 | 92052005 | rs7732515 | A | 3.5E-14 | 1.02 | 0.084 | 0.98 | 0.040 | 1.02 | 0.024 | 0.98 | 0.071 |
| 5q35.2 | 172945918 | rs889017 | C | 2.6E-08 | 1.04 | 6.0E-05 | 1.01 | 0.40 | 1.03 | 1.2E-04 | 1.00 | 0.72 |
| 6p22.3 | 15463139 | rs926309 | T | 1.6E-12 | 1.01 | 0.41 | 0.98 | 9.1E-03 | 1.01 | 0.13 | 0.98 | 0.024 |
| 6p22.3 | 16515154 | rs236961 | T | 8.6E-12 | 0.97 | 3.0E-05 | 0.94 | 5.3E-12 | 0.97 | 5.1E-06 | 0.94 | 1.1E-13 |
| 6p22.1 | 29915061 | rs2248162 | C | 3.6E-10 | 1.04 | 2.9E-06 | 1.01 | 0.35 | 1.03 | 3.9E-06 | 1.01 | 0.45 |
| 6p21.31 | 34212537 | rs551980123 | TG | 1.6E-10 | 1.01 | 0.74 | 0.93 | 1.4E-03 | 1.01 | 0.68 | 0.93 | 1.6E-03 |
| 6p21.1 | 43711981 | rs1535507 | T | 1.5E-22 | 1.05 | 4.8E-07 | 1.00 | 0.72 | 1.05 | 1.1E-07 | 0.99 | 0.50 |
| 6q22.33 | 129168057 | rs73583119 | A | 3.5E-08 | - | - | - | - | - | - | - | - |
| 7p15.3 | 20909622 | rs204592 | G | 1.2E-09 | 1.05 | 9.8E-11 | 1.02 | 6.7E-03 | 1.05 | 2.5E-13 | 1.02 | 1.9E-03 |
| 7p15.3 | 21839896 | rs6956349 | T | 8.6E-09 | 1.03 | 8.9E-05 | 1.01 | 0.55 | 1.03 | 5.6E-06 | 1.01 | 0.26 |
| 7p15.2 | 27281792 | rs17437810 | A | 8.0E-09 | 1.06 | 0.037 | 0.97 | 0.26 | 1.06 | 0.035 | 0.97 | 0.30 |
| 7p15.2 | 27483166 | rs10250340 | A | 2.0E-09 | 1.02 | 2.9E-03 | 1.00 | 0.78 | 1.02 | 1.2E-03 | 1.00 | 0.73 |
| 7p15.2 | 27975919 | rs67152137 | G | 8.6E-31 | 1.15 | 3.2E-53 | 1.08 | 8.9E-16 | 1.14 | 2.0E-63 | 1.08 | 5.5E-18 |
| 8p21.2 | 23437981 | rs2928681 | C | 3.5E-18 | 1.05 | 3.2E-10 | 1.01 | 0.18 | 1.04 | 8.5E-11 | 1.01 | 0.29 |
| 8p21.2 | 23529521 | rs1160267 | G | 6.3E-83 | 1.14 | 2.0E-68 | 1.05 | 1.8E-09 | 1.16 | 3.3E-112 | 1.07 | 2.1E-20 |
| 8q12.1 | 57138676 | rs36112366 | T | 7.3E-11 | 1.02 | 0.045 | 0.98 | 0.18 | 1.02 | 0.074 | 0.98 | 0.099 |
| 8q12.1 | 57834357 | rs4738555 | C | 3.2E-17 | 1.02 | 0.017 | 0.98 | 0.071 | 1.02 | 5.6E-03 | 0.99 | 0.072 |
| 8q21.13 | 81811566 | 8:81811566_CA_C | CA | 7.6E-11 | 0.99 | 0.072 | 0.96 | 2.4E-06 | 0.99 | 0.047 | 0.96 | 6.0E-07 |
| 8q23.1 | 108849527 | rs1494920 | T | 4.8E-08 | 1.02 | 0.041 | 0.99 | 0.44 | 1.01 | 0.063 | 0.99 | 0.23 |
| 8q24.21 | 127817460 | rs7843726 | G | 1.7E-08 | 1.05 | 7.8E-06 | 1.01 | 0.26 | 1.06 | 1.7E-08 | 1.02 | 0.057 |
| 8q24.21 | 128093297 | rs1016343 | T | 3.6E-15 | 1.25 | 2.4E-138 | 1.20 | 6.1E-72 | 1.25 | 5.9E-181 | 1.20 | 5.1E-88 |
| 8q24.21 | 128318755 | rs10107982 | T | 4.1E-30 | 1.15 | 4.6E-64 | 1.09 | 1.6E-20 | 1.15 | 1.3E-69 | 1.09 | 1.7E-22 |

| | | | | | | | | | | | | |
|---|---|---|---|---|---|---|---|---|---|---|---|---|
| 8q24.21 | 128413305 | rs6983267 | G | 2.6E-45 | 1.23 | 2.8E-164 | 1.16 | 2.6E-65 | 1.22 | 2.1E-198 | 1.16 | 1.7E-77 |
| 9q22.32 | 99146751 | rs7020681 | C | 1.4E-09 | 1.02 | 0.052 | 0.99 | 0.26 | 1.02 | 0.026 | 0.99 | 0.26 |
| 9q33.1 | 118258080 | rs150402584 | T | 9.0E-10 | 1.01 | 0.35 | 0.97 | 0.038 | 1.02 | 0.12 | 0.98 | 0.079 |
| 9q33.1 | 120475302 | rs4986790 | A | 7.1E-17 | 1.04 | 0.012 | 0.97 | 0.068 | 1.05 | 8.8E-04 | 0.98 | 0.19 |
| 9q33.2 | 123643426 | rs59482735 | T | 2.9E-12 | 1.03 | 3.3E-04 | 0.99 | 0.47 | 1.03 | 1.1E-04 | 0.99 | 0.53 |
| 10p12.31 | 22363979 | rs3011633 | C | 5.9E-20 | 1.01 | 0.21 | 0.97 | 9.5E-04 | 1.01 | 0.31 | 0.97 | 2.2E-04 |
| 10p12.2 | 23398448 | rs1857279 | C | 3.9E-11 | 1.02 | 0.010 | 0.99 | 0.36 | 1.03 | 9.9E-05 | 1.00 | 0.92 |
| 10p12.1 | 28096754 | rs2815506 | A | 4.9E-15 | 1.03 | 1.7E-03 | 0.99 | 0.32 | 1.03 | 5.8E-04 | 0.99 | 0.19 |
| 10q11.23 | 51549496 | rs10993994 | T | 7.3E-87 | 1.23 | 9.1E-165 | 1.11 | 2.8E-33 | 1.21 | 8.9E-192 | 1.10 | 3.1E-34 |
| 10q25.2 | 111991733 | rs562417881 | G | 5.6E-09 | 1.03 | 0.014 | 0.99 | 0.34 | 1.03 | 0.014 | 0.99 | 0.28 |
| 10q26.12 | 122631067 | rs9325569 | A | 9.9E-41 | 1.02 | 0.052 | 0.96 | 1.9E-06 | 1.01 | 0.19 | 0.95 | 7.9E-09 |
| 10q26.12 | 123049264 | rs10886902 | C | 8.2E-118 | 0.94 | 5.4E-11 | 0.84 | 4.9E-65 | 0.94 | 7.0E-13 | 0.85 | 4.3E-72 |
| 10q26.13 | 123135200 | rs10788173 | C | 2.9E-08 | 1.00 | 0.85 | 0.98 | 5.6E-03 | 1.00 | 0.72 | 0.98 | 1.6E-03 |
| 10q26.13 | 123185303 | rs10749415 | A | 1.8E-45 | 1.10 | 3.2E-08 | 0.97 | 0.14 | 1.11 | 5.2E-16 | 0.99 | 0.51 |
| 10q26.13 | 123349324 | rs45631563 | T | 7.9E-23 | 1.06 | 9.4E-04 | 0.96 | 0.044 | 1.07 | 1.9E-04 | 0.97 | 0.10 |
| 10q26.13 | 126737579 | rs3012065 | C | 2.2E-08 | 1.04 | 1.1E-07 | 1.02 | 0.047 | 1.04 | 1.1E-06 | 1.01 | 0.20 |
| 11p15.5 | 197557 | rs7103852 | G | 1.2E-09 | 1.00 | 0.98 | 0.95 | 4.7E-03 | 1.00 | 0.70 | 0.95 | 5.0E-04 |
| 11p13 | 34718279 | rs553481604 | G | 1.7E-11 | 1.00 | 0.56 | 0.96 | 6.9E-05 | 1.00 | 0.94 | 0.97 | 2.1E-04 |
| 11p13 | 34780936 | rs10466455 | C | 8.0E-26 | 1.01 | 0.23 | 0.97 | 9.7E-05 | 1.00 | 0.49 | 0.96 | 1.9E-06 |
| 11q13.5 | 76153279 | rs58015965 | G | 4.0E-08 | 1.06 | 8.3E-12 | 1.03 | 8.8E-04 | 1.05 | 3.6E-13 | 1.03 | 4.6E-04 |
| 11q22.2 | 102396607 | rs12285347 | C | 2.6E-25 | 0.93 | 4.0E-21 | 0.89 | 3.6E-40 | 0.93 | 5.4E-25 | 0.89 | 2.1E-45 |
| 11q22.2 | 102438778 | rs7129424 | A | 2.5E-11 | 0.98 | 1.8E-03 | 0.95 | 3.5E-09 | 0.97 | 2.8E-05 | 0.95 | 3.1E-12 |
| 12p13.31 | 9222286 | rs1805664 | T | 9.2E-09 | 1.01 | 0.42 | 0.98 | 0.042 | 1.01 | 0.42 | 0.98 | 0.033 |
| 12q14.3 | 66393756 | rs74097857 | T | 1.2E-09 | 1.05 | 2.1E-06 | 1.02 | 0.24 | 1.05 | 7.4E-06 | 1.01 | 0.35 |
| 12q21.31 | 84994000 | rs61928422 | T | 1.1E-08 | 1.02 | 0.031 | 0.99 | 0.40 | 1.02 | 0.016 | 0.99 | 0.37 |
| 12q24.21 | 115099805 | rs10774760 | G | 2.4E-30 | 1.03 | 8.1E-04 | 0.98 | 0.013 | 1.03 | 1.8E-04 | 0.98 | 0.012 |
| 12q24.21 | 115155460 | rs11067251 | G | 1.9E-09 | 1.01 | 0.39 | 0.98 | 0.046 | 1.01 | 0.48 | 0.98 | 0.024 |
| 12q24.23 | 118855432 | rs1045542 | G | 1.2E-13 | 1.00 | 0.89 | 0.95 | 2.2E-04 | 1.00 | 0.74 | 0.95 | 1.1E-04 |
| 13q14.3 | 51087443 | rs202346 | A | 1.8E-27 | 1.04 | 5.3E-06 | 0.99 | 0.21 | 1.04 | 2.9E-08 | 0.99 | 0.38 |
| 13q14.3 | 51165912 | rs12869529 | A | 9.4E-12 | 1.03 | 0.17 | 0.96 | 0.053 | 1.02 | 0.39 | 0.95 | 9.0E-03 |
| 13q14.3 | 51445360 | rs9563006 | A | 5.6E-11 | 1.03 | 2.3E-05 | 1.01 | 0.51 | 1.03 | 2.4E-06 | 1.01 | 0.38 |

| | | | | | | | | | | | | |
|---|---|---|---|---|---|---|---|---|---|---|---|---|
| 13q22.1 | 74055303 | rs1361057 | C | 3.3E-08 | 1.02 | 0.026 | 0.99 | 0.52 | 1.02 | 0.012 | 0.99 | 0.50 |
| 13q32.1 | 95799006 | rs61965887 | A | 3.7E-14 | 0.95 | 1.2E-03 | 0.89 | 1.4E-10 | 0.95 | 2.5E-03 | 0.89 | 6.3E-10 |
| 14q13.3 | 37240591 | rs712329 | G | 2.0E-08 | 1.01 | 0.11 | 0.99 | 0.21 | 1.01 | 0.21 | 0.99 | 0.078 |
| 14q32.13 | 94838142 | rs112635299 | G | 1.2E-15 | 1.14 | 1.7E-06 | 1.01 | 0.65 | 1.14 | 1.7E-06 | 1.02 | 0.56 |
| 14q32.13 | 95104745 | rs72695000 | C | 7.4E-28 | 1.04 | 2.8E-04 | 0.98 | 0.049 | 1.04 | 8.5E-05 | 0.98 | 0.088 |
| 15q22.31 | 66941399 | rs8038722 | G | 3.2E-08 | 1.01 | 0.049 | 0.99 | 0.35 | 1.03 | 4.4E-06 | 1.01 | 0.29 |
| 16p13.3 | 4318206 | rs251739 | G | 6.6E-20 | 1.03 | 1.2E-05 | 0.99 | 0.55 | 1.03 | 1.4E-06 | 1.00 | 0.55 |
| 16p13.13 | 11201428 | rs9933507 | C | 3.8E-10 | 1.00 | 0.54 | 0.97 | 5.1E-04 | 1.00 | 0.94 | 0.97 | 1.2E-03 |
| 16q23.1 | 77507275 | rs7206309 | C | 1.6E-22 | 1.00 | 0.86 | 0.96 | 8.3E-06 | 1.00 | 0.88 | 0.96 | 1.1E-06 |
| 16q24.1 | 86554784 | rs13334022 | T | 3.4E-11 | 1.00 | 0.86 | 0.95 | 2.0E-03 | 1.01 | 0.30 | 0.97 | 0.013 |
| 17p13.3 | 1994071 | rs3760230 | C | 2.8E-08 | 1.00 | 0.69 | 0.98 | 0.026 | 1.01 | 0.45 | 0.98 | 0.033 |
| 17p13.1 | 7571752 | rs78378222 | G | 2.8E-10 | 1.25 | 4.8E-11 | 1.10 | 0.013 | 1.25 | 4.7E-11 | 1.11 | 8.9E-03 |
| 17p13.1 | 7793367 | rs34009884 | C | 1.1E-08 | 1.10 | 1.2E-12 | 1.06 | 3.2E-04 | 1.10 | 1.5E-13 | 1.06 | 1.1E-04 |
| 17p12 | 12320741 | rs7219536 | T | 5.7E-12 | 1.01 | 0.24 | 0.98 | 0.035 | 1.01 | 0.090 | 0.98 | 0.072 |
| 17p12 | 12556260 | rs9889266 | C | 8.9E-10 | 1.02 | 0.039 | 0.99 | 0.27 | 1.01 | 0.21 | 0.98 | 0.040 |
| 17q12 | 36099952 | rs10908278 | A | 2.1E-46 | 1.23 | 2.7E-161 | 1.16 | 1.1E-63 | 1.22 | 1.1E-194 | 1.15 | 5.8E-75 |
| 18q23 | 76777252 | rs71279357 | A | 1.8E-12 | 1.09 | 8.0E-21 | 1.04 | 4.3E-05 | 1.08 | 1.2E-20 | 1.04 | 1.8E-04 |
| 19q12 | 32055024 | rs11084590 | C | 3.4E-18 | 1.00 | 0.69 | 0.96 | 4.7E-06 | 1.00 | 0.52 | 0.96 | 1.4E-06 |
| 19q13.32 | 46229711 | rs6509234 | G | 7.7E-09 | 1.01 | 0.36 | 0.98 | 0.038 | 1.01 | 0.53 | 0.97 | 0.014 |
| 19q13.33 | 49206462 | rs681343 | T | 4.4E-08 | 1.01 | 0.47 | 0.98 | 0.048 | 1.00 | 0.91 | 0.98 | 9.4E-03 |
| 19q13.33 | 51225544 | rs890863 | G | 1.1E-08 | 1.00 | 0.78 | 0.97 | 2.2E-03 | 0.99 | 0.26 | 0.97 | 7.7E-05 |
| 19q13.33 | 51323501 | rs1054713 | A | 4.8E-22 | 1.02 | 5.1E-03 | 0.98 | 0.026 | 1.02 | 9.2E-04 | 0.98 | 0.032 |
| 19q13.33 | 51340794 | rs2659053 | A | 5.4E-94 | 1.06 | 9.8E-15 | 0.97 | 1.2E-03 | 1.05 | 2.7E-12 | 0.96 | 2.7E-06 |
| 19q13.33 | 51349841 | rs113920094 | C | 6.5E-09 | 1.11 | 0.083 | 0.94 | 0.35 | 1.11 | 0.083 | 0.95 | 0.40 |
| 19q13.33 | 51360398 | rs578067733 | GA | 1.7E-12 | 1.06 | 0.61 | 0.88 | 0.30 | 1.10 | 0.025 | 0.92 | 0.081 |
| 19q13.33 | 51361757 | rs17632542 | T | 3.2e-638 | 1.35 | 8.3E-90 | 0.89 | 2.7E-12 | 1.35 | 3.9E-93 | 0.90 | 1.7E-10 |
| 19q13.33 | 51365466 | rs2735837 | G | 7.2E-58 | 1.06 | 1.3E-04 | 0.94 | 2.1E-04 | 1.08 | 3.2E-14 | 0.97 | 0.017 |
| 19q13.33 | 51373744 | rs12983994 | T | 1.1E-17 | 1.02 | 0.28 | 0.94 | 1.9E-03 | 1.01 | 0.40 | 0.94 | 1.4E-04 |
| 19q13.33 | 51378275 | rs112103380 | C | 1.7E-18 | 1.15 | 3.6E-03 | 0.90 | 0.059 | 1.15 | 3.6E-03 | 0.91 | 0.080 |
| 19q13.33 | 51383072 | rs198978 | G | 8.4E-31 | 1.00 | 0.55 | 0.95 | 9.1E-10 | 1.00 | 0.73 | 0.95 | 3.3E-10 |
| 19q13.41 | 51421334 | rs141135092 | C | 1.2E-09 | 1.05 | 0.097 | 0.95 | 0.14 | 1.06 | 0.050 | 0.96 | 0.25 |

| | | | | | | | | | | | | |
|---|---|---|---|---|---|---|---|---|---|---|---|---|
| 19q13.41 | 51441425 | rs80122351 | G | 1.2E-10 | 1.01 | 0.33 | 0.97 | 0.019 | 1.00 | 0.60 | 0.97 | 6.2E-03 |
| 20q11.21 | 31950845 | rs291671 | G | 1.2E-18 | 1.03 | 0.015 | 0.97 | 0.047 | 1.03 | 2.4E-03 | 0.97 | 0.038 |
| 21q22.2 | 40289167 | rs2836750 | T | 1.6E-12 | 1.04 | 3.5E-06 | 1.01 | 0.48 | 1.04 | 6.7E-07 | 1.01 | 0.47 |
| 22q13.31 | 45996298 | rs13268 | A | 1.6E-10 | 1.02 | 0.47 | 0.93 | 8.8E-03 | 1.02 | 0.48 | 0.93 | 0.010 |
| Xp22.31 | 8912815 | rs112062538 | C | 4.7E-19 | 1.02 | 2.5E-03 | 0.99 | 0.035 | 1.02 | 4.7E-03 | 0.98 | 8.8E-03 |
| Xp22.2 | 16844831 | rs62586587 | G | 3.6E-25 | 1.02 | 1.2E-03 | 0.98 | 8.0E-03 | 1.02 | 4.3E-04 | 0.98 | 6.6E-03 |
| Xp22.13 | 17831188 | rs200615037 | TTG | 3.5E-22 | 1.06 | 1.2E-04 | 0.98 | 0.26 | 1.06 | 1.9E-05 | 0.98 | 0.19 |
| Xp22.11 | 24109619 | rs7065158 | A | 1.1E-29 | 1.01 | 0.10 | 0.97 | 1.0E-05 | 1.01 | 0.15 | 0.97 | 3.4E-06 |
| Xp11.22 | 51242364 | rs5987424 | A | 2.1E-08 | 1.11 | 1.1E-78 | 1.09 | 2.4E-37 | 1.10 | 4.6E-84 | 1.08 | 4.1E-39 |
| Xp11.21 | 55936822 | rs10855058 | A | 7.6E-09 | 1.02 | 5.7E-04 | 1.00 | 1.00 | 1.02 | 8.3E-05 | 1.00 | 0.89 |
| Xq13.1 | 68996186 | rs2520386 | G | 4.2E-11 | 1.02 | 0.025 | 0.99 | 0.16 | 1.01 | 0.060 | 0.99 | 0.089 |
| Xq13.1 | 70125103 | rs62608084 | A | 1.7E-24 | 1.04 | 1.4E-14 | 1.01 | 0.37 | 1.04 | 4.6E-14 | 1.00 | 0.49 |
| Xq26.2 | 132935739 | rs4829762 | T | 5.9E-12 | 1.02 | 3.8E-03 | 0.99 | 0.23 | 1.01 | 0.012 | 0.99 | 0.056 |
| Xq26.3 | 133703924 | rs5930651 | T | 1.0E-08 | 1.01 | 0.13 | 0.99 | 0.14 | 1.01 | 0.14 | 0.99 | 0.077 |

[1] Bias-corrected estimated were obtained as follows: $\beta^{adj} = \beta - b * \beta_{PSA}$ and the corresponding $OR^{adj} = \exp(\beta^{adj})$

[2] Bias-corrected multi-ancestry effects should be interpreted with caution since the bias correction factor ($b$) was estimated from variants that were LD-pruned ($r^2$<0.10) based on 1000G EUR. The same set of pruned variants was used for European and multi-ancestry bias analyses. For the multi-ancestry analysis SNP effects on PSA and prostate cancer were obtained from multi-ancestry GWAS summary statistics.

Abbreviations:

EAF Effect allele frequency

**Supplementary Table 9: Comparison of methods for quantifying index event bias.** Estimates of the bias correction factor ($b$) derived using the Dudbridge method[33] and SlopeHunter[35] are presented. In each analysis PSA variation corresponds to the selection trait and prostate cancer is the outcome trait.

| **Method** | **GWAS** | $P_{thresh}$[1] | $b$ | $SE_b$ | **(95% CI)**[2] |
|---|---|---|---|---|---|
| Dudbridge | EUR | - | 1.1436 | $2.91\times10^{-4}$ | (1.1431 – 1.1442) |
| | Multi-ancestry[3] | - | 1.1039 | $1.49\times10^{-4}$ | (1.1037 – 1.1043) |
| SlopeHunter | EUR | 0.1 | 0.382 | 0.096 | (0.195 – 0.570) |
| | | 0.05 | 0.368 | 0.036 | (0.298 – 0.439) |
| | | 0.01 | 0.322 | 0.157 | (0.014 – 0.630) |
| | | 0.001 | 0.476 | 0.135 | (0.213 – 0.740) |
| | | 0.005 | 0.333 | 0.134 | (0.070 – 0.595) |
| | | $5.0\times10^{-4}$ | 0.452 | 0.106 | (0.245 – 0.659) |

[1] P-value threshold for SNP-incidence associations. Associations with p-values larger than $P_{thresh}$ are excluded prior to fitting the main model-based clustering in SlopeHunter. The default recommended setting is 0.001. This parameter is only applicable to SlopeHunter, no thresholding or clustering is performed in the Dudbridge method.

[2] Bootstrapped 95% confidence intervals estimated based on 1500 replicates are expected to have better coverage than empirical 95% confidence intervals based on $SE_b$ when the normal approximation for outcome effect sizes may be inappropriate.

[3] Multi-ancestry results should be interpreted with caution because they were estimated using the same set of variants as for the European ancestry analysis, selected by LD-pruning ($r^2$<0.10) 1000G EUR. Effect estimates for the reference panel variants were obtained from multi-ancestry GWAS meta-analyses for PSA and prostate cancer and used as inputs to estimate the bias correction factor ($b$)

**Supplementary Table 10: Impact of index event bias correction on 209 prostate cancer variants.** Independent risk variants for prostate cancer were selected from the PRACTICAL GWAS meta-analysis by Conti et al.[32] using linkage disequilibrium (LD) clumping (LD $r^2$<0.01, $P$<5×$10^{-8}$). GWAS odds ratios (OR) and two-sided p-values for prostate cancer are based on an inverse-variance-weighted fixed-effects meta-analysis of European ancestry studies (85,554 cases and 91,972 controls). Original GWAS results are compared to for associations adjusted for index event bias using the Dudbridge et al. method[33] and SlopeHunter[35]. For each variant, bias-adjusted odds ratios ($OR_{adj}$) and two-sided p-values ($P_{adj}$) were calculated from a chi-squared test statistic derived from bias-corrected effect sizes and standard errors. For additional context, associations with PSA levels are provided. For each variant, the PSA effect size ($β_{PSA}$) and two-sided p-value ($P_{PSA}$) are based on an inverse-variance-weighted fixed-effects meta-analysis in men of European ancestry (n=85,824).

| Region | Position (b37) | Index variant ID[1] | Prostate Cancer Alleles | | PSA Estimates | | Prostate Cancer Estimates: Original GWAS | | Prostate Cancer Estimates: Bias Corrected[3] | | Prostate Cancer Estimates: Bias Corrected[4] | |
|---|---|---|---|---|---|---|---|---|---|---|---|---|
| | | | Risk | Other | $β_{PSA}$[2] | $P_{PSA}$ | OR | P | $OR_{adj}$ | $P_{adj}$ | $OR_{adj\text{-}S}$ | $P_{adj\text{-}S}$ |
| 1p36.22 | 10561604 | rs6658216 | T | C | 0.005 | 0.20 | 1.05 | 4.47E-08 | 1.04 | 4.40E-05 | 1.04 | 6.21E-07 |
| 1p36.13 | 16380393 | rs35708002 | A | G | 0.016 | 2.99E-03 | 1.06 | 3.06E-08 | 1.04 | 9.56E-04 | 1.05 | 4.81E-06 |
| 1p22.3 | 88224230 | rs305423 | T | C | 0.032 | 1.55E-10 | 1.06 | 2.80E-09 | 1.02 | 0.062 | 1.04 | 1.02E-04 |
| 1q21.3 | 150609429 | rs116173394 | A | G | -0.002 | 0.83 | 1.10 | 1.72E-08 | 1.10 | 6.79E-07 | 1.10 | 3.55E-08 |
| 1q21.3 | 150868102 | rs78132593 | A | C | 0.007 | 0.13 | 1.08 | 3.26E-16 | 1.07 | 3.58E-10 | 1.07 | 5.40E-14 |
| 1q21.3 | 150930926 | rs114908993 | C | T | -0.018 | 0.11 | 1.16 | 3.49E-12 | 1.18 | 1.57E-11 | 1.17 | 1.58E-12 |
| 1q21.3 | 153923276 | rs10127983 | T | C | 0.013 | 2.38E-03 | 1.07 | 1.38E-16 | 1.05 | 2.31E-08 | 1.06 | 9.31E-13 |
| 1q21.3 | 154980351 | rs56103503 | T | C | 0.026 | 6.27E-08 | 1.07 | 3.16E-17 | 1.04 | 2.27E-04 | 1.05 | 2.27E-09 |
| 1q22 | 155552910 | rs61812109 | G | A | 0.037 | 3.04E-03 | 1.23 | 9.04E-12 | 1.18 | 9.09E-07 | 1.21 | 1.56E-09 |
| 1q22 | 155668846 | rs190811727 | G | A | -0.003 | 0.85 | 1.17 | 1.54E-08 | 1.18 | 3.25E-07 | 1.17 | 2.79E-08 |
| 1q32.1 | 204029748 | rs4593892 | T | C | 0.000 | 0.99 | 1.05 | 5.38E-10 | 1.05 | 8.19E-08 | 1.05 | 1.74E-09 |
| 1q32.1 | 204518842 | rs4245739 | A | C | 0.014 | 1.39E-03 | 1.09 | 2.61E-26 | 1.08 | 1.97E-14 | 1.09 | 5.04E-21 |
| 1q32.1 | 205738302 | rs823137 | G | A | 0.012 | 1.20E-03 | 1.05 | 3.84E-09 | 1.03 | 4.87E-04 | 1.04 | 1.17E-06 |
| 2p25.1 | 8597123 | rs62106670 | T | C | -0.005 | 0.31 | 1.05 | 5.98E-09 | 1.06 | 8.34E-08 | 1.05 | 4.72E-09 |
| 2p25.1 | 10094526 | rs73913932 | G | A | -0.004 | 0.58 | 1.09 | 2.14E-09 | 1.09 | 4.52E-08 | 1.09 | 2.75E-09 |
| 2p25.1 | 10781975 | rs1990613 | T | C | 0.003 | 0.49 | 1.07 | 1.87E-19 | 1.07 | 7.87E-14 | 1.07 | 6.89E-18 |
| 2p24.3 | 16024757 | rs12611818 | T | G | 0.001 | 0.78 | 1.05 | 1.01E-08 | 1.05 | 8.64E-07 | 1.05 | 3.40E-08 |
| 2p24.1 | 20878105 | rs9306894 | G | A | 0.030 | 1.53E-14 | 1.08 | 1.21E-21 | 1.04 | 1.25E-05 | 1.06 | 3.03E-11 |
| 2p21 | 43148487 | rs4952948 | G | A | 0.010 | 0.015 | 1.04 | 2.86E-08 | 1.03 | 3.32E-04 | 1.04 | 1.94E-06 |
| 2p21 | 43637998 | rs7591218 | A | G | 0.015 | 3.45E-04 | 1.08 | 2.54E-24 | 1.07 | 3.54E-12 | 1.08 | 2.06E-18 |
| 2p15 | 62792689 | rs13008096 | A | C | 0.001 | 0.84 | 1.05 | 2.51E-11 | 1.05 | 1.51E-08 | 1.05 | 1.28E-10 |

| | | | | | | | | | | | | |
|---|---|---|---|---|---|---|---|---|---|---|---|---|
| 2p15 | 63301164 | rs6545977 | G | A | 0.026 | 4.25E-12 | 1.12 | 7.06E-50 | 1.08 | 8.93E-21 | 1.10 | 3.37E-31 |
| 2p15 | 63982690 | rs72808211 | G | A | 0.016 | 0.11 | 1.15 | 2.85E-11 | 1.12 | 3.80E-07 | 1.14 | 1.18E-09 |
| 2p14 | 66652885 | rs74702681 | T | C | 0.010 | 0.45 | 1.17 | 9.89E-11 | 1.16 | 2.29E-07 | 1.17 | 1.24E-09 |
| 2p11.2 | 85764960 | rs1446668 | G | T | -0.014 | 3.95E-04 | 1.09 | 2.51E-28 | 1.10 | 6.18E-30 | 1.09 | 1.65E-29 |
| 2q13 | 111893096 | rs11691517 | T | G | 0.007 | 0.10 | 1.07 | 1.33E-13 | 1.06 | 2.73E-08 | 1.06 | 1.61E-11 |
| 2q13 | 111906762 | rs1877330 | A | C | 0.004 | 0.60 | 1.11 | 6.85E-12 | 1.10 | 1.32E-08 | 1.11 | 6.30E-11 |
| 2q31.1 | 173287607 | rs11686909 | G | A | 0.001 | 0.80 | 1.05 | 4.66E-10 | 1.05 | 1.48E-07 | 1.05 | 1.90E-09 |
| 2q31.1 | 173303031 | rs28485589 | A | G | 0.019 | 0.020 | 1.27 | 5.01E-48 | 1.25 | 2.56E-31 | 1.26 | 1.38E-41 |
| 2q33.1 | 202126615 | rs1861270 | G | A | 0.005 | 0.28 | 1.05 | 2.81E-09 | 1.05 | 3.53E-06 | 1.05 | 3.14E-08 |
| 2q33.3 | 208118301 | rs12621900 | C | T | 0.006 | 0.17 | 1.05 | 3.45E-08 | 1.05 | 2.69E-05 | 1.05 | 4.02E-07 |
| 2q37.3 | 238440449 | rs11891348 | G | T | 0.014 | 2.04E-03 | 1.07 | 2.39E-13 | 1.05 | 2.39E-06 | 1.06 | 5.85E-10 |
| 2q37.3 | 242135265 | rs77559646 | A | G | 0.047 | 4.96E-04 | 1.27 | 1.59E-21 | 1.21 | 2.64E-10 | 1.25 | 4.32E-16 |
| 2q37.3 | 242139600 | rs77482050 | G | A | 0.057 | 0.012 | 1.48 | 1.45E-22 | 1.39 | 6.61E-12 | 1.44 | 6.42E-18 |
| 2q37.3 | 242157241 | rs76832527 | A | G | 0.007 | 0.16 | 1.10 | 5.00E-22 | 1.09 | 3.18E-14 | 1.10 | 2.27E-19 |
| 3p24.3 | 23153062 | rs7618603 | A | C | 0.019 | 1.15E-04 | 1.06 | 3.08E-08 | 1.03 | 3.69E-03 | 1.05 | 1.33E-05 |
| 3p21.31 | 49626306 | rs6446285 | G | A | 0.005 | 0.30 | 1.06 | 1.12E-08 | 1.05 | 1.06E-05 | 1.06 | 1.25E-07 |
| 3p14.1 | 64920084 | rs55821157 | G | A | 0.010 | 0.24 | 1.11 | 6.87E-09 | 1.10 | 7.07E-06 | 1.11 | 8.75E-08 |
| 3p13 | 70874997 | rs7622519 | G | A | 0.017 | 7.72E-06 | 1.04 | 2.46E-08 | 1.02 | 7.87E-03 | 1.04 | 2.49E-05 |
| 3p12.1 | 87184311 | rs6770433 | T | C | 0.005 | 0.16 | 1.12 | 1.32E-50 | 1.11 | 1.94E-34 | 1.11 | 1.56E-45 |
| 3p11.2 | 87395326 | rs114423020 | G | A | 0.014 | 0.19 | 1.16 | 4.41E-13 | 1.14 | 1.88E-08 | 1.15 | 2.05E-11 |
| 3q13.12 | 106962305 | rs1283105 | G | C | 0.000 | 0.94 | 1.05 | 3.71E-11 | 1.05 | 1.34E-08 | 1.05 | 1.62E-10 |
| 3q13.2 | 113290450 | rs13092681 | T | C | 0.023 | 1.15E-09 | 1.09 | 2.24E-29 | 1.06 | 2.12E-11 | 1.08 | 1.14E-18 |
| 3q21.3 | 127898501 | rs2811476 | C | A | 0.024 | 1.79E-08 | 1.10 | 4.59E-32 | 1.07 | 4.23E-13 | 1.09 | 6.04E-21 |
| 3q21.3 | 128275459 | rs4857906 | C | T | 0.010 | 0.036 | 1.06 | 2.74E-11 | 1.05 | 2.59E-06 | 1.06 | 3.40E-09 |
| 3q23 | 141140968 | rs9846396 | T | C | 0.030 | 4.68E-15 | 1.05 | 2.91E-10 | 1.01 | 0.13 | 1.03 | 1.43E-04 |
| 3q25.1 | 152004202 | rs182314334 | T | C | -0.013 | 0.075 | 1.09 | 1.08E-11 | 1.11 | 3.05E-11 | 1.10 | 3.42E-12 |
| 3q26.2 | 170074517 | rs78416326 | G | C | 0.017 | 4.31E-04 | 1.18 | 4.06E-68 | 1.16 | 3.40E-41 | 1.17 | 5.67E-56 |
| 4q13.3 | 74326647 | rs113573136 | T | C | 0.005 | 0.49 | 1.06 | 2.50E-10 | 1.05 | 2.78E-05 | 1.05 | 1.77E-08 |
| 4q13.3 | 74442349 | rs17804499 | G | C | 0.016 | 0.047 | 1.17 | 9.68E-19 | 1.15 | 4.02E-12 | 1.16 | 3.72E-16 |
| 4q22.3 | 95547467 | rs7679542 | A | G | 0.006 | 0.11 | 1.09 | 5.03E-27 | 1.08 | 1.84E-17 | 1.08 | 1.22E-23 |
| 4q24 | 106065308 | rs10007915 | C | G | 0.011 | 5.96E-03 | 1.13 | 5.22E-55 | 1.11 | 7.46E-34 | 1.12 | 2.28E-46 |

| | | | | | | | | | | | | |
|---|---|---|---|---|---|---|---|---|---|---|---|---|
| 4q31.1 | 140915940 | rs13148756 | T | G | -0.006 | 0.35 | 1.05 | 4.50E-08 | 1.06 | 2.23E-06 | 1.05 | 6.62E-08 |
| 5p15.33 | 1228166 | rs114674839 | T | C | 0.043 | 1.10E-05 | 1.12 | 3.78E-09 | 1.07 | 3.78E-03 | 1.10 | 6.59E-06 |
| 5p15.33 | 1272074 | rs138895564 | T | C | 0.101 | 7.29E-06 | 1.27 | 1.75E-08 | 1.13 | 0.013 | 1.21 | 3.34E-05 |
| 5p15.33 | 1280028 | rs2242652 | G | A | 0.011 | 0.031 | 1.16 | 5.75E-51 | 1.15 | 6.29E-33 | 1.16 | 2.78E-44 |
| 5p15.33 | 1292118 | rs71595003 | A | G | -0.023 | 0.059 | 1.19 | 1.73E-15 | 1.22 | 1.00E-14 | 1.20 | 6.15E-16 |
| 5p15.33 | 1895829 | rs12653946 | T | C | 0.010 | 9.17E-03 | 1.08 | 2.57E-22 | 1.07 | 1.24E-12 | 1.07 | 3.04E-18 |
| 5q31.1 | 133836209 | rs10793821 | T | C | 0.005 | 0.17 | 1.06 | 2.26E-14 | 1.05 | 2.54E-09 | 1.06 | 1.25E-12 |
| 5q35.2 | 172959030 | rs9686557 | C | A | 0.016 | 2.42E-05 | 1.04 | 4.69E-09 | 1.03 | 3.03E-03 | 1.04 | 5.84E-06 |
| 5q35.3 | 177891551 | rs2672843 | G | A | 0.006 | 0.15 | 1.05 | 4.19E-09 | 1.04 | 1.08E-05 | 1.04 | 8.87E-08 |
| 6p24.2 | 11217897 | rs2018336 | T | C | 0.012 | 6.10E-03 | 1.07 | 1.51E-12 | 1.05 | 1.61E-06 | 1.06 | 7.66E-10 |
| 6p22.3 | 22314766 | rs112292679 | C | T | -0.001 | 0.93 | 1.11 | 2.02E-08 | 1.11 | 8.66E-07 | 1.11 | 4.41E-08 |
| 6p21.33 | 30611676 | rs1140809 | C | A | 0.011 | 4.85E-03 | 1.05 | 1.01E-09 | 1.03 | 1.27E-04 | 1.04 | 2.41E-07 |
| 6p21.33 | 31329386 | rs2596546 | A | G | 0.019 | 2.58E-06 | 1.07 | 4.71E-16 | 1.04 | 4.07E-06 | 1.06 | 1.31E-10 |
| 6p21.32 | 32604795 | rs9272390 | T | C | 0.007 | 0.22 | 1.06 | 1.62E-11 | 1.06 | 1.27E-06 | 1.06 | 1.04E-09 |
| 6p21.31 | 34793124 | rs9469899 | A | G | 0.022 | 1.34E-08 | 1.05 | 1.64E-09 | 1.02 | 0.017 | 1.04 | 2.31E-05 |
| 6p21.1 | 41538545 | rs913074 | C | T | 0.010 | 0.046 | 1.09 | 3.68E-24 | 1.07 | 2.22E-12 | 1.08 | 3.17E-19 |
| 6q14.1 | 76479489 | rs6907476 | T | C | 0.009 | 0.093 | 1.06 | 6.51E-09 | 1.05 | 3.06E-05 | 1.06 | 2.18E-07 |
| 6q21 | 109295293 | rs2038542 | C | T | 0.005 | 0.41 | 1.08 | 5.49E-13 | 1.07 | 7.33E-09 | 1.08 | 1.06E-11 |
| 6q22.1 | 117200434 | rs339351 | C | A | 0.011 | 7.84E-03 | 1.09 | 2.48E-27 | 1.08 | 7.00E-16 | 1.09 | 2.04E-22 |
| 6q23.2 | 134300641 | rs6919121 | T | C | 0.007 | 0.061 | 1.05 | 4.04E-09 | 1.04 | 1.74E-05 | 1.05 | 1.33E-07 |
| 6q25.2 | 153430868 | rs63739861 | A | C | 0.002 | 0.60 | 1.08 | 5.55E-22 | 1.07 | 4.99E-16 | 1.07 | 2.04E-20 |
| 6q25.3 | 160184600 | rs144384356 | G | A | 0.018 | 0.071 | 1.13 | 2.31E-10 | 1.11 | 5.70E-06 | 1.12 | 1.43E-08 |
| 6q25.3 | 160255934 | rs73030808 | A | G | 0.014 | 0.011 | 1.08 | 1.52E-12 | 1.06 | 1.42E-06 | 1.07 | 6.66E-10 |
| 6q25.3 | 160560845 | rs628031 | A | G | 0.004 | 0.32 | 1.07 | 2.83E-19 | 1.07 | 3.20E-13 | 1.07 | 2.31E-17 |
| 6q25.3 | 160584693 | rs7749378 | A | G | 0.013 | 0.26 | 1.14 | 4.28E-09 | 1.12 | 6.86E-06 | 1.13 | 5.98E-08 |
| 6q25.3 | 160584841 | rs116910573 | G | A | 0.000 | 0.97 | 1.13 | 8.54E-10 | 1.13 | 9.59E-08 | 1.13 | 2.51E-09 |
| 6q25.3 | 160628128 | rs117648937 | C | T | 0.000 | 0.99 | 1.29 | 1.47E-11 | 1.29 | 3.68E-09 | 1.29 | 5.41E-11 |
| 6q25.3 | 160822675 | rs3105751 | G | A | 0.018 | 2.33E-05 | 1.11 | 6.02E-39 | 1.09 | 1.76E-19 | 1.10 | 6.49E-29 |
| 7p21.1 | 20444081 | rs6973059 | T | C | -0.001 | 0.76 | 1.05 | 8.67E-09 | 1.05 | 2.85E-07 | 1.05 | 1.74E-08 |
| 7p15.3 | 20999211 | rs9655205 | C | A | 0.028 | 4.43E-07 | 1.09 | 2.22E-24 | 1.06 | 7.46E-08 | 1.08 | 1.15E-14 |
| 7p15.3 | 21800369 | rs10281087 | T | G | 0.014 | 3.53E-04 | 1.04 | 2.38E-08 | 1.03 | 2.39E-03 | 1.04 | 7.89E-06 |

| | | | | | | | | | | | | |
|---|---|---|---|---|---|---|---|---|---|---|---|---|
| 7p15.2 | 27220831 | rs6461992 | G | A | 0.006 | 0.43 | 1.09 | 7.43E-10 | 1.09 | 9.08E-07 | 1.09 | 7.79E-09 |
| 7p15.2 | 27976563 | rs10486567 | G | A | 0.049 | 2.42E-28 | 1.15 | 2.48E-53 | 1.08 | 4.91E-15 | 1.12 | 1.76E-23 |
| 7p14.1 | 40875192 | rs17621345 | A | C | 0.021 | 1.91E-06 | 1.07 | 1.96E-14 | 1.05 | 1.19E-05 | 1.06 | 1.07E-09 |
| 7p12.3 | 47482829 | rs4724578 | C | A | 0.002 | 0.61 | 1.06 | 2.40E-13 | 1.05 | 1.11E-09 | 1.06 | 2.59E-12 |
| 7p12.3 | 47506994 | rs2964944 | C | T | 0.015 | 3.93E-04 | 1.05 | 2.61E-08 | 1.03 | 1.44E-03 | 1.04 | 6.09E-06 |
| 7p12.3 | 47627603 | rs112289173 | C | T | 0.002 | 0.67 | 1.07 | 4.13E-08 | 1.07 | 3.19E-06 | 1.07 | 1.48E-07 |
| 7q21.3 | 97688440 | rs4727386 | A | G | 0.015 | 1.11E-03 | 1.10 | 6.50E-40 | 1.09 | 1.10E-19 | 1.10 | 2.33E-30 |
| 8p21.3 | 22879734 | rs4871844 | T | C | 0.020 | 5.87E-05 | 1.04 | 4.75E-08 | 1.02 | 0.029 | 1.04 | 9.38E-05 |
| 8p21.2 | 23443320 | rs11996109 | C | A | 0.045 | 1.93E-13 | 1.06 | 1.14E-13 | 1.00 | 0.63 | 1.04 | 5.86E-04 |
| 8p21.2 | 23533623 | rs11135766 | C | T | 0.069 | 2.27E-72 | 1.14 | 4.17E-69 | 1.05 | 1.39E-09 | 1.10 | 3.00E-16 |
| 8p21.2 | 25892142 | rs11135910 | T | C | 0.006 | 0.24 | 1.08 | 2.23E-13 | 1.07 | 6.73E-09 | 1.08 | 8.25E-12 |
| 8q24.21 | 127855936 | rs4870985 | T | C | -0.009 | 0.036 | 1.08 | 1.88E-24 | 1.09 | 6.26E-22 | 1.08 | 1.12E-24 |
| 8q24.21 | 127906604 | rs13282012 | G | A | -0.004 | 0.27 | 1.10 | 2.92E-39 | 1.11 | 9.19E-33 | 1.10 | 1.29E-38 |
| 8q24.21 | 128018466 | rs73351629 | C | G | 0.009 | 0.020 | 1.15 | 9.42E-69 | 1.14 | 6.91E-45 | 1.15 | 1.17E-59 |
| 8q24.21 | 128077146 | rs77541621 | A | G | 0.048 | 2.05E-04 | 1.83 | 1.13E-174 | 1.73 | 1.16E-98 | 1.79 | 3.93E-137 |
| 8q24.21 | 128103979 | rs5013678 | T | C | 0.009 | 0.047 | 1.20 | 1.87E-79 | 1.18 | 1.07E-54 | 1.19 | 3.71E-71 |
| 8q24.21 | 128108418 | rs111724742 | G | T | 0.039 | 0.066 | 1.61 | 3.15E-43 | 1.54 | 6.87E-25 | 1.58 | 2.61E-36 |
| 8q24.21 | 128117736 | rs143368544 | T | C | 0.050 | 2.51E-04 | 1.25 | 2.48E-17 | 1.18 | 5.35E-08 | 1.22 | 1.09E-12 |
| 8q24.21 | 128151672 | rs17446776 | G | C | 0.007 | 0.58 | 1.25 | 9.43E-22 | 1.24 | 2.48E-15 | 1.25 | 6.16E-20 |
| 8q24.21 | 128228233 | rs79125478 | T | C | 0.006 | 0.52 | 1.11 | 1.18E-11 | 1.10 | 2.66E-07 | 1.10 | 2.95E-10 |
| 8q24.21 | 128294091 | rs283713 | G | T | 0.009 | 0.16 | 1.09 | 1.64E-12 | 1.08 | 8.84E-08 | 1.09 | 9.92E-11 |
| 8q24.21 | 128324147 | rs382434 | C | T | 0.036 | 1.98E-14 | 1.15 | 1.80E-72 | 1.11 | 6.24E-26 | 1.13 | 5.03E-39 |
| 8q24.21 | 128376862 | rs117627785 | T | C | 0.040 | 5.35E-03 | 1.23 | 1.36E-13 | 1.18 | 6.42E-07 | 1.21 | 1.50E-10 |
| 8q24.21 | 128413305 | rs6983267 | G | T | 0.052 | 1.49E-42 | 1.23 | 2.84E-164 | 1.15 | 3.71E-63 | 1.20 | 8.38E-67 |
| 8q24.21 | 128514011 | rs74592433 | T | C | 0.020 | 0.13 | 1.23 | 8.17E-10 | 1.20 | 6.44E-07 | 1.22 | 1.01E-08 |
| 8q24.21 | 128532137 | rs10090154 | T | C | 0.012 | 0.055 | 1.44 | 4.15E-225 | 1.42 | 4.10E-146 | 1.43 | 2.00E-197 |
| 8q24.21 | 128582965 | rs73709253 | G | A | 0.013 | 0.30 | 1.14 | 8.74E-09 | 1.13 | 8.58E-06 | 1.14 | 9.96E-08 |
| 9p22.1 | 18556233 | rs951227 | A | G | 0.001 | 0.80 | 1.05 | 6.68E-10 | 1.05 | 1.84E-07 | 1.05 | 3.44E-09 |
| 9p22.1 | 19055965 | rs1048169 | C | T | 0.007 | 0.085 | 1.06 | 2.81E-14 | 1.05 | 1.19E-08 | 1.06 | 4.18E-12 |
| 9p21.3 | 22041998 | rs17694493 | G | C | -0.001 | 0.88 | 1.08 | 3.37E-13 | 1.08 | 2.27E-10 | 1.08 | 1.44E-12 |
| 9p13.3 | 33935073 | rs307651 | A | C | 0.010 | 0.010 | 1.05 | 3.24E-11 | 1.04 | 9.30E-06 | 1.05 | 9.01E-09 |

| | | | | | | | | | | | | |
|---|---|---|---|---|---|---|---|---|---|---|---|---|
| 9q21.31 | 82087596 | rs9314679 | T | A | -0.011 | 0.052 | 1.07 | 1.02E-08 | 1.08 | 2.88E-09 | 1.08 | 2.48E-09 |
| 9q31.2 | 110290217 | rs143655302 | G | A | 0.064 | 5.48E-06 | 1.27 | 2.29E-14 | 1.18 | 2.63E-06 | 1.23 | 3.61E-10 |
| 9q34.11 | 132576060 | rs1182 | A | C | 0.006 | 0.23 | 1.06 | 5.65E-11 | 1.05 | 4.42E-07 | 1.06 | 1.34E-09 |
| 10p15.3 | 890714 | rs34318754 | A | G | 0.014 | 5.99E-03 | 1.08 | 9.23E-13 | 1.06 | 1.17E-06 | 1.07 | 5.00E-10 |
| 10q11.21 | 45854778 | rs11511618 | G | A | 0.023 | 2.58E-06 | 1.06 | 1.66E-10 | 1.04 | 1.22E-03 | 1.05 | 1.01E-06 |
| 10q11.22 | 47671874 | rs34583261 | C | T | 0.019 | 0.11 | 1.09 | 3.52E-08 | 1.06 | 2.55E-03 | 1.08 | 6.56E-06 |
| 10q11.23 | 51549496 | rs10993994 | T | C | 0.085 | 2.66E-76 | 1.23 | 9.11E-165 | 1.11 | 2.35E-32 | 1.18 | 1.01E-32 |
| 10q23.31 | 90195149 | rs1935581 | C | T | -0.006 | 0.12 | 1.05 | 6.36E-09 | 1.05 | 6.85E-09 | 1.05 | 2.52E-09 |
| 10q24.32 | 104408272 | rs12241712 | C | A | 0.015 | 2.86E-04 | 1.07 | 5.24E-18 | 1.06 | 1.35E-08 | 1.07 | 2.32E-13 |
| 10q25.2 | 114715598 | rs2104598 | A | G | 0.007 | 0.069 | 1.05 | 3.70E-09 | 1.04 | 2.68E-05 | 1.04 | 1.35E-07 |
| 10q26.12 | 122787452 | rs12245671 | G | A | 0.001 | 0.81 | 1.06 | 7.05E-14 | 1.06 | 2.13E-10 | 1.06 | 5.45E-13 |
| 10q26.12 | 123034391 | rs4468286 | C | A | -0.086 | 1.82E-89 | 1.06 | 2.08E-10 | 1.17 | 1.02E-53 | 1.10 | 6.06E-11 |
| 10q26.13 | 123190200 | rs17101982 | C | G | 0.101 | 1.19E-31 | 1.11 | 5.40E-09 | 0.99 | 0.48 | 1.05 | 0.018 |
| 10q26.13 | 126697114 | rs4962419 | A | G | 0.021 | 6.08E-07 | 1.06 | 3.19E-11 | 1.03 | 1.23E-03 | 1.05 | 5.79E-07 |
| 11p15.5 | 1507512 | rs1881502 | T | C | 0.003 | 0.54 | 1.06 | 3.04E-09 | 1.05 | 1.37E-06 | 1.06 | 2.10E-08 |
| 11p15.5 | 2233797 | rs10840603 | A | G | 0.018 | 2.61E-04 | 1.18 | 1.04E-68 | 1.15 | 1.06E-38 | 1.17 | 3.73E-54 |
| 11p15.4 | 7547587 | rs61890184 | A | G | -0.004 | 0.45 | 1.08 | 2.73E-10 | 1.08 | 6.35E-09 | 1.08 | 3.59E-10 |
| 11q12.3 | 61908440 | rs2277283 | C | T | 0.006 | 0.12 | 1.05 | 1.61E-10 | 1.05 | 1.34E-06 | 1.05 | 4.55E-09 |
| 11q13.2 | 66951965 | rs12785905 | C | G | 0.015 | 0.075 | 1.12 | 5.10E-10 | 1.10 | 3.32E-06 | 1.11 | 1.68E-08 |
| 11q13.3 | 68912221 | rs3018667 | G | A | -0.001 | 0.90 | 1.06 | 5.04E-15 | 1.06 | 7.74E-12 | 1.06 | 2.36E-14 |
| 11q13.3 | 68981359 | rs12275055 | G | A | 0.015 | 3.49E-03 | 1.25 | 2.76E-111 | 1.23 | 2.00E-70 | 1.24 | 1.06E-93 |
| 11q13.3 | 69071624 | rs11228619 | G | A | 0.012 | 0.14 | 1.10 | 5.33E-10 | 1.09 | 5.79E-06 | 1.09 | 1.97E-08 |
| 11q13.3 | 69456000 | rs55911137 | C | G | 0.030 | 0.014 | 1.20 | 4.32E-14 | 1.16 | 1.25E-07 | 1.18 | 3.00E-11 |
| 11q13.5 | 76251818 | rs17749618 | A | G | 0.021 | 2.32E-07 | 1.06 | 8.08E-13 | 1.03 | 3.02E-04 | 1.05 | 5.80E-08 |
| 11q22.2 | 102396607 | rs12285347 | T | C | -0.039 | 6.15E-25 | 1.08 | 4.01E-21 | 1.12 | 5.79E-40 | 1.10 | 9.39E-22 |
| 11q22.3 | 108357137 | rs74911261 | A | G | 0.037 | 1.95E-03 | 1.15 | 1.05E-08 | 1.11 | 3.98E-04 | 1.13 | 1.58E-06 |
| 11q23.2 | 113807181 | rs11214775 | G | A | 0.008 | 0.063 | 1.07 | 1.88E-16 | 1.06 | 3.21E-10 | 1.07 | 4.68E-14 |
| 11q24.2 | 125144426 | rs117015177 | G | A | 0.058 | 2.38E-04 | 1.20 | 1.69E-08 | 1.13 | 1.40E-03 | 1.17 | 5.11E-06 |
| 11q25 | 134266372 | rs878987 | G | A | -0.002 | 0.68 | 1.07 | 1.87E-10 | 1.07 | 7.37E-09 | 1.07 | 2.99E-10 |
| 12p13.1 | 12871099 | rs2066827 | T | G | 0.011 | 0.040 | 1.05 | 4.42E-09 | 1.04 | 1.64E-04 | 1.05 | 4.48E-07 |
| 12p13.1 | 14416918 | rs10845938 | G | A | 0.001 | 0.74 | 1.06 | 2.60E-15 | 1.06 | 2.19E-11 | 1.06 | 2.68E-14 |

| | | | | | | | | | | | | |
|---|---|---|---|---|---|---|---|---|---|---|---|---|
| 12q13.11 | 48419618 | rs80130819 | A | C | -0.004 | 0.52 | 1.10 | 1.05E-11 | 1.10 | 5.71E-10 | 1.10 | 1.67E-11 |
| 12q13.12 | 49676010 | rs10875943 | C | T | 0.000 | 0.99 | 1.08 | 1.49E-19 | 1.08 | 9.20E-15 | 1.08 | 2.20E-18 |
| 12q13.13 | 53312612 | rs73110464 | T | C | 0.009 | 0.13 | 1.18 | 4.82E-51 | 1.17 | 3.95E-33 | 1.17 | 5.86E-45 |
| 12q14.2 | 65012824 | rs7968403 | T | C | -0.003 | 0.52 | 1.06 | 7.22E-14 | 1.06 | 1.05E-11 | 1.06 | 1.42E-13 |
| 12q21.33 | 90192866 | rs4842503 | G | A | 0.004 | 0.39 | 1.06 | 3.78E-12 | 1.05 | 2.60E-08 | 1.06 | 7.56E-11 |
| 12q24.21 | 114666202 | rs10774740 | G | T | 0.013 | 1.36E-03 | 1.08 | 3.77E-22 | 1.06 | 1.23E-11 | 1.07 | 2.69E-17 |
| 12q24.33 | 133067989 | rs7295014 | G | A | 0.015 | 4.68E-04 | 1.05 | 1.69E-09 | 1.03 | 8.67E-04 | 1.04 | 1.44E-06 |
| 13q22.1 | 73719777 | rs9543236 | T | C | 0.012 | 0.015 | 1.07 | 1.55E-11 | 1.05 | 2.59E-06 | 1.06 | 2.78E-09 |
| 13q22.1 | 73995877 | rs7336001 | G | C | 0.038 | 5.88E-08 | 1.10 | 2.27E-11 | 1.06 | 1.07E-03 | 1.08 | 5.14E-07 |
| 14q11.2 | 23305649 | rs1004030 | T | C | 0.006 | 0.15 | 1.05 | 8.17E-10 | 1.04 | 4.31E-06 | 1.05 | 2.18E-08 |
| 14q13.3 | 37137280 | rs12893199 | C | T | 0.007 | 0.072 | 1.06 | 4.27E-13 | 1.05 | 8.33E-08 | 1.05 | 4.97E-11 |
| 14q22.1 | 53375302 | rs7141054 | A | C | 0.011 | 0.023 | 1.09 | 6.27E-18 | 1.07 | 2.00E-10 | 1.08 | 8.59E-15 |
| 14q24.1 | 69027523 | rs34882439 | G | A | 0.004 | 0.31 | 1.06 | 1.11E-10 | 1.05 | 3.93E-07 | 1.05 | 2.10E-09 |
| 14q24.1 | 69134264 | rs767127 | G | A | 0.003 | 0.52 | 1.05 | 2.69E-12 | 1.05 | 1.03E-08 | 1.05 | 2.97E-11 |
| 14q24.2 | 71091142 | rs11158871 | C | T | -0.003 | 0.40 | 1.05 | 1.23E-08 | 1.05 | 7.15E-08 | 1.05 | 9.08E-09 |
| 15q13.3 | 33417586 | rs1902149 | G | A | -0.004 | 0.32 | 1.04 | 4.84E-08 | 1.05 | 1.97E-07 | 1.05 | 4.00E-08 |
| 15q15.1 | 40922915 | rs4924487 | C | G | 0.015 | 3.18E-03 | 1.07 | 3.03E-10 | 1.05 | 7.48E-05 | 1.06 | 1.16E-07 |
| 15q21.3 | 56385868 | rs33984059 | A | G | -0.008 | 0.51 | 1.19 | 1.62E-09 | 1.20 | 1.16E-08 | 1.20 | 1.66E-09 |
| 15q22.31 | 66705043 | rs80326387 | A | G | 0.004 | 0.35 | 1.05 | 2.13E-09 | 1.05 | 2.16E-06 | 1.05 | 2.10E-08 |
| 16q12.2 | 54473072 | rs1420285 | C | T | 0.006 | 0.13 | 1.05 | 1.47E-08 | 1.04 | 2.82E-05 | 1.04 | 3.14E-07 |
| 16q21 | 57654576 | rs11863709 | C | T | -0.002 | 0.86 | 1.15 | 1.43E-11 | 1.16 | 2.40E-09 | 1.15 | 4.11E-11 |
| 16q21 | 57693055 | rs16958674 | C | T | 0.013 | 0.011 | 1.06 | 2.14E-09 | 1.04 | 1.75E-04 | 1.05 | 4.11E-07 |
| 16q23.3 | 82166181 | rs8052913 | C | T | 0.017 | 1.27E-05 | 1.04 | 3.71E-08 | 1.02 | 0.011 | 1.03 | 3.50E-05 |
| 17p13.3 | 618965 | rs684232 | C | T | 0.008 | 0.053 | 1.09 | 1.77E-26 | 1.08 | 2.26E-16 | 1.08 | 1.10E-22 |
| 17p13.1 | 7571752 | rs78378222 | G | T | 0.112 | 4.08E-10 | 1.25 | 4.78E-11 | 1.10 | 0.013 | 1.19 | 6.80E-06 |
| 17p13.1 | 7803118 | rs28441558 | C | T | 0.042 | 3.34E-07 | 1.16 | 3.51E-19 | 1.11 | 1.05E-07 | 1.14 | 7.97E-13 |
| 17q11.2 | 30098749 | rs142444269 | C | T | 0.010 | 0.096 | 1.07 | 1.45E-11 | 1.06 | 2.18E-06 | 1.06 | 1.72E-09 |
| 17q12 | 36047417 | rs3110641 | A | G | 0.011 | 0.025 | 1.07 | 1.24E-12 | 1.05 | 8.73E-07 | 1.06 | 3.65E-10 |
| 17q12 | 36074979 | rs11649743 | G | A | 0.014 | 3.21E-03 | 1.13 | 7.16E-38 | 1.11 | 2.58E-22 | 1.12 | 3.61E-31 |
| 17q12 | 36103565 | rs11263763 | A | G | 0.049 | 3.13E-38 | 1.23 | 3.63E-162 | 1.16 | 5.06E-65 | 1.20 | 3.71E-70 |
| 17q12 | 36229129 | rs8081051 | A | C | 0.007 | 0.22 | 1.05 | 1.35E-08 | 1.04 | 9.01E-05 | 1.05 | 3.84E-07 |

| | | | | | | | | | | | | |
|---|---|---|---|---|---|---|---|---|---|---|---|---|
| 17q21.32 | 45993959 | rs199905768 | A | C | 0.020 | 0.51 | 1.19 | 3.79E-09 | 1.17 | 8.14E-04 | 1.18 | 6.79E-07 |
| 17q21.32 | 46546346 | rs12944450 | T | C | -0.025 | 0.18 | 1.14 | 7.44E-09 | 1.17 | 3.14E-07 | 1.15 | 7.58E-09 |
| 17q21.32 | 46820676 | rs117576373 | T | C | 0.009 | 0.34 | 1.21 | 4.88E-24 | 1.20 | 8.71E-17 | 1.20 | 8.32E-22 |
| 17q21.33 | 47451749 | rs9893373 | G | A | -0.003 | 0.58 | 1.05 | 1.93E-08 | 1.05 | 5.65E-07 | 1.05 | 3.13E-08 |
| 17q21.33 | 47455752 | rs73324348 | A | G | 0.011 | 0.13 | 1.11 | 5.81E-14 | 1.10 | 8.02E-09 | 1.10 | 4.51E-12 |
| 17q22 | 56456120 | rs2680708 | G | A | 0.014 | 3.57E-04 | 1.05 | 5.64E-09 | 1.03 | 1.07E-03 | 1.04 | 2.59E-06 |
| 17q24.3 | 69115358 | rs9911515 | A | G | 0.002 | 0.53 | 1.17 | 7.26E-100 | 1.17 | 8.68E-73 | 1.17 | 3.78E-93 |
| 18q21.2 | 51771322 | rs8089411 | C | T | 0.015 | 6.59E-05 | 1.04 | 1.98E-08 | 1.03 | 3.83E-03 | 1.04 | 1.18E-05 |
| 18q21.2 | 53230859 | rs28607662 | C | T | 0.010 | 0.14 | 1.07 | 2.68E-08 | 1.06 | 5.14E-05 | 1.07 | 5.22E-07 |
| 18q21.32 | 56744666 | rs12327532 | G | T | 0.013 | 1.70E-03 | 1.04 | 4.38E-08 | 1.03 | 1.66E-03 | 1.04 | 7.77E-06 |
| 18q21.33 | 60998567 | rs2850766 | C | G | -0.004 | 0.53 | 1.05 | 3.83E-08 | 1.05 | 6.16E-06 | 1.05 | 8.58E-08 |
| 18q22.3 | 73036165 | rs10460109 | T | C | 0.001 | 0.78 | 1.04 | 1.56E-08 | 1.04 | 1.85E-06 | 1.04 | 5.31E-08 |
| 18q23 | 76770820 | rs9959454 | A | G | 0.025 | 3.29E-09 | 1.08 | 7.09E-22 | 1.05 | 9.76E-08 | 1.07 | 1.74E-13 |
| 19p13.11 | 17214073 | rs11666569 | C | T | 0.009 | 0.033 | 1.05 | 3.61E-09 | 1.04 | 4.23E-05 | 1.05 | 1.84E-07 |
| 19q13.13 | 38555154 | rs35486025 | T | C | -0.0004 | 0.94 | 1.06 | 8.95E-10 | 1.06 | 7.83E-08 | 1.06 | 2.61E-09 |
| 19q13.2 | 38744733 | rs12610267 | A | G | 0.010 | 0.010 | 1.10 | 1.24E-36 | 1.09 | 5.04E-22 | 1.09 | 9.44E-31 |
| 19q13.2 | 41985931 | rs11673591 | T | A | -0.005 | 0.28 | 1.09 | 1.32E-23 | 1.10 | 2.88E-20 | 1.09 | 3.53E-23 |
| 19q13.33 | 51340794 | rs2659053 | A | G | 0.082 | 4.35E-92 | 1.06 | 9.76E-15 | 0.97 | 2.37E-04 | 1.02 | 0.12 |
| 19q13.33 | 51362715 | rs76765083 | T | G | 0.366 | 6.0E-604 | 1.35 | 4.99E-90 | 0.89 | 7.21E-12 | 1.14 | 0.013 |
| 20q11.22 | 34003073 | rs142570322 | C | T | 0.023 | 3.49E-04 | 1.04 | 2.60E-08 | 1.02 | 0.096 | 1.03 | 2.50E-04 |
| 20q13.13 | 49548807 | rs73909841 | T | C | 0.019 | 8.71E-03 | 1.11 | 3.28E-12 | 1.08 | 1.88E-06 | 1.10 | 1.14E-09 |
| 20q13.2 | 52455205 | rs6091758 | G | A | 0.009 | 0.15 | 1.07 | 1.59E-17 | 1.06 | 1.09E-07 | 1.06 | 2.36E-13 |
| 20q13.33 | 61015240 | rs34585497 | A | C | 0.015 | 4.87E-04 | 1.04 | 2.05E-08 | 1.03 | 3.30E-03 | 1.04 | 1.05E-05 |
| 20q13.33 | 62272248 | rs2259797 | T | C | 0.025 | 5.78E-04 | 1.08 | 1.74E-08 | 1.05 | 1.77E-03 | 1.07 | 5.82E-06 |
| 20q13.33 | 62310240 | rs62207046 | C | A | 0.007 | 0.41 | 1.09 | 3.73E-09 | 1.08 | 2.23E-05 | 1.08 | 8.80E-08 |
| 20q13.33 | 62374389 | rs1058319 | C | T | 0.016 | 5.10E-03 | 1.12 | 1.82E-23 | 1.10 | 1.74E-13 | 1.11 | 3.44E-19 |
| 21q22.3 | 42866332 | rs61735792 | A | G | 0.006 | 0.75 | 1.26 | 6.95E-14 | 1.26 | 9.43E-10 | 1.26 | 1.04E-12 |
| 21q22.3 | 42889410 | rs112714648 | A | G | -0.002 | 0.67 | 1.06 | 1.66E-13 | 1.07 | 3.41E-11 | 1.06 | 3.22E-13 |
| 22q11.21 | 19749525 | rs1978060 | G | A | 0.009 | 0.080 | 1.06 | 1.05E-11 | 1.04 | 1.26E-05 | 1.05 | 3.73E-09 |
| 22q12.1 | 28888939 | rs9625483 | A | G | 0.023 | 0.087 | 1.14 | 3.82E-09 | 1.11 | 7.42E-05 | 1.13 | 2.45E-07 |
| 22q13.1 | 40499103 | rs34419824 | T | A | 0.001 | 0.80 | 1.08 | 2.54E-15 | 1.07 | 2.41E-11 | 1.08 | 3.04E-14 |

| | | | | | | | | | | | | |
|---|---|---|---|---|---|---|---|---|---|---|---|---|
| 22q13.2 | 43500212 | rs5759167 | G | T | 0.007 | 0.058 | 1.15 | 3.38E-78 | 1.14 | 4.15E-53 | 1.15 | 6.09E-70 |
| 22q13.2 | 43502152 | rs2007886 | T | C | 0.016 | 0.011 | 1.10 | 2.51E-14 | 1.08 | 4.48E-08 | 1.09 | 1.21E-11 |

[1] Index variants were selected using LD-based clumping: $r^2$<0.01 within ±10 Mb

[2] Per allele change in log(PSA)

[3] Bias-corrected estimated were obtained as follows: $\beta^{adj} = \beta - b * \beta_{PSA}$ and the corresponding $OR^{adj} = \exp(\beta^{adj})$ where $b$ is estimated using the Dudbridge method

[4] Bias-corrected estimated were obtained as follows: $\beta^{adj} = \beta - b * \beta_{PSA}$ and the corresponding $OR^{adj} = \exp(\beta^{adj})$ where $b$ is estimated using SlopeHunter

**Supplementary Table 11: Development and validation of a polygenic score for PSA levels.** Associations with baseline PSA in cancer-free participants from the Prostate Cancer Prevention Trial (PCPT) and Selenium and Vitamin E Cancer Prevention Trial (SELECT) were estimated using linear regression models. All p-values are two-sided. Effect estimates are presented for a 128-variant PSA polygenic score ($PGS_{128}$) developed based on clumping and thresholding (LD $r^2<0.01$, $P<5\times10^{-8}$) and a genome-wide polygenic score with continuous shrinkage priors ($PGS_{CSx}$) comprised of 1,058,173 and 1,071,278 variants in PCPT and SELECT, respectively.

| Study | Ancestry[1] | N | $PGS_{128}$ | | | | $PGS_{CSx}$ | | | |
|---|---|---|---|---|---|---|---|---|---|---|
| | | | $\beta_{PGS}$[2] | (SE) | P | (%) | $\beta_{PGS}$[2] | (SE) | P | (%) |
| PCPT | Pooled[3] | 5,883 | 0.168 | (0.008) | $1.61\times10^{-98}$ | 7.16 | 0.186 | (0.008) | $3.26\times10^{-112}$ | 8.13 |
| | EUR | 5,725 | 0.169 | (0.008) | $5.29\times10^{-98}$ | 7.33 | 0.194 | (0.008) | $1.69\times10^{-115}$ | 8.60 |
| | AFR/EUR | 103 | 0.088 | (0.080) | 0.27 | - | 0.029 | (0.055) | 0.59 | - |
| | EAS | 55 | 0.104 | (0.126) | 0.41 | - | 0.062 | (0.109) | 0.57 | - |
| SELECT | Pooled[3] | 25,917 | 0.207 | (0.004) | $6.69\times10^{-505}$ | 8.00 | 0.258 | (0.005) | $1.25\times10^{-619}$ | 9.61 |
| | EUR | 22,173 | 0.213 | (0.005) | $4.24\times10^{-478}$ | 8.78 | 0.283 | (0.005) | $5.51\times10^{-610}$ | 10.94 |
| | AFR pooled | 2,936 | 0.154 | (0.015) | $1.97\times10^{-24}$ | 3.36 | 0.134 | (0.014) | $9.39\times10^{-23}$ | 3.11 |
| | AFR/EUR | 1,763 | 0.146 | (0.018) | $3.00\times10^{-15}$ | 3.32 | 0.157 | (0.017) | $4.77\times10^{-19}$ | 4.22 |
| | AFR | 1,173 | 0.163 | (0.025) | $8.22\times10^{-11}$ | 3.45 | 0.098 | (0.022) | $8.04\times10^{-6}$ | 1.64 |
| | EAS pooled | 578 | 0.190 | (0.032) | $7.24\times10^{-9}$ | 5.60 | 0.283 | (0.033) | $4.94\times10^{-17}$ | 11.39 |
| | EAS/EUR | 321 | 0.229 | (0.042) | $8.46\times10^{-8}$ | 8.45 | 0.315 | (0.044) | $5.15\times10^{-12}$ | 13.61 |
| | EAS | 257 | 0.136 | (0.054) | 0.012 | 2.45 | 0.258 | (0.050) | $5.94\times10^{-7}$ | 9.22 |

[1] Individuals were assigned to a global ancestry cluster if their ancestry score exceeded 0.80 for a single group. Individuals with ancestry scores that were >0.20 and <0.80 for at least one ancestry were designated to admixed clusters. The AFR/EUR admixed cluster includes participants with 0.20<AFR<0.80 and EAS/EUR includes participants with 0.20<EAS<0.80. AFR pooled combines the AFR and AFR/EUR clusters. EAS pooled combines the EAS and EAS/EUR clusters. Up to 10 genetic ancestry principal components were calculated within each global ancestry cluster.

[2] Effect per standard deviation increase in the standardized PGS on baseline log(PSA), derived from a linear regression model adjusted for age at PSA measurement and the top 10 population-specific genetic ancestry principal components.

[3] Global genetic ancestry proportions (AFR, EAS) were included as covariates in addition to the top 10 population-specific principal components in the pooled analysis including all ancestries

**Supplementary Table 12: Polygenic score associations with measures of temporal change in PSA levels.** Associations with metrics of temporal change in PSA were estimated in participants from the Prostate Cancer Prevention Trial (PCPT) and Selenium and Vitamin E Cancer Prevention Trial (SELECT) using linear regression. All p-values are two-sided. Temporal phenotypes were derived using PSA measures in men without prostate cancer or pre-diagnostic PSA values for men who were diagnosed with prostate cancer during the follow up period. Effect estimates are presented for a 128-variant PSA polygenic score ($PGS_{128}$) developed based on clumping and thresholding (LD $r^2$<0.01, $P<5\times10^{-8}$) and a genome-wide polygenic score with continuous shrinkage priors ($PGS_{CSx}$) comprised of 1,058,173 and 1,071,278 variants in PCPT and SELECT, respectively. Pooled analyses included individuals of all ancestries.

| Phenotype[1,2] | | Predictor | SELECT | | | | PCPT | | | |
|---|---|---|---|---|---|---|---|---|---|---|
| | | | N | β[3] | (SE) | P | N | β[3] | (SE) | P |
| PSA Increase | Velocity | $PGS_{128}$ | 14908 | $-3.30\times10^{-4}$ | $(8.6\times10^{-5})$ | $1.3\times10^{-4}$ | 2933 | $-2.34\times10^{-4}$ | $(1.2\times10^{-4})$ | 0.052 |
| | | $PGS_{CSx}$ | | $-4.06\times10^{-4}$ | $(9.8\times10^{-5})$ | $3.7\times10^{-5}$ | | $-2.65\times10^{-4}$ | $(1.3\times10^{-4})$ | 0.034 |
| | log(Velocity) | $PGS_{128}$ | | -0.033 | (0.007) | $3.2\times10^{-6}$ | | -0.039 | (0.015) | $6.8\times10^{-3}$ |
| | | $PGS_{CSx}$ | | -0.041 | (0.008) | $4.4\times10^{-7}$ | | -0.048 | (0.015) | $1.5\times10^{-3}$ |
| | Doubling time | $PGS_{128}$ | | 8.44 | (1.62) | $2.0\times10^{-7}$ | | 10.25 | (3.14) | $1.1\times10^{-3}$ |
| | | $PGS_{CSx}$ | | 10.37 | (1.85) | $2.2\times10^{-8}$ | | 12.88 | (3.25) | $7.2\times10^{-5}$ |
| PSA Decrease | Velocity | $PGS_{128}$ | 6970 | $3.08\times10^{-4}$ | $(2.3\times10^{-4})$ | 0.21 | 1728 | $-4.70\times10^{-5}$ | $(1.5\times10^{-4})$ | 0.75 |
| | | $PGS_{CSx}$ | | $5.02\times10^{-4}$ | $(2.7\times10^{-4})$ | 0.067 | | $-7.92\times10^{-5}$ | $(1.5\times10^{-4})$ | 0.61 |
| Any PSA change | Velocity | $PGS_{128}$ | 21878 | $3.22\times10^{-6}$ | $(3.7\times10^{-6})$ | 0.39 | 4745 | $-7.95\times10^{-6}$ | $(4.7\times10^{-5})$ | 0.089 |
| | | $PGS_{CSx}$ | | $3.50\times10^{-6}$ | $(4.2\times10^{-6})$ | 0.41 | | $-1.02\times10^{-5}$ | $(4.9\times10^{-6})$ | 0.035 |
| | Doubling time | $PGS_{128}$ | | 4.79 | (1.92) | 0.013 | | 3.33 | 3.59 | 0.35 |
| | | $PGS_{CSx}$ | | 3.65 | (2.18) | 0.094 | | 3.44 | 3.73 | 0.36 |

[1] PSA velocity was calculated as the difference between the first and last log(PSA) values divided by the elapsed time in months: last[log(PSA)] - first[log(PSA)] / elapsed time. Due to the highly skewed velocity distribution estimates are also presented for log-transformed velocity.

[2] PSA doubling time calculated as: log(2)/(PSA velocity). Associations with negative doubling time are not reported, but negative values were included in the combined analysis of any (positive or negative) change in PSA levels.

[3] Associations were estimated per SD increase in PGS using linear regression with adjustment for age at baseline, randomization arm, top 10 population-specific genetic ancestry principal components, and global ancestry proportions (AFR, ASN)

**Supplementary Table 13: Impact of index bias correction on associations between genetics scores for PSA levels and prostate cancer.** Associations between genetic scores for PSA and prostate cancer were estimated using linear regression models in men of European ancestry from the UK Biobank who were excluded from the PSA GWAS. All p-values are two-sided. For PSA, a genome-wide polygenic score with continuous shrinkage priors ($PGS_{CSx}$) comprised of 1,197,312 variants was used. For prostate cancer, the 269-variant polygenic risk score ($PGS_{269}$), as described in Conti et al.[32] was used. Associations were compared with two versions of the bias-corrected risk scores, one with risk allele weights corrected using the Dudbridge method[33] ($PGS_{269}^{adj}$) and another version of the score with risk allele weights derived using SlopeHunter[35] ($PGS_{269}^{adj\text{-}S}$).

| Sample size | | Original ($PGS_{269}$) | | | Bias-corrected ($PGS_{269}^{adj}$) | | | Bias-corrected ($PGS_{269}^{adj\text{-}S}$) | | |
|---|---|---|---|---|---|---|---|---|---|---|
| | | β[1] | (SE) | P | β[2] | (SE) | P | β[3] | (SE) | P |
| Combined | 164,452 | 0.241 | (0.002) | $2.7\times10^{-2174}$ | 0.063 | (0.002) | $2.1\times10^{-144}$ | 0.176 | (0.002) | $2.3\times10^{-1128}$ |
| Cases | 11,568 | 0.190 | (0.009) | $2.3\times10^{-96}$ | 0.029 | (0.009) | $2.7\times10^{-3}$ | 0.130 | (0.009) | $1.1\times10^{-45}$ |
| Controls | 152,884 | 0.236 | (0.003) | $6.7\times10^{-1881}$ | 0.052 | (0.003) | $2.2\times10^{-89}$ | 0.169 | (0.003) | $1.5\times10^{-948}$ |

[1] Effect estimate per standard deviation increase in $PGS_{269}$ on $PGS_{PSA}$, estimated using a linear regression model with adjustment genetic ancestry principal components and genotyping array

[2] Effect estimate per standard deviation increase in $PGS_{269}^{adj}$ on $PGS_{PSA}$, where $PGS_{269}^{adj}$ has bias-corrected risk allele weights based on the Dudbridge estimate of index event bias. Linear regression model was adjusted for the top 10 genetic ancestry principal components and genotyping array

[3] Effect estimate per standard deviation increase in $PGS_{269}^{adj\text{-}S}$ on $PGS_{PSA}$, where $PGS_{269}^{adj}$ has bias-corrected risk allele weights based on the SlopeHunter estimate of index event bias. Linear regression model was adjusted for the top 10 genetic ancestry principal components and genotyping array

**Supplementary Table 14: Impact of index bias corrections on associations with prostate cancer, overall and stratified by Gleason score.** Associations with prostate cancer status were compared for the 269-variant genetic risk score ($PGS_{269}$), as described in Conti & Darst et al. (2021)[32], and two versions of this score calculated with weights corrected for index event bias using the Dudbridge et al. method[33] ($PGS_{269}^{adj}$) and SlopeHunter[35] ($PGS_{269}^{adj\text{-}S}$). Associations were estimated in Genetic Epidemiology Research in Adult Health and Aging (GERA) cases (n=3673) and controls (n=2363) of European ancestry who underwent a prostate biopsy.

| | Predictor | Association Estimates | | | | | |
|---|---|---|---|---|---|---|---|
| | | OR[1,2] | (95% CI) | P | AUC[3] | (95% CI) | $P_{AUC}$[4] |
| All cases | $PGS_{269}$ | 2.71 | (2.28 – 3.21) | $2.85\times10^{-30}$ | 0.677 | (0.634 – 0.719) | - |
| | $PGS_{269}^{adj}$ | 3.63 | (3.01 – 4.37) | $4.87\times10^{-42}$ | 0.685 | (0.643 – 0.727) | $3.91\times10^{-3}$ |
| | $PGS_{269}^{adj\text{-}S}$ | 3.30 | (2.76 – 3.95) | $4.53\times10^{-39}$ | 0.683 | (0.640 – 0.723) | $9.77\times10^{-3}$ |
| Gleason score <7 | $PGS_{269}$ | 3.00 | (2.49 – 3.61) | $1.42\times10^{-30}$ | 0.680 | (0.629 – 0.731) | - |
| | $PGS_{269}^{adj}$ | 3.78 | (3.09 – 4.63) | $5.08\times10^{-38}$ | 0.686 | (0.635 – 0.736) | 0.064 |
| | $PGS_{269}^{adj\text{-}S}$ | 3.57 | (2.94 – 4.34) | $2.03\times10^{-37}$ | 0.686 | (0.635 – 0.736) | 0.151 |
| Gleason score ≥7 | $PGS_{269}$ | 2.36 | (1.92 – 2.91) | $1.15\times10^{-15}$ | 0.678 | (0.619 – 0.736) | - |
| | $PGS_{269}^{adj}$ | 3.58 | (2.87 – 4.47) | $8.53\times10^{-30}$ | 0.692 | (0.634 – 0.750) | $1.91\times10^{-3}$ |
| | $PGS_{269}^{adj\text{-}S}$ | 3.02 | (2.43 – 3.75) | $1.27\times10^{-23}$ | 0.688 | (0.629 – 0.744) | $3.91\times10^{-3}$ |

1 Odds ratios (OR) were estimated for a dichotomized PGS comparing the top decile to the rest

2 Logistic regression models were adjusted for the top 10 genetic ancestry principal components and a combined variable for genotyping array and DNA collection kit (chip-kit)

3 AUC estimates and bootstrapped confidence intervals were obtained using 10-fold cross-validation

4 Paired Wilcoxon rank sum test was used to compare AUC distributions across 10 folds

**Supplementary Table 15: Case-only associations with prostate cancer aggressiveness.** Prostate cancer patients were stratified based on disease aggressiveness as indicated by the Gleason score. Associations were estimated for a genome-wide PSA polygenic score comprised of 1,255,848 variants ($PGS_{CSx}$), an established 269-variant prostate cancer genetic risk score ($PGS_{269}$) as described in Conti & Darst et al. (2021), and and two versions of this score calculated with weights corrected for index event bias using the Dudbridge et al. method[33] ($PGS_{269}^{adj}$) and SlopeHunter[35] ($PGS_{269}^{adj\text{-}S}$). Two-sided p-values, odds ratios (OR), and corresponding 95% confidence intervals were estimated using multinomial logistic regression. Analyses were performed in 4584 prostate cancer cases of European ancestry from the Genetic Epidemiology Research in Adult Health and Aging (GERA) cohort.

| | N | (%) | Predictor | Continuous PGS[1,2] | | | Dichotomized PGS[2,3] | | |
|---|---|---|---|---|---|---|---|---|---|
| | | | | OR | (95% CI) | P | OR | (95% CI) | P |
| Gleason score ≤6 | 2798 | (61.0) | - | 1.00 | - | - | 1.00 | - | - |
| Gleason score = 7 | 1366 | (29.8) | $PGS_{PSA}$ | 0.79 | (0.76 – 0.83) | $8.48\times10^{-9}$ | 0.79 | (0.63 – 0.99) | 0.048 |
| | | | $PGS_{269}$ | 0.98 | (0.91 – 1.05) | 0.52 | 0.99 | (0.85 – 1.17) | 0.94 |
| | | | $PGS_{269}^{adj}$ | 1.03 | (0.97 – 1.11) | 0.35 | 1.05 | (0.86 – 1.23) | 0.58 |
| | | | $PGS_{269}^{adj\text{-}S}$ | 1.00 | (0.93 – 1.07) | 0.95 | 1.05 | (0.89 – 1.23) | 0.56 |
| Gleason score ≥8 | 420 | (9.2) | $PGS_{PSA}$ | 0.71 | (0.64 – 0.81) | $1.61\times10^{-7}$ | 0.73 | (0.49 – 1.07) | 0.11 |
| | | | $PGS_{269}$ | 0.87 | (0.77 – 0.97) | 0.011 | 0.72 | (0.54 – 0.96) | 0.024 |
| | | | $PGS_{269}^{adj}$ | 0.95 | (0.85 – 1.06) | 0.34 | 0.94 | (0.75 – 1.17) | 0.56 |
| | | | $PGS_{269}^{adj\text{-}S}$ | 0.90 | (0.80 – 1.00) | 0.053 | 0.93 | (0.71 – 1.23) | 0.62 |

1 Each PGS was standardized to have a mean of 0 and standard deviation (SD) of 1 in the full GERA cohort, including cases and controls. Odds ratio (OR) were estimated per SD increase in PGS

2 Associations were estimated using multinomial logistic regression with Gleason score as the outcome, operationalized as a three-level ordinal variable. Models for each PGS were adjusted for the top 10 genetic ancestry principal components and a combined variable for genotyping array and DNA collection kit

3 Odds ratios (OR) were estimated for a dichotomized PGS comparing those in the top decile to the rest of the PGS distribution

**Supplementary Table 16: Impact of genetic adjustment of PSA levels on prostate biopsy referral status.** Genetic adjustment of PSA levels was implemented using a multi-ancestry genome-wide PSA polygenic score comprised of 1,255,848 variants ($PGS_{PSA}$). Changes in biopsy eligibility based on genetically adjusted PSA values were assessed for participants who underwent a biopsy in the Genetic Epidemiology Research in Adult Health and Aging (GERA) cohort. Reclassification was assessed with respect to age-specific PSA cut-points used to guide biopsy referral in the Kaiser Permanente health system.

| Ancestry | $PGS_{PSA}$[1] | PSA (ng/mL)[2] | Re-classification Status: Eligible | | Ineligible | | Unchanged | | Total | Net Reclassification[3] | |
|---|---|---|---|---|---|---|---|---|---|---|---|
| **European** | **Mean** | **Mean** | **N** | **(%)** | **N** | **(%)** | **N** | **(%)** | **N** | **%** | **(95% CI)** |
| Cases | 0.278 | 5.67 | 138 | (3.76) | 453 | (12.33) | 3082 | (83.91) | 3673 | -8.58 | (-9.48, -7.67) |
| Gleason score <7 | 0.346 | 4.45 | 73 | (3.59) | 300 | (14.73) | 1663 | (81.68) | 2036 | -11.15 | (-12.52, -9.78) |
| Gleason score ≥7 | 0.115 | 6.96 | 51 | (3.71) | 117 | (8.53) | 1204 | (87.76) | 1372 | -4.81 | (-5.94, -3.68) |
| Gleason score missing | 0.238 | 6.67 | 14 | (5.28) | 36 | (13.58) | 215 | (81.13) | 265 | -8.30 | (-11.62, -4.98) |
| Age at biopsy <65 years | 0.358 | 4.75 | 22 | (1.80) | 179 | (14.67) | 1019 | (83.52) | 1220 | -12.87 | (-14.74, -10.99) |
| Controls | 0.934 | 4.49 | 59 | (2.49) | 753 | (31.87) | 1551 | (65.64) | 2363 | 29.37 | (27.53, 31.21) |
| Age at biopsy <65 years | 1.140 | 4.57 | 8 | (0.85) | 349 | (37.09) | 584 | (62.06) | 941 | 36.23 | (33.17, 39.31) |
| Controls without biopsy | -0.099 | 1.49 | - | - | - | - | - | - | - | - | |
| **African** | **Mean** | **Mean** | **N** | **(%)** | **N** | **(%)** | **N** | **(%)** | **N** | **%** | **(95% CI)** |
| Cases | 0.091 | 3.47 | 12 | (3.06) | 18 | (4.59) | 362 | (92.35) | 392 | -1.53 | (-2.75, -0.32) |
| Gleason score <7 | 0.117 | 3.40 | 11 | (5.34) | 14 | (6.80) | 181 | (87.86) | 206 | -1.46 | (-3.09, 0.18) |
| Gleason score ≥7 | 0.047 | 3.38 | 0 | (0.0) | 3 | (1.79) | 165 | (98.21) | 168 | -1.79 | (-3.79, 0.22) |
| Gleason score missing | 0.096 | 4.82 | 1 | (5.56) | 1 | (5.56) | 16 | (88.89) | 18 | 0 | - |
| Age at biopsy <65 years | 0.151 | 3.33 | 3 | (1.33) | 13 | (5.78) | 209 | (92.89) | 225 | -4.44 | (-7.14, -1.75) |
| Controls | 1.710 | 5.13 | 2 | (1.85) | 75 | (69.44) | 31 | (28.70) | 108 | 66.06 | (58.77, 76.42) |
| Age at biopsy <65 years | 2.009 | 4.74 | 0 | (0.0) | 39 | (82.98) | 8 | (17.02) | 47 | 82.98 | (72.23, 93.73) |
| Controls without biopsy | -0.079 | 1.51 | - | - | - | - | - | - | - | - | |

[1] Mean values of the standardized $PGS_{PSA}$ were calculated in each subset of participants. $PGS_{PSA}$ was transformed to a z-score with mean equal to 0 and standard deviation equal to 1 based on the full GERA cohort. Therefore, a value of ~0.5 is equivalent to a PGS value approximately half SD above the mean

[2] For each participant who underwent a biopsy PSA values closest to the time of biopsy were selected. For controls who did not have a biopsy we first summarized all available PSA values for each participant by taking the median and then calculated the mean PSA value across participants

[3] Net re-classification (NR) was tabulated separately for cases as: $NR_{case}$ = eligible(%) – ineligible(%) and for controls as $NR_{control}$ = ineligible(%) – eligible(%). Confidence intervals for each NR proportion were calculated using the normal approximation: $p \pm 1.96 \times \sqrt{p\,(1-p)/n}$

**Supplementary Table 17: Comparison of logistic regression models for incident prostate cancer in the Prostate Cancer Prevention Trial (PCPT).** Pooled analyses include 335 cases and 5548 controls of all ancestries (European ancestry: 323 cases and 5402 controls). Models are indexed from $M_0$ to $M_8$. Odds ratios (OR), corresponding 95% confidence intervals, and two-sided p-values are presented for PSA variables and polygenic scores (PGS) only. AUC is based on the full model with all covariates.

| **Models** | | **Predictor** | **Association Estimates** | | | | | |
|---|---|---|---|---|---|---|---|---|
| **Pooled (Multi-ancestry)** | | | **OR[1]** | **(95% CI)** | **P** | **AUC** | **(95% CI)** | **$P_{AUC}$[2]** |
| 0 | Baseline age + RCT arm | - | - | - | - | 0.5856 | (0.5542 – 0.6169) | - |
| 1 | Baseline age + RCT arm + baseline PSA | log(PSA) | 1.85 | (1.54 – 2.24) | $1.4\times10^{-10}$ | 0.6323 | (0.6029 – 0.6617) | - |
| 2 | Baseline age + RCT arm + baseline $PSA^G$ | $\log(PSA^G)$ | 1.83 | (1.51 – 2.21) | $4.4\times10^{-10}$ | 0.6443 | (0.6142 – 0.6745) | vs $M_1$ = 0.21 |
| 3 | Baseline age + RCT arm + $PGS_{PSA}$ | $PGS_{PSA}$ | 1.01 | (0.90 – 1.14) | 0.83 | 0.5950 | (0.5642 – 0.6259) | - |
| 4 | Baseline age + RCT arm + $PGS_{269}$ | $PGS_{269}$ | 1.52 | (1.36 – 1.70) | $3.9\times10^{-13}$ | 0.6528 | (0.6232 – 0.6824) | - |
| 5 | Baseline age + RCT arm + $PGS_{269}^{adj}$ | $PGS_{269}^{adj}$ | 1.57 | (1.40 – 1.76) | $7.8\times10^{-15}$ | 0.6561 | (0.6262 – 0.6859) | vs $M_4$ = 0.52 |
| 6 | Baseline age + RCT arm + $PGS_{269}$ + baseline PSA | $PGS_{269}$<br>log(PSA) | 1.47<br>1.72 | (1.31 – 1.65)<br>(1.42 – 2.09) | $3.6\times10^{-11}$<br>$3.7\times10^{-8}$ | 0.6800 | (0.6519 – 0.7082) | vs $M_2$ = $2.9\times10^{-3}$<br>vs $M_4$ = $1.1\times10^{-3}$ |
| 7 | Baseline age + RCT arm + $PGS_{269}$ + baseline $PSA^G$ | $PGS_{269}$<br>$\log(PSA^G)$ | 1.52<br>1.85 | (1.36 – 1.70)<br>(1.52 – 2.24) | $4.3\times10^{-13}$<br>$4.8\times10^{-10}$ | 0.6848 | (0.6566 – 0.7130) | vs $M_4$ = $5.5\times10^{-4}$<br>vs $M_6$ = 0.24 |
| 8 | Baseline age + RCT arm + $PGS_{269}^{adj}$ + baseline $PSA^G$ | $PGS_{269}^{adj}$<br>$\log(PSA^G)$ | 1.55<br>1.81 | (1.39 – 1.74)<br>(1.49 – 2.19) | $3.0\times10^{-14}$<br>$1.6\times10^{-9}$ | 0.6855 | (0.6570 – 0.7139) | vs $M_5$ = $7.5\times10^{-4}$<br>vs $M_7$ = 0.87 |
| **European ancestry** | | | **OR[1]** | **(95% CI)** | **P** | **AUC** | **(95% CI)** | **$P_{AUC}$[2]** |
| 0 | Baseline age + RCT arm | - | - | - | - | 0.5895 | (0.5576 – 0.6213) | - |
| 1 | Baseline age + RCT arm + baseline PSA | log(PSA) | 1.88 | (1.55 – 2.29) | $1.8\times10^{-10}$ | 0.6391 | (0.6094 – 0.6688) | - |
| 2 | Baseline age + RCT arm + baseline $PSA^G$ | $\log(PSA^G)$ | 1.90 | (1.56 – 2.31) | $1.3\times10^{-10}$ | 0.6486 | (0.6187 – 0.6786) | vs $M_1$ = 0.24 |
| 3 | Baseline age + RCT arm + $PGS_{PSA}$ | $PGS_{PSA}$ | 1.01 | (0.90 – 1.14) | 0.86 | 0.6054 | (0.5744 – 0.6364) | - |
| 4 | Baseline age + RCT arm + $PGS_{269}$ | $PGS_{269}$ | 1.53 | (1.36 – 1.72) | $4.6\times10^{-13}$ | 0.6550 | (0.6251 – 0.6848) | - |
| 5 | Baseline age + RCT arm + $PGS_{269}^{adj}$ | $PGS_{269}^{adj}$ | 1.57 | (1.40 – 1.76) | $1.9\times10^{-14}$ | 0.6584 | (0.6282 – 0.6885) | vs $M_4$ = 0.49 |
| 6 | Baseline age + RCT arm + $PGS_{269}$ + baseline PSA | $PGS_{269}$<br>log(PSA) | 1.48<br>1.77 | (1.32 – 1.66)<br>(1.45 – 2.15) | $4.2\times10^{-11}$<br>$1.6\times10^{-8}$ | 0.6817 | (0.6531 – 0.7104) | vs $M_2$ = $5.2\times10^{-3}$<br>vs $M_4$ = $1.9\times10^{-3}$ |
| 7 | Baseline age + RCT arm + $PGS_{269}$ + baseline $PSA^G$ | $PGS_{269}$<br>$\log(PSA^G)$ | 1.53<br>1.92 | (1.37 – 1.73)<br>(1.57 – 2.34) | $4.5\times10^{-13}$<br>$1.2\times10^{-10}$ | 0.6870 | (0.6586 – 0.7155) | vs $M_4$ = $8.2\times10^{-4}$<br>vs $M_6$ = 0.19 |
| 8 | Baseline age + RCT arm + $PGS_{269}^{adj}$ + baseline $PSA^G$ | $PGS_{269}^{adj}$<br>$\log(PSA^G)$ | 1.55<br>1.88 | (1.38 – 1.74)<br>(1.55 – 2.29) | $7.2\times10^{-14}$<br>$4.3\times10^{-10}$ | 0.6875 | (0.6589 – 0.7162) | vs $M_5$ = $1.4\times10^{-3}$<br>vs $M_7$ = 0.90 |

[1] Odds ratios were estimated per standard deviation increase in standardized $PGS_{269}$ and $PGS_{PSA}$. Models with genetic predictors ($PSA^G$, $PGS_{269}$, $PGS_{269}^{adj}$, and $PGS_{PSA}$) were adjusted for the top 10 genetic ancestry principal components and global ancestry proportions in the pooled analysis.

[2] Two-sided p-values based on De Long's test for two correlated ROC curves.

**Supplementary Table 18: Comparison of logistic regression models for incident aggressive prostate cancer in the Prostate Cancer Prevention Trial (PCPT).** Pooled, multi-ancestry analyses include 75 cases and 5548 controls (European ancestry: 71 cases, 5402 controls). Models are indexed from $M_0$ to $M_8$. Odds ratios (OR), corresponding 95% confidence intervals, and two-sided p-values are presented for PSA variables and polygenic scores (PGS) only. AUC is based on the full model with all covariates.

| Models | | Predictor | Association Estimates | | | | | |
|---|---|---|---|---|---|---|---|---|
| **Pooled (Multi-ancestry)** | | | **OR**[1] | **(95% CI)** | **P** | **AUC** | **(95% CI)** | **$P_{AUC}$**[2] |
| 0 | Baseline age + RCT arm | - | - | - | - | 0.5647 | (0.5007 – 0.6287) | - |
| 1 | Baseline age + RCT arm + baseline PSA | log(PSA) | 2.84 | (1.86 – 4.31) | $1.2\times10^{-6}$ | 0.6781 | (0.6190 – 0.7371) | - |
| 2 | Baseline age + RCT arm + baseline $PSA^G$ | log($PSA^G$) | 2.87 | (1.98 – 4.65) | $6.9\times10^{-7}$ | 0.7055 | (0.6451 – 0.7649) | vs $M_1$ = 0.23 |
| 3 | Baseline age + RCT arm + $PGS_{PSA}$ | $PGS_{PSA}$ | 0.98 | (0.77 – 1.25) | 0.89 | 0.5925 | (0.5271 – 0.6579) | - |
| 4 | Baseline age + RCT arm + $PGS_{269}$ | $PGS_{269}$ | 1.46 | (1.16 – 1.84) | $1.5\times10^{-3}$ | 0.6409 | (0.5740 – 0.7077) | - |
| 5 | Baseline age + RCT arm + $PGS_{269}^{adj}$ | $PGS_{269}^{adj}$ | 1.55 | (1.23 – 1.95) | $2.4\times10^{-4}$ | 0.6509 | (0.5836 – 0.7183) | vs $M_4$ = 0.21 |
| 6 | Baseline age + RCT arm + $PGS_{269}$ + baseline PSA | $PGS_{269}$ | 1.37 | (1.08 – 1.73) | $8.7\times10^{-3}$ | 0.7113 | (0.6518 – 0.7708) | vs $M_2$ = 0.035 |
| | | log(PSA) | 2.67 | (1.74 – 4.11) | $8.0\times10^{-5}$ | | | vs $M_4$ = $2.2\times10^{-3}$ |
| 7 | Baseline age + RCT arm + $PGS_{269}$ + baseline $PSA^G$ | $PGS_{269}$ | 1.45 | (1.15 – 1.83) | $1.6\times10^{-3}$ | 0.7238 | (0.6632 – 0.7844) | vs $M_4$ = $4.7\times10^{-4}$ |
| | | log($PSA^G$) | 2.91 | (1.91 – 4.44) | $7.4\times10^{-7}$ | | | vs $M_6$ = 0.18 |
| 8 | Baseline age + RCT arm + $PGS_{269}^{adj}$ + baseline $PSA^G$ | $PGS_{269}^{adj}$ | 1.52 | (1.20 – 1.92) | $4.3\times10^{-4}$ | 0.7264 | (0.6653 – 0.7876) | vs $M_5$ = $8.1\times10^{-4}$ |
| | | log($PSA^G$) | 2.84 | (1.86 – 4.31) | $1.1\times10^{-6}$ | | | vs $M_7$ = 0.58 |
| **European ancestry** | | | **OR**[2] | **(95% CI)** | **P** | **AUC** | **(95% CI)** | **$P_{AUC}$**[3] |
| 0 | Baseline age + RCT arm | - | - | - | - | 0.5807 | (0.5156 – 0.6458) | - |
| 1 | Baseline age + RCT arm + baseline PSA | log(PSA) | 2.80 | (1.81 – 4.31) | $3.2\times10^{-6}$ | 0.6842 | (0.6234 – 0.7450) | - |
| 2 | Baseline age + RCT arm + baseline $PSA^G$ | log($PSA^G$) | 2.99 | (1.95 – 4.59) | $4.8\times10^{-7}$ | 0.7108 | (0.6519 – 0.7697) | vs $M_1$ = 0.061 |
| 3 | Baseline age + RCT arm + $PGS_{PSA}$ | $PGS_{PSA}$ | 0.94 | (0.73 – 1.20) | 0.63 | 0.6139 | (0.5579 – 0.6698) | - |
| 4 | Baseline age + RCT arm + $PGS_{269}$ | $PGS_{269}$ | 1.45 | (1.14 – 1.83) | $2.3\times10^{-3}$ | 0.6453 | (0.5819 – 0.7087) | - |
| 5 | Baseline age + RCT arm + $PGS_{269}^{adj}$ | $PGS_{269}^{adj}$ | 1.55 | (1.22 – 1.96) | $3.0\times10^{-4}$ | 0.6572 | (0.5941 – 0.7203) | vs $M_4$ = 0.22 |
| 6 | Baseline age + RCT arm + $PGS_{269}$ + baseline PSA | $PGS_{269}$ | 1.36 | (1.08 – 1.73) | 0.010 | 0.7093 | (0.6471 – 0.7715) | vs $M_2$ = 0.92 |
| | | log(PSA) | 2.66 | (1.72 – 4.13) | $1.1\times10^{-5}$ | | | vs $M_4$ = $6.3\times10^{-3}$ |
| 7 | Baseline age + RCT arm + $PGS_{269}$ + baseline $PSA^G$ | $PGS_{269}$ | 1.45 | (1.15 – 1.84) | $2.0\times10^{-3}$ | 0.7273 | (0.6659 – 0.7887) | vs $M_4$ = $1.5\times10^{-3}$ |
| | | log($PSA^G$) | 3.05 | (1.98 – 4.71) | $4.5\times10^{-7}$ | | | vs $M_6$ = 0.073 |
| 8 | Baseline age + RCT arm + $PGS_{269}^{adj}$ + baseline $PSA^G$ | $PGS_{269}^{adj}$ | 1.53 | (1.21 – 1.93) | $4.6\times10^{-4}$ | 0.7335 | (0.6722 – 0.7948) | vs $M_5$ = $1.5\times10^{-3}$ |
| | | log($PSA^G$) | 2.97 | (1.93 – 4.57) | $6.8\times10^{-7}$ | | | vs $M_7$ = 0.26 |

[1] Odds ratios were estimated per standard deviation increase in standardized $PGS_{269}$ and $PGS_{PSA}$. Models with genetic predictors ($PSA^G$, $PGS_{269}$, $PGS_{269}^{adj}$, and $PGS_{PSA}$) were adjusted for the top 10 genetic ancestry principal components and global ancestry proportions in the pooled analysis.

[2] P-values based on De Long's test for two correlated ROC curves.

**Supplementary Table 19: Comparison of logistic regression models for incident non-aggressive prostate cancer in the Prostate Cancer Prevention Trial (PCPT).** Pooled, multi-ancestry analyses include 260 cases and 5548 controls (European ancestry: 252 cases, 5402 controls). Models are indexed from $M_0$ to $M_8$. Odds ratios (OR), corresponding 95% confidence intervals, and two-sided p-values are presented for PSA variables and polygenic scores (PGS) only. AUC is based on the full model with all covariates.

| Models | | Predictor | Association Estimates | | | | | |
|---|---|---|---|---|---|---|---|---|
| Pooled (Multi-ancestry) | | | OR[1] | (95% CI) | P | AUC | (95% CI) | $P_{AUC}$[2] |
| 0 | Baseline age + RCT arm | - | - | - | - | 0.5915 | (0.5560 – 0.6270) | - |
| 1 | Baseline age + RCT arm + baseline PSA | log(PSA) | 1.66 | (1.34 – 2.04) | $2.2\times10^{-6}$ | 0.6228 | (0.5895 – 0.6561) | - |
| 2 | Baseline age + RCT arm + baseline $PSA^G$ | log($PSA^G$) | 1.63 | (1.32 – 2.01) | $5.6\times10^{-6}$ | 0.6341 | (0.6000 – 0.6682) | vs $M_1$ = 0.24 |
| 3 | Baseline age + RCT arm + $PGS_{PSA}$ | $PGS_{PSA}$ | 1.02 | (0.90 – 1.16) | 0.77 | 0.6060 | (0.5711 – 0.6408) | - |
| 4 | Baseline age + RCT arm + $PGS_{269}$ | $PGS_{269}$ | 1.54 | (1.36 – 1.75) | $3.1\times10^{-11}$ | 0.6599 | (0.6274 – 0.6924) | - |
| 5 | Baseline age + RCT arm + $PGS_{269}^{adj}$ | $PGS_{269}^{adj}$ | 1.58 | (1.39 – 1.79) | $2.8\times10^{-12}$ | 0.6614 | (0.6285 – 0.6924) | vs $M_4$ = 0.79 |
| 6 | Baseline age + RCT arm + $PGS_{269}$ + baseline PSA | $PGS_{269}$<br>log(PSA) | 1.50<br>1.54 | (1.32 – 1.70)<br>(1.24 – 1.91) | $5.7\times10^{-10}$<br>$7.8\times10^{-5}$ | 0.6786 | (0.6473 – 0.7100) | vs $M_2$ = $2.3\times10^{-3}$<br>vs $M_4$ = 0.014 |
| 7 | Baseline age + RCT arm + $PGS_{269}$ + baseline $PSA^G$ | $PGS_{269}$<br>log($PSA^G$) | 1.54<br>1.65 | (1.36 – 1.75)<br>(1.33 – 2.04) | $2.8\times10^{-11}$<br>$5.2\times10^{-6}$ | 0.6822 | (0.6510 – 0.7134) | vs $M_4$ = 0.010<br>vs $M_6$ = 0.35 |
| 8 | Baseline age + RCT arm + $PGS_{269}^{adj}$ + baseline $PSA^G$ | $PGS_{269}^{adj}$<br>log($PSA^G$) | 1.56<br>1.61 | (1.38 – 1.78)<br>(1.30 – 2.00) | $5.8\times10^{-12}$<br>$1.2\times10^{-5}$ | 0.6814 | (0.6500 – 0.7128) | vs $M_5$ = 0.015<br>vs $M_7$ = 0.87 |
| European ancestry | | | OR[2] | (95% CI) | P | AUC | (95% CI) | $P_{AUC}$[3] |
| 0 | Baseline age + RCT arm | - | - | - | - | 0.5919 | (0.5558 – 0.6280) | - |
| 1 | Baseline age + RCT arm + baseline PSA | log(PSA) | 1.70 | (1.37 – 2.10) | $1.5\times10^{-6}$ | 0.6275 | (0.5939 – 0.6610) | - |
| 2 | Baseline age + RCT arm + baseline $PSA^G$ | log($PSA^G$) | 1.68 | (1.35 – 2.09) | $2.6\times10^{-6}$ | 0.6376 | (0.6033 – 0.6719) | vs $M_1$ = 0.28 |
| 3 | Baseline age + RCT arm + $PGS_{PSA}$ | $PGS_{PSA}$ | 1.03 | (0.90 – 1.18) | 0.65 | 0.6082 | (0.5727 – 0.6437) | - |
| 4 | Baseline age + RCT arm + $PGS_{269}$ | $PGS_{269}$ | 1.55 | (1.36 – 1.77) | $2.5\times10^{-11}$ | 0.6597 | (0.6264 – 0.6930) | - |
| 5 | Baseline age + RCT arm + $PGS_{269}^{adj}$ | $PGS_{269}^{adj}$ | 1.57 | (1.38 – 1.79) | $6.0\times10^{-12}$ | 0.6606 | (0.6268 – 0.6945) | vs $M_4$ = 0.87 |
| 6 | Baseline age + RCT arm + $PGS_{269}$ + baseline PSA | $PGS_{269}$<br>log(PSA) | 1.51<br>1.59 | (1.33 – 1.72)<br>(1.28 – 1.98) | $5.2\times10^{-10}$<br>$3.2\times10^{-5}$ | 0.6788 | (0.6467 – 0.7110) | vs $M_2$ = $4.5\times10^{-3}$<br>vs $M_4$ = 0.019 |
| 7 | Baseline age + RCT arm + $PGS_{269}$ + baseline $PSA^G$ | $PGS_{269}$<br>log($PSA^G$) | 1.55<br>1.70 | (1.37 – 1.77)<br>(1.37 – 2.12) | $2.2\times10^{-11}$<br>$2.3\times10^{-6}$ | 0.6821 | (0.6503 – 0.7139) | vs $M_4$ = 0.013<br>vs $M_6$ = 0.40 |
| 8 | Baseline age + RCT arm + $PGS_{269}^{adj}$ + baseline $PSA^G$ | $PGS_{269}^{adj}$<br>log($PSA^G$) | 1.56<br>1.66 | (1.37 – 1.78)<br>(1.34 – 2.07) | $1.3\times10^{-12}$<br>$5.4\times10^{-6}$ | 0.6810 | (0.6490 – 0.7130) | vs $M_5$ = 0.019<br>vs $M_7$ = 0.82 |

[1] Odds ratios were estimated per standard deviation increase in standardized $PGS_{269}$ and $PGS_{PSA}$. Models with genetic predictors ($PSA^G$, $PGS_{269}$, $PGS_{269}^{adj}$, and $PGS_{PSA}$) were adjusted for the top 10 genetic ancestry principal components and global ancestry proportions in the pooled analysis.

[2] Two-sided p-values based on De Long's test for two correlated ROC curves.

**Supplementary Table 20: Case-only analyses of prostate cancer aggressiveness in the Prostate Cancer Prevention Trial (PCPT).** Comparison of logistic regression models for distinguishing aggressive (Pooled: n=75; European ancestry: n=71) from non-aggressive (Pooled: n=260; European ancestry: n=252) disease. Models are indexed from $M_0$ to $M_8$. Odds ratios (OR), corresponding 95% confidence intervals, and two-sided p-values are presented for PSA variables and polygenic scores (PGS) only. AUC is based on the full model with all covariates.

| Models | | Predictor | Association Estimates | | | | | |
|---|---|---|---|---|---|---|---|---|
| **Pooled (Multi-ancestry)** | | | **OR**[1] | **(95% CI)** | **P** | **AUC** | **(95% CI)** | **$P_{AUC}$**[2] |
| 0 | Baseline age + RCT arm | - | - | - | - | 0.5124 | (0.4360 – 0.5888) | - |
| 1 | Baseline age + RCT arm + baseline PSA | log(PSA) | 1.86 | (1.12 – 3.10) | 0.017 | 0.6033 | (0.5286 – 0.6781) | - |
| 2 | Baseline age + RCT arm + baseline $PSA^G$ | log($PSA^G$) | 2.06 | (1.23 – 3.45) | $5.9\times10^{-3}$ | 0.6456 | (0.5716 – 0.7197) | vs $M_1$ = 0.19 |
| 3 | Baseline age + RCT arm + $PGS_{PSA}$ | $PGS_{PSA}$ | 0.88 | (0.66 – 1.19) | 0.41 | 0.6055 | (0.5285 – 0.6825) | - |
| 4 | Baseline age + RCT arm + $PGS_{269}$ | $PGS_{269}$ | 0.91 | (0.68 – 1.22) | 0.54 | 0.6029 | (0.5259 – 0.6799) | - |
| 5 | Baseline age + RCT arm + $PGS_{269}^{adj}$ | $PGS_{269}^{adj}$ | 0.97 | (0.73 – 1.29) | 0.85 | 0.5944 | (0.5157 – 0.6730) | vs $M_4$ = 0.17 |
| 6 | Baseline age + RCT arm + $PGS_{269}$ + baseline PSA | $PGS_{269}$ | 0.88 | (0.65 – 1.18) | 0.39 | 0.6396 | (0.5656 – 0.7137) | vs $M_2$ = 0.69 |
| | | log(PSA) | 1.94 | (1.14 – 3.28) | 0.014 | | | vs $M_4$ = 0.23 |
| 7 | Baseline age + RCT arm + $PGS_{269}$ + baseline $PSA^G$ | $PGS_{269}$ | 0.93 | (0.69 – 1.25) | 0.63 | 0.6476 | (0.5748 – 0.7205) | vs $M_4$ = 0.16 |
| | | log($PSA^G$) | 2.04 | (1.22 – 3.42) | $6.4\times10^{-3}$ | | | vs $M_6$ = 0.55 |
| 8 | Baseline age + RCT arm + $PGS_{269}^{adj}$ + baseline $PSA^G$ | $PGS_{269}^{adj}$ | 0.98 | (0.73 – 1.31) | 0.88 | 0.6457 | (0.5720 – 0.7195) | vs $M_5$ = 0.11 |
| | | log($PSA^G$) | 2.06 | (1.23 – 3.44) | $5.9\times10^{-3}$ | | | vs $M_7$ = 0.62 |
| **European ancestry** | | | **OR**[2] | **(95% CI)** | **P** | **AUC** | **(95% CI)** | **$P_{AUC}$**[3] |
| 0 | Baseline age + RCT arm | - | - | - | - | 0.5142 | (0.4380 – 0.5904) | - |
| 1 | Baseline age + RCT arm + baseline PSA | log(PSA) | 1.81 | (1.07 – 3.05) | 0.026 | 0.5924 | (0.5156 – 0.6692) | - |
| 2 | Baseline age + RCT arm + baseline $PSA^G$ | log($PSA^G$) | 2.02 | (1.20 – 3.38) | $7.8\times10^{-3}$ | 0.6254 | (0.5505 – 0.7003) | vs $M_1$ = 0.24 |
| 3 | Baseline age + RCT arm + $PGS_{PSA}$ | $PGS_{PSA}$ | 0.87 | (0.64 – 1.17) | 0.34 | 0.5822 | (0.5068 – 0.6575) | - |
| 4 | Baseline age + RCT arm + $PGS_{269}$ | $PGS_{269}$ | 0.90 | (0.67 – 1.21) | 0.50 | 0.5787 | (0.5016 – 0.6558) | - |
| 5 | Baseline age + RCT arm + $PGS_{269}^{adj}$ | $PGS_{269}^{adj}$ | 0.97 | (0.73 – 1.29) | 0.84 | 0.5740 | (0.4954 – 0.6526) | vs $M_4$ = 0.63 |
| 6 | Baseline age + RCT arm + $PGS_{269}$ + baseline PSA | $PGS_{269}$ | 0.87 | (0.65 – 1.18) | 0.38 | 0.6159 | (0.5397 – 0.6920) | vs $M_2$ = 0.60 |
| | | log(PSA) | 1.87 | (1.10 – 3.17) | 0.020 | | | vs $M_4$ = 0.29 |
| 7 | Baseline age + RCT arm + $PGS_{269}$ + baseline $PSA^G$ | $PGS_{269}$ | 0.92 | (0.68 – 1.24) | 0.59 | 0.6249 | (0.5520 – 0.6977) | vs $M_4$ = 0.22 |
| | | log($PSA^G$) | 2.00 | (1.19 – 3.35) | $8.4\times10^{-3}$ | | | vs $M_6$ = 0.57 |
| 8 | Baseline age + RCT arm + $PGS_{269}^{adj}$ + baseline $PSA^G$ | $PGS_{269}^{adj}$ | 0.97 | (0.73 – 1.30) | 0.86 | 0.6245 | (0.5500 – 0.6989) | vs $M_5$ = 0.19 |
| | | log($PSA^G$) | 2.02 | (1.20 – 3.38) | $7.8\times10^{-3}$ | | | vs $M_7$ = 0.94 |

[1] Odds ratios were estimated per standard deviation increase in standardized $PGS_{269}$ and $PGS_{PSA}$. Models with genetic predictors ($PSA^G$, $PGS_{269}$, $PGS_{269}^{adj}$, and $PGS_{PSA}$) were adjusted for the top 10 genetic ancestry principal components and global ancestry proportions in the pooled analysis.

[2] Two-sided p-values based on De Long's test for two correlated ROC curves.

**Supplementary Table 21: Comparison of logistic regression models for incident prostate cancer in the Selenium and Vitamin E Cancer Prevention Trial (SELECT).** Pooled analysis includes 572 cases and 23,667 cancer-free controls of all ancestries. Ancestry-stratified analyses were also performed. Models are indexed from $M_0$ to $M_8$. Odds ratios (OR), corresponding 95% confidence intervals, and two-sided p-values are presented for PSA variables and polygenic scores (PGS) only. AUC is based on the full model with all covariates.

| Models | | Predictor | Association Estimates | | | | | |
|---|---|---|---|---|---|---|---|---|
| **Pooled (Multi-ancestry)** | | | **OR**[1] | **(95% CI)** | **P** | **AUC** | **(95% CI)** | **$P_{AUC}$**[2] |
| 0 | Baseline age + RCT arm | - | - | - | - | 0.5082 | (0.4844 – 0.5321) | - |
| 1 | Baseline age + RCT arm + baseline PSA | log(PSA) | 3.88 | (3.35 – 4.50) | $4.1\times10^{-73}$ | 0.7365 | (0.7176 – 0.7555) | - |
| 2 | Baseline age + RCT arm + baseline $PSA^G$ | log($PSA^G$) | 3.53 | (3.07 – 4.07) | $5.3\times10^{-68}$ | 0.7278 | (0.7080 – 0.7476) | vs $M_1$ = 0.096 |
| 3 | Baseline age + RCT arm + $PGS_{PSA}$ | $PGS_{PSA}$ | 1.23 | (1.12 – 1.35) | $2.4\times10^{-5}$ | 0.5682 | (0.5446 – 0.5917) | - |
| 4 | Baseline age + RCT arm + $PGS_{269}$ | $PGS_{269}$ | 1.74 | (1.59 – 1.90) | $2.9\times10^{-35}$ | 0.6587 | (0.6361 – 0.6813) | - |
| 5 | Baseline age + RCT arm + $PGS_{269}^{adj}$ | $PGS_{269}^{adj}$ | 1.65 | (1.51 – 1.79) | $3.8\times10^{-30}$ | 0.6496 | (0.6269 – 0.6724) | vs $M_4$ = 0.019 |
| 6 | Baseline age + RCT arm + $PGS_{269}$ + baseline PSA | $PGS_{269}$ | 1.58 | (1.44 – 1.73) | $1.0\times10^{-23}$ | 0.7634 | (0.7447 – 0.7820) | vs $M_2$ = $2.7\times10^{-7}$ |
| | | log(PSA) | 3.68 | (3.17 – 4.28) | $3.8\times10^{-65}$ | | | vs $M_4$ = $1.5\times10^{-25}$ |
| 7 | Baseline age + RCT arm + $PGS_{269}$ + baseline $PSA^G$ | $PGS_{269}$ | 1.70 | (1.55 – 1.85) | $1.1\times10^{-31}$ | 0.7606 | (0.7415 – 0.7797) | vs $M_4$ = $4.2\times10^{-24}$ |
| | | log($PSA^G$) | 3.53 | (3.05 – 4.08) | $5.3\times10^{-65}$ | | | vs $M_6$ = 0.48 |
| 8 | Baseline age + RCT arm + $PGS_{269}^{adj}$ + baseline $PSA^G$ | $PGS_{269}^{adj}$ | 1.60 | (1.46 – 1.74) | $2.6\times10^{-26}$ | 0.7561 | (0.7370 – 0.7752) | vs $M_5$ = $2.3\times10^{-24}$ |
| | | log($PSA^G$) | 3.47 | (3.01 – 4.01) | $7.8\times10^{-65}$ | | | vs $M_7$ = 0.011 |
| **European (467 cases 20,173 controls)** | | | **OR**[1] | **(95% CI)** | **P** | **AUC** | **(95% CI)** | **$P_{AUC}$**[2] |
| 0 | Baseline age + RCT arm | - | - | - | - | 0.5035 | (0.4788 – 0.5281) | - |
| 1 | Baseline age + RCT arm + baseline PSA | log(PSA) | 3.98 | (3.38 – 4.69) | $7.9\times10^{-61}$ | 0.7382 | (0.7175 – 0.7588) | - |
| 2 | Baseline age + RCT arm + baseline $PSA^G$ | log($PSA^G$) | 3.85 | (3.27 – 4.53) | $2.1\times10^{-59}$ | 0.7346 | (0.7132 – 0.7561) | vs $M_1$ = 0.55 |
| 3 | Baseline age + RCT arm + $PGS_{PSA}$ | $PGS_{PSA}$ | 1.21 | (1.09 – 1.35) | $5.9\times10^{-4}$ | 0.5702 | (0.5450 – 0.5953) | - |
| 4 | Baseline age + RCT arm + $PGS_{269}$ | $PGS_{269}$ | 1.81 | (1.64 – 2.00) | $8.8\times10^{-33}$ | 0.6670 | (0.6428 – 0.6912) | - |
| 5 | Baseline age + RCT arm + $PGS_{269}^{adj}$ | $PGS_{269}^{adj}$ | 1.70 | (1.55 – 1.87) | $7.9\times10^{-28}$ | 0.6562 | (0.6319 – 0.6804) | vs $M_4$ = 0.015 |
| 6 | Baseline age + RCT arm + $PGS_{269}$ + baseline PSA | $PGS_{269}$ | 1.63 | (1.47 – 1.80) | $6.8\times10^{-22}$ | 0.7671 | (0.7470 – 0.7872) | vs $M_2$ = $1.9\times10^{-5}$ |
| | | log(PSA) | 3.70 | (3.13 – 4.37) | $5.7\times10^{-53}$ | | | vs $M_4$ = $2.7\times10^{-22}$ |
| 7 | Baseline age + RCT arm + $PGS_{269}$ + baseline $PSA^G$ | $PGS_{269}$ | 1.76 | (1.59 – 1.94) | $3.1\times10^{-29}$ | 0.7681 | (0.7471 – 0.7891) | vs $M_4$ = $1.3\times10^{-21}$ |
| | | log($PSA^G$) | 3.85 | (3.26 – 4.55) | $1.8\times10^{-56}$ | | | vs $M_6$ = 0.81 |
| 8 | Baseline age + RCT arm + $PGS_{269}^{adj}$ + baseline $PSA^G$ | $PGS_{269}^{adj}$ | 1.64 | (1.49 – 1.80) | $7.7\times10^{-24}$ | 0.7629 | (0.7419 – 0.7839) | vs $M_5$ = $5.9\times10^{-22}$ |
| | | log($PSA^G$) | 3.46 | (3.19 – 4.43) | $3.6\times10^{-56}$ | | | vs $M_7$ = $9.2\times10^{-3}$ |
| **African**[3] **(88 cases 2733 controls)** | | | **OR**[1] | **(95% CI)** | **P** | **AUC** | **(95% CI)** | **$P_{AUC}$**[2] |
| 0 | Baseline age + RCT arm | - | - | - | - | 0.5808 | (0.5175 – 0.6441) | - |
| 1 | Baseline age + RCT arm + baseline PSA | log(PSA) | 3.63 | (2.54 – 5.20) | $2.0\times10^{-12}$ | 0.7423 | (0.6925 – 0.7921) | - |

| | | | | | | | | |
|---|---|---|---|---|---|---|---|---|
| 2 | Baseline age + RCT arm + baseline $PSA^G$ [$PGS_{CSx}$] | log($PSA^G$) | 2.68 | (1.94 – 3.69) | $1.7\times10^{-9}$ | 0.7266 | (0.6760 – 0.7772) | vs $M_1$ = 0.35 |
| | Baseline age + RCT arm + baseline $PSA^G$ [$PGS_{128}$] | log($PSA^G$) | 3.37 | (2.38 – 4.78) | $7.6\times10^{-12}$ | 0.7500 | (0.7003 – 0.7998) | vs $M_1$ = 0.56 |
| 3 | Baseline age + RCT arm + $PGS_{PSA}$ | $PGS_{PSA}$ | 1.30 | (1.05 – 1.62) | 0.018 | 0.6383 | (0.5784 – 0.6981) | - |
| 4 | Baseline age + RCT arm + $PGS_{269}$ | $PGS_{269}$ | 1.43 | (1.15 – 1.78) | $1.3\times10^{-3}$ | 0.6573 | (0.5950 – 0.7196) | - |
| 5 | Baseline age + RCT arm + $PGS_{269}^{adj}$ | $PGS_{269}^{adj}$ | 1.42 | (1.15 – 1.75) | $1.3\times10^{-3}$ | 0.6638 | (0.6026 – 0.7250) | vs $M_4$ = 0.22 |
| 6 | Baseline age + RCT arm + $PGS_{269}$ + baseline PSA | $PGS_{269}$ | 1.34 | (1.07 – 1.68) | 0.012 | 0.7689 | (0.7202 – 0.8176) | vs $M_2$ = $4.4\times10^{-3}$ |
| | | log(PSA) | 3.63 | (2.52 – 5.25) | $5.7\times10^{-12}$ | | | vs $M_4$ = $3.4\times10^{-5}$ |
| 7 | Baseline age + RCT arm + $PGS_{269}$ + baseline $PSA^G$ [$PGS_{CSx}$] | $PGS_{269}$ | 1.42 | (1.13 – 1.77) | $2.3\times10^{-3}$ | 0.7432 | (0.6934 – 0.7930) | vs $M_4$ = $4.7\times10^{-4}$ |
| | | log($PSA^G$) | 2.66 | (1.93 – 3.68) | $3.1\times10^{-9}$ | | | vs $M_6$ = 0.016 |
| | Baseline age + RCT arm + $PGS_{269}$ + baseline $PSA^G$ [$PGS_{128}$] | $PGS_{269}$ | 1.43 | (1.14 – 1.78) | $1.8\times10^{-3}$ | 0.7636 | (0.7134 – 0.8138) | vs $M_4$ = $8.1\times10^{-5}$ |
| | | log($PSA^G$) | 3.42 | (2.40 – 4.87) | $1.2\times10^{-11}$ | | | vs $M_6$ = 0.54 |
| 8 | Baseline age + RCT arm + $PGS_{269}^{adj}$ + baseline $PSA^G$ | $PGS_{269}^{adj}$ | 1.41 | (1.13 – 1.76) | $2.0\times10^{-3}$ | 0.7460 | (0.6969 – 0.7951) | vs $M_5$ = $7.4\times10^{-4}$ |
| | | log($PSA^G$) | 2.66 | (1.93 – 3.68) | $2.7\times10^{-9}$ | | | vs $M_7$ = 0.33 |
| **East Asian[4] (13 cases 544 controls)** | | | **OR[1]** | **(95% CI)** | **P** | **AUC** | **(95% CI)** | **$P_{AUC}$[2]** |
| 0 | Baseline age + RCT arm | - | - | - | - | 0.6361 | (0.4532 – 0.8190) | - |
| 1 | Baseline age + RCT arm + baseline PSA | log(PSA) | 2.60 | (1.03 – 6.54) | 0.043 | 0.7039 | (0.5673 – 0.8405) | - |
| 2 | Baseline age + RCT arm + baseline $PSA^G$ | log($PSA^G$) | 2.15 | (0.82 – 5.62) | 0.12 | 0.8271 | (0.7104 – 0.9438) | vs $M_1$ = 0.011 |
| 3 | Baseline age + RCT arm + $PGS_{PSA}$ | $PGS_{PSA}$ | 1.54 | (0.76 – 3.12) | 0.23 | 0.8183 | (0.6961 – 0.9405) | - |
| 4 | Baseline age + RCT arm + $PGS_{269}$ | $PGS_{269}$ | 2.07 | (0.97 – 4.41) | 0.059 | 0.8334 | (0.7120 – 0.9548) | - |
| 5 | Baseline age + RCT arm + $PGS_{269}^{adj}$ | $PGS_{269}^{adj}$ | 1.62 | (0.81 – 3.24) | 0.17 | 0.8262 | (0.7046 – 0.9478) | vs $M_4$ = 0.49 |
| 6 | Baseline age + RCT arm + $PGS_{269}$ + baseline PSA | $PGS_{269}$ | 2.07 | (0.97 – 4.38) | 0.059 | 0.8501 | (0.7453 – 0.9550) | vs $M_2$ = 0.099 |
| | | log(PSA) | 2.91 | (1.01 – 8.42) | 0.049 | | | vs $M_4$ = 0.62 |
| 7 | Baseline age + RCT arm + $PGS_{269}$ + baseline $PSA^G$ | $PGS_{269}$ | 2.02 | (0.96 – 4.24) | 0.063 | 0.8484 | (0.7399 – 0.9569) | vs $M_4$ = 0.54 |
| | | log($PSA^G$) | 2.17 | (0.80 – 5.87) | 0.13 | | | vs $M_6$ = 0.91 |
| 8 | Baseline age + RCT arm + $PGS_{269}^{adj}$ + baseline $PSA^G$ | $PGS_{269}^{adj}$ | 1.59 | (0.81 – 3.14) | 0.18 | 0.8391 | (0.7248 – 0.9533) | vs $M_5$ = 0.59 |
| | | log($PSA^G$) | 2.14 | (0.81 – 5.65) | 0.13 | | | vs $M_7$ = 0.28 |

[1] Odds ratios were estimated per standard deviation increase in standardized $PGS_{269}$ and $PGS_{PSA}$. Models with genetic predictors ($PSA^G$, $PGS_{269}$, $PGS_{269}^{adj}$, and $PGS_{PSA}$) were adjusted for the top 10 genetic ancestry principal components and global ancestry proportions in the pooled analysis.

[2] Two-sided p-values based on De Long's test for two correlated ROC curves.

[3] African ancestry group includes men of African (AFR≥0.80) and admixed African and European (0.20<AFR/EUR<0.80) ancestry.

[4] East Asian group includes men of East Asian (AFR≥0.80) and admixed East Asian and European (0.20<EAS/EUR<0.80) ancestry.

**Supplementary Table 22: Comparison of logistic regression models for incident aggressive prostate cancer in the Selenium and Vitamin E Cancer Prevention Trial (SELECT).** Pooled analysis includes 106 cases and 23,667 cancer-free controls of all ancestries. Ancestry-stratified analyses were also performed. Models are indexed from $M_0$ to $M_8$. Odds ratios (OR), corresponding 95% confidence intervals, and two-sided p-values are presented for PSA variables and polygenic scores (PGS) only. AUC is based on the full model with all covariates.

| Models | | Predictor | Association Estimates | | | | | |
|---|---|---|---|---|---|---|---|---|
| **Pooled (Multi-ancestry)** | | | **OR**[1] | **(95% CI)** | **P** | **AUC** | **(95% CI)** | **$P_{AUC}$**[2] |
| 0 | Baseline age + RCT arm | - | - | - | - | 0.6090 | (0.5553 – 0.6628) | - |
| 1 | Baseline age + RCT arm + baseline PSA | log(PSA) | 3.31 | (2.38 – 4.60) | $1.1\times10^{-12}$ | 0.7375 | (0.6919 – 0.7831) | - |
| 2 | Baseline age + RCT arm + baseline $PSA^G$ | log($PSA^G$) | 3.44 | (2.49 – 4.75) | $6.2\times10^{-14}$ | 0.7549 | (0.7091 – 0.8008) | vs $M_1$ = 0.12 |
| 3 | Baseline age + RCT arm + $PGS_{PSA}$ | $PGS_{PSA}$ | 1.02 | (0.82 – 1.27) | 0.86 | 0.6579 | (0.5952 – 0.7006) | - |
| 4 | Baseline age + RCT arm + $PGS_{269}$ | $PGS_{269}$ | 1.73 | (1.42 – 2.11) | $8.5\times10^{-8}$ | 0.7120 | (0.6638 – 0.7077) | - |
| 5 | Baseline age + RCT arm + $PGS_{269}^{adj}$ | $PGS_{269}^{adj}$ | 1.76 | (1.44 – 2.14) | $1.7\times10^{-8}$ | 0.7198 | (0.6718 – 0.7677) | vs $M_4$ = $5.9\times10^{-3}$ |
| 6 | Baseline age + RCT arm + $PGS_{269}$ + baseline PSA | $PGS_{269}$<br>log(PSA) | 1.59<br>3.16 | (1.30 – 1.95)<br>(2.26 – 4.42) | $6.3\times10^{-6}$<br>$1.7\times10^{-11}$ | 0.7810 | (0.7392 – 0.8228) | vs $M_2$ = 0.056<br>vs $M_4$ = $3.1\times10^{-4}$ |
| 7 | Baseline age + RCT arm + $PGS_{269}$ + baseline $PSA^G$ | $PGS_{269}$<br>log($PSA^G$) | 1.70<br>3.46 | (1.39 – 2.07)<br>(2.49 – 4.81) | $2.2\times10^{-7}$<br>$1.5\times10^{-13}$ | 0.7861 | (0.7413 – 0.8309) | vs $M_4$ = $7.2\times10^{-4}$<br>vs $M_6$ = 0.51 |
| 8 | Baseline age + RCT arm + $PGS_{269}^{adj}$ + baseline $PSA^G$ | $PGS_{269}^{adj}$<br>log($PSA^G$) | 1.71<br>3.38 | (1.40 – 2.08)<br>(2.44 – 4.67) | $8.0\times10^{-8}$<br>$2.4\times10^{-13}$ | 0.7875 | (0.7436 – 0.8314) | vs $M_5$ = $1.3\times10^{-3}$<br>vs $M_7$ = 0.71 |
| **European (85 cases 20,173 controls)** | | | **OR**[1] | **(95% CI)** | **P** | **AUC** | **(95% CI)** | **$P_{AUC}$**[2] |
| 0 | Baseline age + RCT arm | - | - | - | - | 0.6045 | (0.5443 – 0.6648) | - |
| 1 | Baseline age + RCT arm + baseline PSA | log(PSA) | 3.40 | (2.34 – 4.93) | $1.1\times10^{-10}$ | 0.7420 | (0.6938 – 0.7901) | - |
| 2 | Baseline age + RCT arm + baseline $PSA^G$ | log($PSA^G$) | 3.81 | (2.62 – 5.54) | $2.5\times10^{-12}$ | 0.7766 | (0.7291 – 0.8242) | vs $M_1$ = 0.026 |
| 3 | Baseline age + RCT arm + $PGS_{PSA}$ | $PGS_{PSA}$ | 0.98 | (0.76 – 1.26) | 0.87 | 0.6768 | (0.6196 – 0.7339) | - |
| 4 | Baseline age + RCT arm + $PGS_{269}$ | $PGS_{269}$ | 1.76 | (1.41 – 2.21) | $7.4\times10^{-7}$ | 0.7264 | (0.6709 – 0.7819) | - |
| 5 | Baseline age + RCT arm + $PGS_{269}^{adj}$ | $PGS_{269}^{adj}$ | 1.81 | (1.45 – 2.25) | $1.3\times10^{-7}$ | 0.7301 | (0.6753 – 0.7849) | vs $M_4$ = 0.58 |
| 6 | Baseline age + RCT arm + $PGS_{269}$ + baseline PSA | $PGS_{269}$<br>log(PSA) | 1.61<br>3.20 | (1.28 – 2.02)<br>(2.19 – 4.67) | $3.7\times10^{-5}$<br>$1.5\times10^{-9}$ | 0.7956 | (0.7501 – 0.8410) | vs $M_2$ = 0.13<br>vs $M_4$ = $3.4\times10^{-4}$ |
| 7 | Baseline age + RCT arm + $PGS_{269}$ + baseline $PSA^G$ | $PGS_{269}$<br>log($PSA^G$) | 1.72<br>3.82 | (1.38 – 2.15)<br>(2.61 – 5.60) | $2.0\times10^{-6}$<br>$6.1\times10^{-12}$ | 0.8026 | (0.7545 – 0.8506) | vs $M_4$ = $6.8\times10^{-4}$<br>vs $M_6$ = 0.29 |
| 8 | Baseline age + RCT arm + $PGS_{269}^{adj}$ + baseline $PSA^G$ | $PGS_{269}^{adj}$<br>log($PSA^G$) | 1.74<br>3.69 | (1.40 – 2.16)<br>(2.53 – 5.38) | $7.3\times10^{-7}$<br>$1.0\times10^{-11}$ | 0.8039 | (0.7568 – 0.8510) | vs $M_5$ = $5.4\times10^{-4}$<br>vs $M_7$ = 0.26 |
| **African[3] (18 cases 2733 controls)** | | | **OR**[1] | **(95% CI)** | **P** | **AUC** | **(95% CI)** | **$P_{AUC}$**[2] |
| 0 | Baseline age + RCT arm | - | - | - | - | 0.7201 | (0.6015 – 0.8387) | - |
| 1 | Baseline age + RCT arm + baseline PSA | log(PSA) | 2.82 | (1.33 – 5.99) | $6.8\times10^{-3}$ | 0.7541 | (0.6219 – 0.8862) | - |

| | | | | | | | | |
|---|---|---|---|---|---|---|---|---|
| 2 | Baseline age + RCT arm + baseline $PSA^G$ [$PGS_{CSx}$] | log($PSA^G$) | 2.48 | (1.24 – 4.97) | 0.011 | 0.8066 | (0.6954 – 0.9178) | vs $M_1$ = 0.076 |
| | Baseline age + RCT arm + baseline $PSA^G$ [$PGS_{128}$] | log($PSA^G$) | 2.96 | (1.43 – 6.12) | $3.5\times10^{-3}$ | 0.8079 | (0.6991 – 0.9167) | vs $M_1$ = 0.051 |
| 3 | Baseline age + RCT arm + $PGS_{PSA}$ | $PGS_{PSA}$ | 1.09 | (0.67 – 1.79) | 0.72 | 0.7875 | (0.6998 – 0.8752) | - |
| 4 | Baseline age + RCT arm + $PGS_{269}$ | $PGS_{269}$ | 1.63 | (1.01 – 2.64) | 0.045 | 0.8030 | (0.7219 – 0.8841) | - |
| 5 | Baseline age + RCT arm + $PGS_{269}^{adj}$ | $PGS_{269}^{adj}$ | 1.62 | (1.02 – 2.59) | 0.042 | 0.8070 | (0.7243 – 0.8897) | vs $M_4$ = 0.54 |
| 6 | Baseline age + RCT arm + $PGS_{269}$ + baseline PSA | $PGS_{269}$ | 1.54 | (0.95 – 2.51) | 0.081 | 0.8277 | (0.7392 – 0.9162) | vs $M_2$ = 0.40 |
| | | log(PSA) | 2.75 | (1.28 – 5.90) | $9.5\times10^{-3}$ | | | vs $M_4$ = 0.36 |
| 7 | Baseline age + RCT arm + $PGS_{269}$ + baseline $PSA^G$ [$PGS_{CSx}$] | $PGS_{269}$ | 1.62 | (0.99 – 2.62) | 0.053 | 0.8246 | (0.7284 – 0.9207) | vs $M_4$ = 0.49 |
| | | log($PSA^G$) | 2.47 | (1.22 – 5.00) | 0.012 | | | vs $M_6$ = 0.83 |
| | Baseline age + RCT arm + $PGS_{269}$ + baseline $PSA^G$ [$PGS_{128}$] | $PGS_{269}$ | 1.61 | (0.99 – 2.60) | 0.052 | 0.8234 | (0.7239 – 0.9228) | vs $M_4$ = 0.50 |
| | | log($PSA^G$) | 2.97 | (1.41 – 6.27) | $4.1\times10^{-3}$ | | | vs $M_6$ = 0.68 |
| 8 | Baseline age + RCT arm + $PGS_{269}^{adj}$ + baseline $PSA^G$ | $PGS_{269}^{adj}$ | 1.61 | (1.00 – 2.57) | 0.048 | 0.8268 | (0.7304 – 0.9231) | vs $M_5$ = 0.52 |
| | | log($PSA^G$) | 2.46 | (1.22 – 4.98) | 0.012 | | | vs $M_7$ = 0.65 |

[1] Odds ratios were estimated per standard deviation increase in standardized PGS269 and PGSPSA. Models with genetic predictors ($PSA^G$, $PGS_{269}$, $PGS_{269}^{adj}$, and $PGS_{PSA}$) were adjusted for the top 10 genetic ancestry principal components and global ancestry proportions in the pooled analysis.

[2] Two-sided p-values based on De Long’s test for two correlated ROC curves.

[3] African ancestry group includes men of African (AFR≥0.80) and admixed African and European (0.20<AFR/EUR<0.80) ancestry.

**Supplementary Table 23: Comparison of logistic regression models for incident non-aggressive prostate cancer in the Selenium and Vitamin E Cancer Prevention Trial (SELECT).** Pooled analysis includes 466 cases and 23,667 cancer-free controls of all ancestries. Ancestry-stratified analyses were also performed. Models are indexed from $M_0$ to $M_8$. Odds ratios (OR), corresponding 95% confidence intervals, and two-sided p-values are presented for PSA variables and polygenic scores (PGS) only. AUC is based on the full model with all covariates.

| Models | | Predictor | Association Estimates | | | | | |
|---|---|---|---|---|---|---|---|---|
| **Pooled (Multi-ancestry)** | | | **OR[1]** | **(95% CI)** | **P** | **AUC** | **(95% CI)** | **$P_{AUC}$[2]** |
| 0 | Baseline age + RCT arm | - | - | - | - | 0.5254 | (0.5000 – 0.5507) | - |
| 1 | Baseline age + RCT arm + baseline PSA | log(PSA) | 4.03 | (3.42 – 4.74) | $6.3\times10^{-63}$ | 0.7428 | (0.7220 – 0.7636) | - |
| 2 | Baseline age + RCT arm + baseline $PSA^G$ | $\log(PSA^G)$ | 3.56 | (3.04 – 4.16) | $1.2\times10^{-56}$ | 0.7290 | (0.7071 – 0.7509) | vs $M_1$ = 0.017 |
| 3 | Baseline age + RCT arm + $PGS_{PSA}$ | $PGS_{PSA}$ | 1.28 | (1.15 – 1.42) | $4.0\times10^{-6}$ | 0.5776 | (0.5510 – 0.6043) | - |
| 4 | Baseline age + RCT arm + $PGS_{269}$ | $PGS_{269}$ | 1.74 | (1.58 – 1.92) | $2.5\times10^{-29}$ | 0.6569 | (0.6318 – 0.6821) | - |
| 5 | Baseline age + RCT arm + $PGS_{269}^{adj}$ | $PGS_{269}^{adj}$ | 1.62 | (1.47 – 1.78) | $1.3\times10^{-23}$ | 0.6445 | (0.6192 – 0.6698) | vs $M_4$ = $3.2\times10^{-3}$ |
| 6 | Baseline age + RCT arm + $PGS_{269}$ + baseline PSA | $PGS_{269}$<br>log(PSA) | 1.57<br>3.82 | (1.43 – 1.74)<br>(3.23 – 4.51) | $1.3\times10^{-19}$<br>$3.7\times10^{-56}$ | 0.7657 | (0.7450 – 0.7865) | vs $M_2$ = $1.5\times10^{-6}$<br>vs $M_4$ = $9.1\times10^{-23}$ |
| 7 | Baseline age + RCT arm + $PGS_{269}$ + baseline $PSA^G$ | $PGS_{269}$<br>$\log(PSA^G)$ | 1.69<br>3.55 | (1.54 – 1.87)<br>(3.02 – 4.17) | $2.7\times10^{-26}$<br>$3.8\times10^{-54}$ | 0.7606 | (0.7395 – 0.7817) | vs $M_4$ = $1.8\times10^{-21}$<br>vs $M_6$ = 0.24 |
| 8 | Baseline age + RCT arm + $PGS_{269}^{adj}$ + baseline $PSA^G$ | $PGS_{269}^{adj}$<br>$\log(PSA^G)$ | 1.57<br>3.49 | (1.43 – 1.73)<br>(2.98 – 4.09) | $1.4\times10^{-20}$<br>$3.6\times10^{-54}$ | 0.7551 | (0.7340 – 0.7762) | vs $M_5$ = $4.6\times10^{-22}$<br>vs $M_7$ = $5.9\times10^{-3}$ |
| **European (382 cases 20,173 controls)** | | | **OR[1]** | **(95% CI)** | **P** | **AUC** | **(95% CI)** | **$P_{AUC}$[2]** |
| 0 | Baseline age + RCT arm | - | - | - | - | 0.5209 | (0.4928 – 0.5491) | - |
| 1 | Baseline age + RCT arm + baseline PSA | log(PSA) | 4.12 | (3.43 – 4.94) | $2.2\times10^{-52}$ | 0.7430 | (0.7202 – 0.7658) | - |
| 2 | Baseline age + RCT arm + baseline $PSA^G$ | $\log(PSA^G)$ | 3.86 | (3.23 – 4.62) | $2.1\times10^{-49}$ | 0.7357 | (0.7119 – 0.7594) | vs $M_1$ = 0.26 |
| 3 | Baseline age + RCT arm + $PGS_{PSA}$ | $PGS_{PSA}$ | 1.27 | (1.13 – 1.44) | $1.0\times10^{-4}$ | 0.5792 | (0.5511 – 0.6073) | - |
| 4 | Baseline age + RCT arm + $PGS_{269}$ | $PGS_{269}$ | 1.82 | (1.63 – 2.03) | $1.0\times10^{-27}$ | 0.6669 | (0.6396 – 0.6942) | - |
| 5 | Baseline age + RCT arm + $PGS_{269}^{adj}$ | $PGS_{269}^{adj}$ | 1.68 | (1.51 – 1.86) | $5.0\times10^{-23}$ | 0.6522 | (0.6248 – 0.6796) | vs $M_4$ = $2.1\times10^{-3}$ |
| 6 | Baseline age + RCT arm + $PGS_{269}$ + baseline PSA | $PGS_{269}$<br>log(PSA) | 1.63<br>3.82 | (1.46 – 1.82)<br>(3.17 – 4.60) | $2.0\times10^{-19}$<br>$1.2\times10^{-45}$ | 0.7694 | (0.7470 – 0.7918) | vs $M_2$ = $7.1\times10^{-5}$<br>vs $M_4$ = $2.9\times10^{-19}$ |
| 7 | Baseline age + RCT arm + $PGS_{269}$ + baseline $PSA^G$ | $PGS_{269}$<br>$\log(PSA^G)$ | 1.76<br>3.86 | (1.58 – 1.96)<br>(3.21 – 4.63) | $1.0\times10^{-24}$<br>$5.6\times10^{-47}$ | 0.7684 | (0.7452 – 0.7916) | vs $M_4$ = $7.7\times10^{-19}$<br>vs $M_6$ = 0.83 |
| 8 | Baseline age + RCT arm + $PGS_{269}^{adj}$ + baseline $PSA^G$ | $PGS_{269}^{adj}$<br>$\log(PSA^G)$ | 1.61<br>3.77 | (1.45 – 1.79)<br>(3.15 – 4.52) | $7.3\times10^{-19}$<br>$7.2\times10^{-47}$ | 0.7620 | (0.7386 – 0.7853) | vs $M_5$ = $4.6\times10^{-22}$<br>vs $M_7$ = $5.9\times10^{-3}$ |
| **African[3] (70 cases 2733 controls)** | | | **OR[1]** | **(95% CI)** | **P** | **AUC** | **(95% CI)** | **$P_{AUC}$[2]** |
| 0 | Baseline age + RCT arm | - | - | - | - | 0.5602 | (0.4964 – 0.6240) | - |
| 1 | Baseline age + RCT arm + baseline PSA | log(PSA) | 3.90 | (2.60 – 5.86) | $5.5\times10^{-11}$ | 0.7565 | (0.7030 – 0.8099) | - |

| | | | | | | | | |
|---|---|---|---|---|---|---|---|---|
| 2 | Baseline age + RCT arm + baseline $PSA^G$ | $\log(PSA^G)$ | 2.75 | (1.92 – 3.94) | $3.6\times10^{-8}$ | 0.7385 | (0.6838 – 0.7931) | vs $M_1$ = 0.054 |
| 3 | Baseline age + RCT arm + $PGS_{PSA}$ | $PGS_{PSA}$ | 1.36 | (1.07 – 1.74) | 0.012 | 0.6634 | (0.5978 – 0.7290) | - |
| 4 | Baseline age + RCT arm + $PGS_{269}$ | $PGS_{269}$ | 1.39 | (1.09 – 1.77) | $7.9\times10^{-3}$ | 0.6630 | (0.5964 – 0.7295) | - |
| 5 | Baseline age + RCT arm + $PGS_{269}^{adj}$ | $PGS_{269}^{adj}$ | 1.37 | (1.08 – 1.74) | $8.6\times10^{-3}$ | 0.6666 | (0.6006 – 0.7324) | vs $M_4$ = 0.47 |
| 6 | Baseline age + RCT arm + $PGS_{269}$ + baseline PSA | $PGS_{269}$<br>log(PSA) | 1.29<br>3.97 | (1.01 – 1.66)<br>(2.62 – 6.03) | 0.046<br>$8.9\times10^{-11}$ | 0.7835 | (0.7288 – 0.8382) | vs $M_2$ = $1.6\times10^{-3}$<br>vs $M_4$ = $2.3\times10^{-5}$ |
| 7 | Baseline age + RCT arm + $PGS_{269}$ + baseline $PSA^G$ | $PGS_{269}$<br>$\log(PSA^G)$ | 1.37<br>2.73 | (1.07 – 1.75)<br>(1.90 – 3.92) | 0.014<br>$6.0\times10^{-8}$ | 0.7499 | (0.6946 – 0.8051) | vs $M_4$ = $5.3\times10^{-4}$<br>vs $M_6$ = $2.6\times10^{-3}$ |
| 8 | Baseline age + RCT arm + $PGS_{269}^{adj}$ + baseline $PSA^G$ | $PGS_{269}^{adj}$<br>$\log(PSA^G)$ | 1.36<br>2.73 | (1.06 – 1.73)<br>(1.90 – 3.92) | 0.014<br>$5.4\times10^{-8}$ | 0.7520 | (0.6977 – 0.8062) | vs $M_5$ = $8.2\times10^{-4}$<br>vs $M_7$ = 0.47 |

[1] Odds ratios were estimated per standard deviation increase in standardized PGS269 and PGSPSA. Models with genetic predictors ($PSA^G$, $PGS_{269}$, $PGS_{269}^{adj}$, and $PGS_{PSA}$) were adjusted for the top 10 genetic ancestry principal components and global ancestry proportions in the pooled analysis.

[2] Two-sided p-values based on De Long's test for two correlated ROC curves.

[3] African ancestry group includes men of African (AFR≥0.80) and admixed African and European (0.20<AFR/EUR<0.80) ancestry.

**Supplementary Table 24: Case-only analyses of prostate cancer aggressiveness in the Selenium and Vitamin E Cancer Prevention Trial (SELECT).** Pooled analysis includes 106 aggressive cases and 466 non-aggressive cases of all ancestries. Stratified analyses were performed in participants of European ancestry (85 aggressive, 382 non-aggressive) and African ancestry (18 aggressive, 70 non-aggressive). Models are indexed from $M_0$ to $M_8$. Odds ratios (OR), corresponding 95% confidence intervals, and two-sided p-values are presented for PSA variables and polygenic scores (PGS) only. AUC is based on the full model with all covariates.

| Models | | Predictor | Association Estimates | | | | | |
|---|---|---|---|---|---|---|---|---|
| **Pooled (Multi-ancestry)** | | | **OR[1]** | **(95% CI)** | **P** | **AUC** | **(95% CI)** | **$P_{AUC}$[2]** |
| 0 | Baseline age + RCT arm | - | - | - | - | 0.6383 | (0.5790 – 0.6975) | - |
| 1 | Baseline age + RCT arm + baseline PSA | log(PSA) | 0.89 | (0.61 – 1.30) | 0.54 | 0.6375 | (0.5790 – 0.6960) | - |
| 2 | Baseline age + RCT arm + baseline $PSA^G$ | log($PSA^G$) | 1.07 | (0.72 – 1.59) | 0.75 | 0.6565 | (0.5975 - 0.7156) | - |
| 3 | Baseline age + RCT arm + $PGS_{PSA}$ | $PGS_{PSA}$ | 0.79 | (0.61 – 1.03) | 0.078 | 0.6697 | (0.6118 - 0.7276) | - |
| 4 | Baseline age + RCT arm + $PGS_{269}$ | $PGS_{269}$ | 1.01 | (0.80 – 1.27) | 0.96 | 0.6554 | (0.5961 – 0.7147) | - |
| 5 | Baseline age + RCT arm + $PGS_{269}^{adj}$ | $PGS_{269}^{adj}$ | 1.10 | (0.87 – 1.39) | 0.42 | 0.6585 | (0.5990 – 0.7180) | - |
| 6 | Baseline age + RCT arm + $PGS_{269}$ + baseline PSA | $PGS_{269}$ | 1.02 | (0.80 – 1.29) | 0.90 | 0.6549 | (0.5956 – 0.7143) | - |
| | | log(PSA) | 0.89 | (0.60 – 1.31) | 0.55 | | | - |
| 7 | Baseline age + RCT arm + $PGS_{269}$ + baseline $PSA^G$ | $PGS_{269}$ | 1.00 | (0.79 – 1.27) | 0.98 | 0.6564 | (0.5973 – 0.7155) | - |
| | | log($PSA^G$) | 1.07 | (0.72 – 1.59) | 0.75 | | | - |
| 8 | Baseline age + RCT arm + $PGS_{269}^{adj}$ + baseline $PSA^G$ | $PGS_{269}^{adj}$ | 1.10 | (0.87 – 1.39) | 0.43 | 0.6600 | (0.6006 – 0.7194) | - |
| | | log($PSA^G$) | 1.06 | (0.71 – 1.57) | 0.78 | | | - |
| **European ancestry** | | | **OR[1]** | **(95% CI)** | **P** | **AUC** | **(95% CI)** | **$P_{AUC}$[2]** |
| 0 | Baseline age + RCT arm | - | - | - | - | 0.6302 | (0.5641 – 0.6962) | - |
| 1 | Baseline age + RCT arm + baseline PSA | log(PSA) | 0.92 | (0.62 – 1.42) | 0.71 | 0.6313 | (0.5654 – 0.6972) | - |
| 2 | Baseline age + RCT arm + baseline $PSA^G$ | log($PSA^G$) | 1.13 | (0.71 – 1.78) | 0.61 | 0.7015 | (0.6380 – 0.7650) | - |
| 3 | Baseline age + RCT arm + $PGS_{PSA}$ | $PGS_{PSA}$ | 0.75 | (0.55 – 1.02) | 0.065 | 0.7068 | (0.6447 – 0.7688) | - |
| 4 | Baseline age + RCT arm + $PGS_{269}$ | $PGS_{269}$ | 1.05 | (0.81 – 1.36) | 0.72 | 0.6983 | (0.6346 – 0.7621) | - |
| 5 | Baseline age + RCT arm + $PGS_{269}^{adj}$ | $PGS_{269}^{adj}$ | 1.17 | (0.90 – 1.51) | 0.24 | 0.7017 | (0.6369 – 0.7664) | - |
| 6 | Baseline age + RCT arm + $PGS_{269}$ + baseline PSA | $PGS_{269}$ | 0.89 | (0.56 – 1.41) | 0.62 | 0.6975 | (0.6339 – 0.7611) | - |
| | | log(PSA) | 1.06 | (0.81 – 1.38) | 0.67 | | | - |
| 7 | Baseline age + RCT arm + $PGS_{269}$ + baseline $PSA^G$ | $PGS_{269}$ | 1.12 | (0.71 – 1.78) | 0.63 | 0.7013 | (0.6373 – 0.7652) | - |
| | | log($PSA^G$) | 1.04 | (0.80 – 1.35) | 0.76 | | | - |
| 8 | Baseline age + RCT arm + $PGS_{269}^{adj}$ + baseline $PSA^G$ | $PGS_{269}^{adj}$ | 1.10 | (0.70 – 1.75) | 0.67 | 0.7039 | (0.6391 – 0.7687) | - |
| | | log($PSA^G$) | 1.16 | (0.90 – 1.50) | 0.26 | | | - |

| African ancestry | | | OR[1] | (95% CI) | P | AUC | (95% CI) | $P_{AUC}$[2] |
|---|---|---|---|---|---|---|---|---|
| 0 | Baseline age + RCT arm | - | - | - | - | 0.7201 | (0.5840 – 0.8580) | - |
| 1 | Baseline age + RCT arm + baseline PSA | log(PSA) | 0.66 | (0.27 – 1.61) | 0.37 | 0.7298 | (0.6036 – 0.8559) | - |
| 2 | Baseline age + RCT arm + baseline $PSA^{G}$ | log($PSA^{G}$) | 0.83 | (0.29 – 2.38) | 0.73 | 0.8254 | (0.7260 – 0.9248) | - |
| 3 | Baseline age + RCT arm + $PGS_{PSA}$ | $PGS_{PSA}$ | 0.64 | (0.31 – 1.33) | 0.23 | 0.8294 | (0.7241 – 0.9346) | - |
| 4 | Baseline age + RCT arm + $PGS_{269}$ | $PGS_{269}$ | 0.99 | (0.42 – 2.35) | 0.98 | 0.8222 | (0.7212 – 0.9233) | - |
| 5 | Baseline age + RCT arm + $PGS_{269}^{adj}$ | $PGS_{269}^{adj}$ | 0.98 | (0.40 – 2.40) | 0.97 | 0.8222 | (0.7210 – 0.9234) | - |
| 6 | Baseline age + RCT arm + $PGS_{269}$ + baseline PSA | $PGS_{269}$ | 0.94 | (0.39 – 2.23) | 0.88 | 0.8437 | (0.7491 – 0.9382) | - |
| | | log(PSA) | 0.55 | (0.19 – 1.61) | 0.27 | | | - |
| 7 | Baseline age + RCT arm + $PGS_{269}$ + baseline $PSA^{G}$ | $PGS_{269}$ | 0.95 | (0.39 – 2.30) | 0.92 | 0.8270 | (0.7291 – 0.9249) | - |
| | | log($PSA^{G}$) | 0.82 | (0.28 – 2.40) | 0.71 | | | - |
| 8 | Baseline age + RCT arm + $PGS_{269}^{adj}$ + baseline $PSA^{G}$ | $PGS_{269}^{adj}$ | 0.93 | (0.37 – 2.36) | 0.89 | 0.8278 | (0.7301 – 0.9255) | - |
| | | log($PSA^{G}$) | 0.81 | (0.27 – 2.42) | 0.70 | | | - |

[1] Odds ratios were estimated per standard deviation increase in standardized PGS269 and PGSPSA. Models with genetic predictors ($PSA^{G}$, $PGS_{269}$, $PGS_{269}^{adj}$, and $PGS_{PSA}$) were adjusted for the top 10 genetic ancestry principal components and global ancestry proportions in the pooled analysis.

[2] Two-sided p-values based on De Long's test for two correlated ROC curves.

[3] African ancestry group includes men of African (AFR≥0.80) and admixed African and European (0.20<AFR/EUR<0.80) ancestry.

**Supplementary Table 25:** Field codes used to extract PSA values from linked GP records in the UK Biobank

| HES Field code | Description |
|---|---|
| X80QD | Prostate specific antigen |
| XE25C | Prostate-specific antigen level |
| XabAM | Serum prostate specific antigen level |
| XaPqN | Total prostate specific antigen level |
| XaQ9n | Ultra-sensitive prostate specific antigen level |

**Supplementary Table 26:** Study-specific GWAS approach and adjustment covariates

| Study | GWAS Phenotype | Analysis | Covariates |
|---|---|---|---|
| UKB | Median log(PSA) | Linear regression | Age at PSA measurement, baseline BMI, baseline smoking status (never, current, former smokers), diagnosed with benign prostatic hyperplasia (BPH) or prostatitis (1=ever, 0=never), time between median PSA and BPH or prostatitis diagnosis (coded as 0 for those without a corresponding diagnosis), finasteride use (1=ever, 0=never), genotyping array (UK BiLEVE vs. UKB Affymetrix Axiom), PC1-PC15 |
| | Random Intercept | Linear mixed model | Age at each observation, baseline BMI, baseline smoking status (never, current, former smokers), diagnosed with benign prostatic hyperplasia (BPH) or prostatitis (1=ever, 0=never), finasteride use (1=ever, 0=never), genotyping array (UK BiLEVE vs. UKB Affymetrix Axiom), PC1-PC15 |
| GERA | Random Intercept | Linear mixed model | Age at each observation, composite variable for DNA collection kit and genotyping array (chip-kit), PC1-PC10 |
| PLCO | Baseline log(PSA) | Linear mixed model | Age at PSA measurement, baseline BMI, baseline smoking status (never, current, former smokers), benign prostatic hyperplasia (surgically-treated BPH, BPH without surgery, none) and ancestry-specific PC1-PC10. Residuals and models were created and run separately by ancestry, genotype array, and imputation batch. |
| BioVU | Median log(PSA) | Linear regression | Median age of PSA measurements, median BMI, diagnosed with BPH (1=ever, 0=never), testosterone use (1=ever, 0=never), finasteride use (1=ever, 0=never), PC1-PC10 |
| MDCS | Baseline log(PSA) | Linear regression | Age at blood draw for PSA measurement, genotyping batch, PC1-PC10 |
